# Trends in Cardiovascular Risk Factors, Disease Management, and Predicted Cardiovascular Risk Among US Adults, 1999–2023

**DOI:** 10.64898/2026.11.09.26367683

**Authors:** Hafiz M Ahmed, Nawal Younas, Muhammad Owais

## Abstract

**Background:** Cardiovascular risk factors have changed in the United States, but trends in prevalence, management, and predicted risk through the post-pandemic period remain incompletely characterized.

**Methods:** We analyzed 11 National Health and Nutrition Examination Survey (NHANES) periods (1999–2000 through 2021–2023) among 60,199 nonpregnant adults aged ≥20 years. Survey-weighted regression assessed age-standardized trends, subgroup differences, and measurement-change sensitivity. We assessed awareness, treatment, and control of hypertension, dyslipidemia, and diabetes. PREVENT equations estimated 10- and 30-year risks among adults without self-reported cardiovascular disease (CVD).

**Results:** Obesity increased from 47.4% to 57.2% (odds ratio [OR] per decade, 1.23; 95% CI, 1.16–1.30), with class III obesity more than doubling (4.7% to 9.7%). Diabetes increased from 8.2% to 12.7% (OR, 1.26; 95% CI, 1.19–1.34), while current smoking declined from 24.4% to 14.9% (OR, 0.76; 95% CI, 0.71–0.81). Hypertension, dyslipidemia, chronic kidney disease, and prior CVD showed no significant linear trends, although mean systolic blood pressure (BP) (124.2 to 120.2 mm Hg) and total cholesterol (203.6 to 188.1 mg/dL) declined, alongside improvements in other lipids. Hypertension and dyslipidemia awareness and treatment increased, but BP control peaked at 32.5% in 2013–2014 before declining to 21.5%, while LDL cholesterol control rose from 17.0% to 34.2%. Diabetes awareness increased, but treatment trends were nonsignificant. Glycemic control peaked at 51.2% in 2003–2004 before declining to 41.9% (P for nonlinearity <0.001). Predicted 10-year total CVD risk declined from 5.66% to 5.28%; atherosclerotic CVD (ASCVD) risk declined by 0.16 percentage points per decade (95% CI, −0.22 to −0.09), while heart failure (HF) risk increased by 0.10 (95% CI, 0.03–0.17). Their 2021–2023 estimates were nearly identical (3.20% and 3.19%, respectively); 30-year risks showed similar divergence.

**Conclusions:** Improvements in smoking, blood pressure, and lipids coexisted with rising obesity and diabetes, declining blood pressure control after earlier gains, and divergent predicted ASCVD and HF risks.

## 1. INTRODUCTION

Cardiovascular disease (CVD) remains the leading cause of death in the United States ^1^. After decades of decline, age-adjusted CVD mortality began to level off around 2011 and increased during the COVID-19 pandemic ^2,3^. Improvements in major cardiovascular risk factors, particularly cholesterol, blood pressure (BP), and smoking, accounted for nearly half of the earlier decline in coronary deaths ^4,5^. These modifiable factors contribute substantially to myocardial infarction risk at the population level and are central to the American Heart Association’s Life’s Essential 8 framework ^6,7^. Tracking their trends is therefore essential to understanding progress in cardiovascular prevention.

Previous analyses of the National Health and Nutrition Examination Survey (NHANES) have shown increasing obesity and diabetes, declining smoking and cholesterol levels, and improvements in BP control that were subsequently reversed through 2018 or March 2020 ^8–11^. More recently, the National Center for Health Statistics (NCHS) reported estimates for individual risk factors, including obesity and cholesterol, from August 2021 through August 2023, the first post-pandemic NHANES cycle ^12,13^ . However, changes in sampling design and medication questionnaires during this cycle pose challenges for comparisons with earlier periods ^14^. Meanwhile, the American Heart Association’s PREVENT equations offer a framework for estimating 10- and 30-year risks of total CVD, atherosclerotic cardiovascular disease (ASCVD), and heart failure (HF) using cardiovascular-kidney-metabolic risk factors ^15–17^.

Recent studies have explored the clustering of cardiovascular risk factors, cardiometabolic risk and its management among adults with hypertension, and population-level risk estimates using the PREVENT equations ^18–21^. Nevertheless, few studies have examined major cardiovascular risk factors alongside their awareness, treatment, and control and predicted cardiovascular risk over the entire 1999–2023 period. Comparisons across NHANES cycles are further complicated by changes in HDL cholesterol assays, BP measurement devices, and medication questionnaires, which may affect observed trends. Examining these trends together, while accounting for changes in measurement, may provide a clearer picture of progress in cardiovascular prevention and help identify areas where important gaps remain.

In this study, we examined trends in major cardiovascular risk factors among US adults from 1999–2000 through August 2021–August 2023. We assessed changes in the awareness, treatment, and control of hypertension, dyslipidemia, and diabetes, along with PREVENT-predicted cardiovascular risk. Using harmonized definitions, we evaluated the potential influence of measurement changes on observed trends and examined differences by sex, age, race and Hispanic origin, income, and education.

## 2. METHODS

### 2.1. Study Design and Population

We analyzed data from 11 National Health and Nutrition Examination Survey (NHANES) periods spanning 1999–2000 through 2021–2023. NHANES uses a stratified, multistage probability sampling design to obtain nationally representative estimates of the civilian, noninstitutionalized US population. Participants complete household interviews and undergo standardized examinations at mobile examination centers (MECs). Following the interruption of data collection in March 2020, the National Center for Health Statistics (NCHS) combined the 2017–2018 data with the partial 2019–2020 data to create the 2017–March 2020 pre-pandemic file **(eMethods)** ^14,22–25^.

The study included nonpregnant adults aged 20 years or older who completed an MEC examination. Each survey period was treated as an independent cross-sectional sample, with analytic denominators varying according to the availability of data for each outcome. The study followed the STROBE and RECORD reporting guidelines ^26,27^. Detailed eligibility criteria and sample sizes are presented in **eFigure 1** and **eTables 1–2**.

### 2.2. Outcomes and Harmonization

The primary outcomes were obesity, hypertension, dyslipidemia, diabetes, chronic kidney disease (CKD), current smoking, alcohol consumption, and prior cardiovascular disease (CVD). Obesity was defined as a body mass index (BMI) ≥30 kg/m² or waist circumference >102 cm in men or >88 cm in women ^28^. Hypertension was defined as blood pressure ≥130/80 mm Hg, antihypertensive medication use, or self-reported diagnosis ^29^. Dyslipidemia was defined as total cholesterol ≥240 mg/dL, low high-density lipoprotein (HDL) cholesterol (<40 mg/dL in men or <50 mg/dL in women), lipid-lowering medication use, or self-reported high cholesterol ^30^. Diabetes was defined as self-reported diagnosis, glycated hemoglobin (HbA1c) ≥6.5%, or glucose-lowering medication use ^31^. CKD was defined as estimated glomerular filtration rate (eGFR) <60 mL/min/1.73 m², urine albumin-to-creatinine ratio ≥30 mg/g, self-reported weak or failing kidneys, or dialysis ^32^. eGFR was calculated using the race-free 2021 CKD Epidemiology Collaboration equation ^33,34^ . Because kidney function was assessed at a single examination, CKD estimates were based on surveillance criteria rather than confirmation of persistent disease (**eMethods**).

Additional outcomes included BMI-defined obesity classes, mean clinical and laboratory measurements. Prior CVD was defined as self-reported heart failure, coronary heart disease, angina, myocardial infarction, or stroke. Among adults meeting the respective disease definitions, we also assessed awareness, treatment, and control based on self-reported diagnosis, medication use, and established clinical thresholds (**eMethods**).

Outcome definitions and measurements were harmonized across survey periods to account for changes in laboratory assays, blood pressure measurement devices, and medication questionnaires. Detailed definitions and harmonization procedures are provided in **eMethods**.

### 2.3. Predicted Cardiovascular Risk

We used the base American Heart Association PREVENT equations to estimate 10-year risks of total CVD, atherosclerotic cardiovascular disease (ASCVD), and heart failure among adults aged 30–79 years without self-reported CVD and with complete input data ^15,16^. We also estimated 30-year risks among adults aged 30–59 years. The equations incorporate demographic, clinical, and laboratory measures, including blood pressure, cholesterol, diabetes, smoking, kidney function, and BMI. Input values outside the equations’ valid ranges were capped at the nearest limit. Risk estimates were age-standardized, with further details provided in **eMethods**.

### 2.4. Statistical Analysis

All analyses accounted for the complex NHANES survey design using the appropriate examination, phlebotomy, or fasting subsample weights, along with masked strata and primary sampling units. Variances were estimated using Taylor series linearization. Prevalence and mean estimates were directly age-standardized to the 2000 US standard population using age groups of 20–39, 40–59, and ≥60 years, with modified age groups for PREVENT risk estimates ^35^.

Confidence intervals for proportions were calculated using the Korn–Graubard method, and estimate reliability was evaluated according to NCHS standards (**eMethods**) ^36,37^.

We assessed temporal trends using pooled survey-weighted logistic regression for binary outcomes and linear regression for continuous outcomes, adjusting for age group. Survey-cycle midpoints were modeled as continuous time in decades, and survey weights were scaled to reflect differences in the duration of each survey period. Trends were reported as odds ratios, mean differences, or ratios per decade, as appropriate. We included quadratic time terms to assess nonlinearity and calculated absolute changes as the difference between estimates from the first and last survey cycles. To identify changes in the direction or magnitude of trends, we also performed joinpoint regression on age-standardized estimates, weighted by inverse variance and allowing a maximum of one joinpoint (**eMethods**) ^38^.

Subgroup analyses were conducted by sex, age, race and Hispanic origin, family income-to-poverty ratio, and education. Differences in temporal trends between subgroups were evaluated using time-by-subgroup interaction terms and design-based Wald tests. Sensitivity analyses examined alternative outcome definitions, changes in measurement methods and questionnaires, complete-case restrictions, missing-data adjustments, Asian-specific BMI thresholds, and exclusion of the 2021–2023 survey period **(eMethods; eTables 5 and 7–9; eFigures 10–12, 16–17)** ^39^.

All statistical tests were two-sided, with P<.05 considered statistically significant. P values were not adjusted for multiple comparisons; subgroup and sensitivity analyses should be considered exploratory. Analyses were performed using R version 4.5.2. Additional details of the statistical methods are provided in **eMethods**.

## 3. RESULTS

### 3.1. Study population

Of 119,555 NHANES participants, 66,553 were aged 20 years or older, of whom 61,741 underwent examination. After excluding 1,542 pregnant participants, 60,199 adults were included in the analysis **(eFigure 1; eTable 1)**. From 1999–2000 to 2021–2023, mean age increased from 46.4 to 49.0 years, the proportion aged 60 years or older rose from 24.1% to 31.4%, and the proportion with a college degree increased from 21.8% to 34.0% **(Table 1)**. Unless otherwise specified, prevalence estimates are age-standardized, comparisons are between the first and last survey cycles, and odds ratios (ORs) represent changes per decade.

**Table 1.**
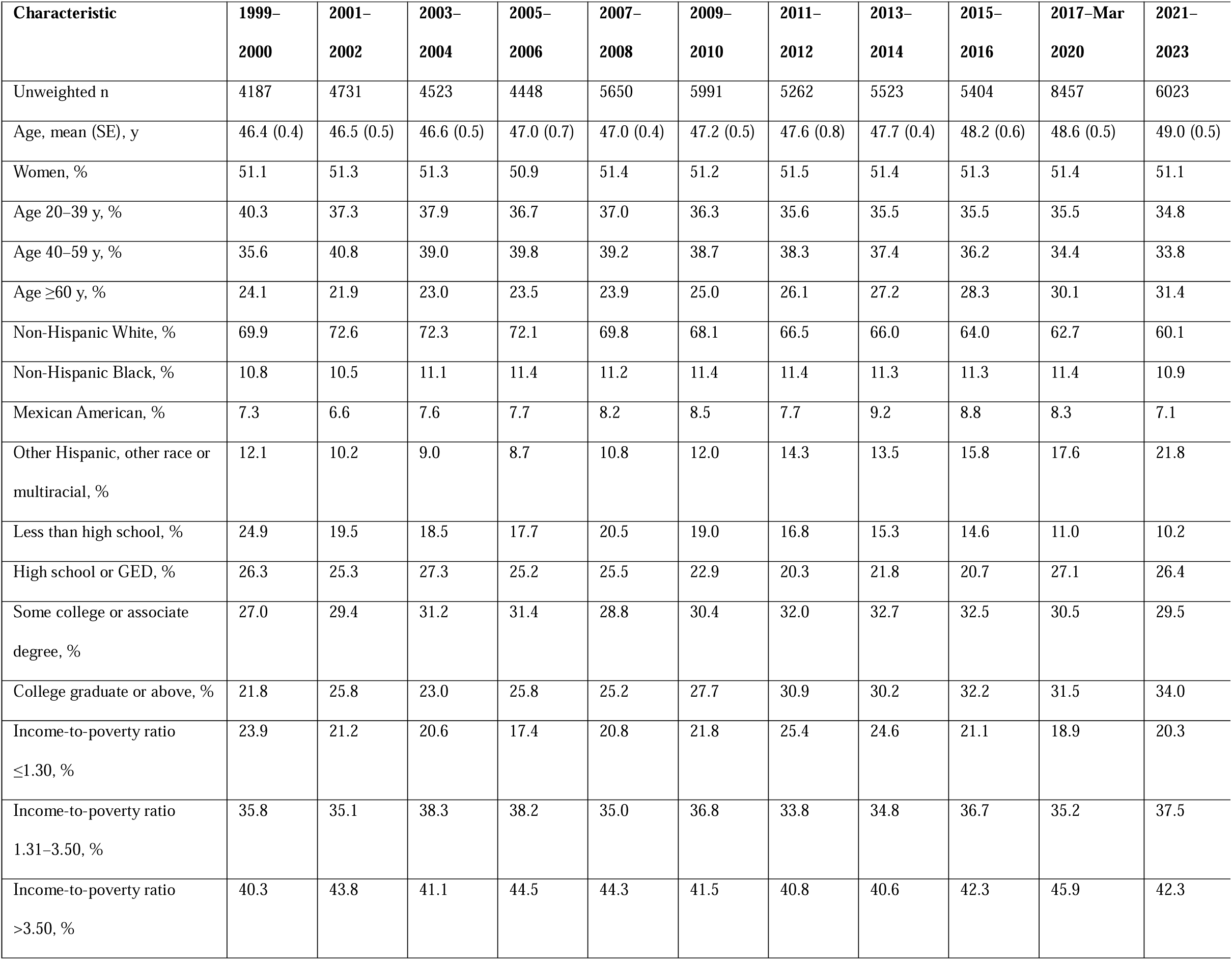
Characteristics of US adults aged ≥20 years, NHANES 1999–2000 to August 2021–August 2023. Demographic and socioeconomic characteristics of US adults aged ≥20 years across NHANES cycles from 1999–2000 through August 2021–August 2023. Values are unweighted sample sizes, survey-weighted means (SEs), or survey-weighted percentages. Pregnant participants were excluded. **Abbreviations:** NHANES, National Health and Nutrition Examination Survey; GED, General Educational Development; SE, standard error.

| **Characteristic** | **1999–2000** | **2001–2002** | **2003–2004** | **2005–2006** | **2007–2008** | **2009–2010** | **2011–2012** | **2013–2014** | **2015–2016** | **2017–Mar 2020** | **2021–2023** |
|---|---|---|---|---|---|---|---|---|---|---|---|
| Unweighted n | 4187 | 4731 | 4523 | 4448 | 5650 | 5991 | 5262 | 5523 | 5404 | 8457 | 6023 |
| Age, mean (SE), y | 46.4 (0.4) | 46.5 (0.5) | 46.6 (0.5) | 47.0 (0.7) | 47.0 (0.4) | 47.2 (0.5) | 47.6 (0.8) | 47.7 (0.4) | 48.2 (0.6) | 48.6 (0.5) | 49.0 (0.5) |
| Women, % | 51.1 | 51.3 | 51.3 | 50.9 | 51.4 | 51.2 | 51.5 | 51.4 | 51.3 | 51.4 | 51.1 |
| Age 20–39 y, % | 40.3 | 37.3 | 37.9 | 36.7 | 37.0 | 36.3 | 35.6 | 35.5 | 35.5 | 35.5 | 34.8 |
| Age 40–59 y, % | 35.6 | 40.8 | 39.0 | 39.8 | 39.2 | 38.7 | 38.3 | 37.4 | 36.2 | 34.4 | 33.8 |
| Age ≥60 y, % | 24.1 | 21.9 | 23.0 | 23.5 | 23.9 | 25.0 | 26.1 | 27.2 | 28.3 | 30.1 | 31.4 |
| Non-Hispanic White, % | 69.9 | 72.6 | 72.3 | 72.1 | 69.8 | 68.1 | 66.5 | 66.0 | 64.0 | 62.7 | 60.1 |
| Non-Hispanic Black, % | 10.8 | 10.5 | 11.1 | 11.4 | 11.2 | 11.4 | 11.4 | 11.3 | 11.3 | 11.4 | 10.9 |
| Mexican American, % | 7.3 | 6.6 | 7.6 | 7.7 | 8.2 | 8.5 | 7.7 | 9.2 | 8.8 | 8.3 | 7.1 |
| Other Hispanic, other race or multiracial, % | 12.1 | 10.2 | 9.0 | 8.7 | 10.8 | 12.0 | 14.3 | 13.5 | 15.8 | 17.6 | 21.8 |
| Less than high school, % | 24.9 | 19.5 | 18.5 | 17.7 | 20.5 | 19.0 | 16.8 | 15.3 | 14.6 | 11.0 | 10.2 |
| High school or GED, % | 26.3 | 25.3 | 27.3 | 25.2 | 25.5 | 22.9 | 20.3 | 21.8 | 20.7 | 27.1 | 26.4 |
| Some college or associate degree, % | 27.0 | 29.4 | 31.2 | 31.4 | 28.8 | 30.4 | 32.0 | 32.7 | 32.5 | 30.5 | 29.5 |
| College graduate or above, % | 21.8 | 25.8 | 23.0 | 25.8 | 25.2 | 27.7 | 30.9 | 30.2 | 32.2 | 31.5 | 34.0 |
| Income-to-poverty ratio ≤1.30, % | 23.9 | 21.2 | 20.6 | 17.4 | 20.8 | 21.8 | 25.4 | 24.6 | 21.1 | 18.9 | 20.3 |
| Income-to-poverty ratio 1.31–3.50, % | 35.8 | 35.1 | 38.3 | 38.2 | 35.0 | 36.8 | 33.8 | 34.8 | 36.7 | 35.2 | 37.5 |
| Income-to-poverty ratio >3.50, % | 40.3 | 43.8 | 41.1 | 44.5 | 44.3 | 41.5 | 40.8 | 40.6 | 42.3 | 45.9 | 42.3 |

### 3.2. Obesity

Obesity, defined by BMI or waist circumference, increased from 47.4% to 57.2% (OR, 1.23; 95% CI, 1.16–1.30) **(Figure 1**; **Table 2)**. Over the same period, mean BMI rose from 28.0 to 29.7 kg/m² **(eFigure 3)**. The prevalence of class III obesity more than doubled, from 4.7% to 9.7%, with the rate of increase accelerating after 2007–2008 **(eFigure 2; eTable 6)**.

**Figure 1.**
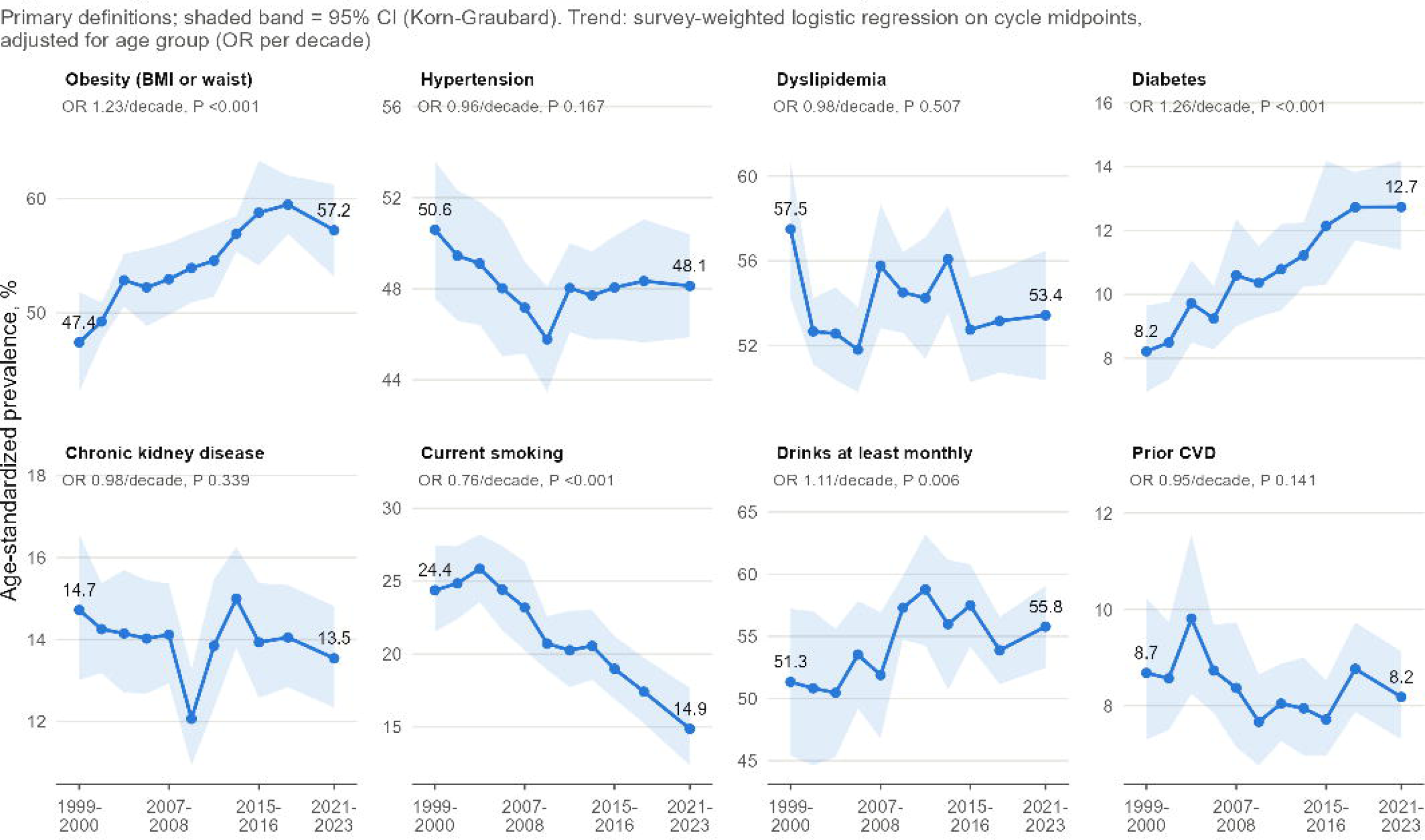
Age-standardized prevalence of major cardiovascular risk factors among US adults, NHANES 1999–2000 to 2021–2023. Panels show trends in obesity (BMI ≥30 kg/m² or waist circumference >102 cm in men or >88 cm in women), hypertension, dyslipidemia, diabetes, chronic kidney disease (CKD), current smoking, alcohol consumption at least monthly during the past year, and prior cardiovascular disease (CVD). Points represent estimates for each survey period, with shaded bands indicating 95% confidence intervals (CIs). Odds ratios (ORs) per decade and P values were obtained from survey-weighted logistic regression adjusted for age group. Outcome definitions are detailed in the Methods.

**Table 2.** Age-standardized prevalence and mean levels of cardiovascular risk factors among US adults, NHANES 1999–2000 to 2021–2023, with trends per decade. Estimates were age-standardized to the 2000 US standard population, with PREVENT risk estimates standardized within their respective eligible age ranges. Absolute change was calculated as the difference between the 2021–2023 and first-cycle estimates. Trends were assessed using survey-weighted regression adjusted for age group and expressed as odds ratios (ORs), mean differences, or ratios per decade, as appropriate. P values for nonlinearity were based on a quadratic time term. LDL cholesterol, triglycerides, and glucose were assessed in the fasting subsample. Treatment and control estimates were restricted to adults meeting the primary definition of each condition. Dialysis was first assessed in 2001–2002; therefore, its absolute change and trend were calculated from 2001–2002 to 2021–2023. **Abbreviations:** ASCVD, atherosclerotic cardiovascular disease; BMI, body mass index; BP, blood pressure; CI, confidence interval; CKD, chronic kidney disease; CVD, cardiovascular disease; DBP, diastolic blood pressure; eGFR, estimated glomerular filtration rate; HbA1c, glycated hemoglobin; HDL, high-density lipoprotein; LDL, low-density lipoprotein; OR, odds ratio; PREVENT, Predicting Risk of Cardiovascular Disease Events; SBP, systolic blood pressure; TC, total cholesterol; TG, triglycerides; UACR, urine albumin-to-creatinine ratio.

| **Outcome** | **1999–2000** | **2021–2023** | **Change (95% CI)** | **Trend measure** | **Per decade (95% CI)** | **P for trend** | **P for nonlinearity** |
|---|---|---|---|---|---|---|---|
| **Adiposity** |  |  |  |  |  |  |  |
| Obesity (BMI ≥30 or waist >102 cm in men or >88 cm in women), % | 47.4 | 57.2 | 9.8 (4.2 to 15.3) | OR | 1.23 (1.16–1.30) | <0.001 | 0.072 |
| Obesity by BMI ≥30 alone, % | 30.2 | 40.3 | 10.2 (5.2 to 15.2) | OR | 1.28 (1.21–1.35) | <0.001 | 0.356 |
| Abdominal obesity by waist circumference alone, % | 46.6 | 56.2 | 9.6 (4.1 to 15.2) | OR | 1.22 (1.15–1.29) | <0.001 | 0.035 |
| Underweight (BMI <18.5), % | 2.1 | 1.6 | −0.4 (−1.2 to 0.3) | OR | 0.88 (0.78–1.00) | 0.046 | 0.457 |
| Normal weight (BMI 18.5–24.9), % | 33.7 | 26.3 | −7.5 (−11.8 to −3.1) | OR | 0.83 (0.79–0.88) | <0.001 | 0.672 |
| Overweight (BMI 25.0–29.9), % | 34.0 | 31.7 | −2.3 (−5.2 to 0.7) | OR | 0.93 (0.90–0.97) | <0.001 | 0.922 |
| Obesity class I (BMI 30.0–34.9), % | 17.1 | 20.3 | 3.2 (0.5 to 5.9) | OR | 1.11 (1.06–1.16) | <0.001 | 0.022 |
| Obesity class II (BMI 35.0–39.9), % | 8.3 | 10.3 | 2.0 (0.2 to 3.8) | OR | 1.21 (1.13–1.28) | <0.001 | 0.920 |
| Obesity class III (BMI ≥40), % | 4.7 | 9.7 | 5.0 (3.2 to 6.9) | OR | 1.47 (1.35–1.59) | <0.001 | 0.838 |
| Mean BMI, kg/m² | 28.0 | 29.7 | 1.7 (1.0 to 2.4) | Difference | 0.89 (0.70–1.08) | <0.001 | 0.940 |
| Mean waist circumference, cm | 95.6 | 100.2 | 4.5 (2.7 to 6.3) | Difference | 2.20 (1.73–2.67) | <0.001 | 0.106 |
| **Hypertension and blood pressure** |  |  |  |  |  |  |  |
| Hypertension (BP ≥130/80 mm Hg, medication, or self-reported diagnosis), % | 50.6 | 48.1 | −2.5 (−6.0 to 1.1) | OR | 0.96 (0.92–1.02) | 0.167 | 0.054 |
| Measured BP ≥130/80 mm Hg, % | 44.0 | 37.2 | −6.8 (−10.1 to −3.5) | OR | 0.88 (0.84–0.92) | <0.001 | <0.001 |
| Antihypertensive medication use, % | 17.6 | 22.2 | 4.6 (2.2 to 7.1) | OR | 1.14 (1.08–1.20) | <0.001 | <0.001 |
| Self-reported hypertension diagnosis, % | 25.5 | 28.8 | 3.3 (0.4 to 6.2) | OR | 1.08 (1.03–1.14) | 0.003 | <0.001 |
| Mean systolic BP, mm Hg | 124.2 | 120.2 | −4.0 (−5.5 to −2.5) | Difference | −1.52 (−1.92 to −1.13) | <0.001 | 0.100 |
| Mean diastolic BP, mm Hg | 73.1 | 74.9 | 1.8 (0.8 to 2.7) | Difference | 1.02 (0.72–1.31) | <0.001 | <0.001 |
| **Dyslipidemia and lipids** |  |  |  |  |  |  |  |
| Dyslipidemia (total cholesterol ≥240 mg/dL, low HDL, medication, or self-reported diagnosis), % | 57.5 | 53.4 | −4.1 (−8.3 to 0.1) | OR | 0.98 (0.94–1.03) | 0.507 | 0.956 |
| Total cholesterol ≥240 mg/dL, % | 17.8 | 11.0 | −6.7 (−9.2 to −4.2) | OR | 0.73 (0.69–0.78) | <0.001 | 0.101 |
| Low HDL (<40 mg/dL in men, <50 mg/dL in women), % | 39.3 | 26.7 | −12.7 (−17.4 to −7.9) | OR | 0.86 (0.82–0.91) | <0.001 | 0.383 |
| Lipid-lowering medication use, % | 8.8 | 17.8 | 9.0 (7.5 to 10.5) | OR | 1.30 (1.24–1.36) | <0.001 | <0.001 |
| Self-reported high cholesterol, % | 25.9 | 33.3 | 7.3 (5.1 to 9.5) | OR | 1.19 (1.14–1.24) | <0.001 | 0.002 |
| Extended dyslipidemia (primary definition plus fasting LDL ≥160 mg/dL or triglycerides ≥150 mg/dL), % | 67.8 | 57.4 | −10.5 (−14.8 to −6.1) | OR | 0.87 (0.83–0.92) | <0.001 | 0.019 |
| Fasting LDL ≥160 mg/dL, % | 15.9 | 8.6 | −7.3 (−10.8 to −3.8) | OR | 0.75 (0.68–0.83) | <0.001 | 0.027 |
| Fasting triglycerides ≥150 mg/dL, % | 33.1 | 20.9 | −12.2 (−16.0 to −8.4) | OR | 0.68 (0.64–0.72) | <0.001 | 0.156 |
| Mean total cholesterol, mg/dL | 203.6 | 188.1 | −15.5 (−18.9 to −12.1) | Difference | −8.02 (−9.05 to −6.99) | <0.001 | 0.049 |
| Mean HDL cholesterol, mg/dL | 50.3 | 53.3 | 3.0 (1.3 to 4.7) | Difference | 0.92 (0.50–1.34) | <0.001 | 0.011 |
| Mean non-HDL cholesterol, mg/dL | 153.3 | 134.8 | −18.5 (−22.3 to −14.7) | Difference | −8.93 (−9.97 to −7.89) | <0.001 | 0.002 |
| Mean fasting LDL cholesterol, mg/dL | 126.0 | 110.5 | −15.5 (−18.8 to −12.2) | Difference | −5.78 (−6.78 to −4.77) | <0.001 | <0.001 |
| Mean fasting triglycerides, mg/dL | 143.4 | 118.7 | −24.6 (−35.0 to −14.3) | Difference | −18.45 (−21.53 to −15.37) | <0.001 | 0.018 |
| Geometric mean fasting triglycerides, mg/dL | 121.5 | 99.9 | −21.6 (−28.0 to −15.2) | Ratio | 0.87 (0.86–0.88) | <0.001 | 0.002 |
| **Diabetes and glycemia** |  |  |  |  |  |  |  |
| Diabetes (self-reported diagnosis, HbA1c ≥6.5%, or medication), % | 8.2 | 12.7 | 4.5 (2.7 to 6.3) | OR | 1.26 (1.19–1.34) | <0.001 | 0.211 |
| Self-reported diagnosed diabetes, % | 6.6 | 10.6 | 4.1 (2.5 to 5.7) | OR | 1.28 (1.21–1.37) | <0.001 | 0.262 |
| HbA1c ≥6.5%, % | 6.4 | 9.7 | 3.2 (1.5 to 5.0) | OR | 1.29 (1.20–1.38) | <0.001 | 0.782 |
| Insulin or oral glucose-lowering medication use, % | 5.4 | 8.9 | 3.6 (2.0 to 5.2) | OR | 1.28 (1.19–1.37) | <0.001 | 0.151 |
| Undiagnosed diabetes (no diagnosis and HbA1c ≥6.5%), % | 1.7 | 2.1 | 0.4 (−0.2 to 1.0) | OR | 1.10 (1.00–1.22) | 0.060 | 0.291 |
| Extended diabetes (primary definition plus fasting glucose ≥126 mg/dL), % | 8.9 | 14.3 | 5.4 (2.4 to 8.4) | OR | 1.27 (1.17–1.37) | <0.001 | 0.174 |
| Fasting glucose ≥126 mg/dL, % | 6.9 | 10.4 | 3.4 (1.1 to 5.8) | OR | 1.28 (1.18–1.39) | <0.001 | 0.315 |
| Mean HbA1c, % | 5.43 | 5.68 | 0.25 (0.14 to 0.36) | Difference | 0.11 (0.09–0.13) | <0.001 | 0.071 |
| Mean fasting glucose, mg/dL | 100.9 | 108.0 | 7.1 (4.0 to 10.2) | Difference | 3.83 (2.98–4.68) | <0.001 | 0.413 |
| **Chronic kidney disease** |  |  |  |  |  |  |  |
| CKD (eGFR <60 mL/min/1.73 m², UACR ≥30 mg/g, weak/failing kidneys, or dialysis), % | 14.7 | 13.5 | −1.2 (−3.2 to 0.9) | OR | 0.98 (0.93–1.03) | 0.339 | 0.559 |
| eGFR <60 mL/min/1.73 m², % | 5.4 | 5.3 | −0.1 (−1.2 to 1.0) | OR | 0.91 (0.85–0.97) | 0.006 | 0.975 |
| UACR ≥30 mg/g, % | 10.7 | 9.4 | −1.3 (−2.7 to 0.1) | OR | 0.98 (0.93–1.03) | 0.476 | 0.155 |
| Self-reported weak or failing kidneys, % | 2.4 | 2.5 | 0.0 (−1.0 to 1.0) | OR | 1.19 (1.07–1.32) | 0.002 | 0.965 |
| Dialysis in past 12 months, % | — | 0.1 | −0.1 (−0.2 to 0.1) | OR | 1.10 (0.88–1.39) | 0.400 | 0.025 |
| Mean eGFR, mL/min/1.73 m² | 95.1 | 97.3 | 2.2 (1.0 to 3.5) | Difference | 1.30 (0.89–1.70) | <0.001 | 0.068 |
| **Smoking** |  |  |  |  |  |  |  |
| Current smoking, % | 24.4 | 14.9 | −9.5 (−13.2 to −5.8) | OR | 0.76 (0.71–0.81) | <0.001 | 0.158 |
| Former smoking, % | 25.0 | 22.1 | −2.9 (−5.7 to −0.1) | OR | 0.95 (0.90–0.99) | 0.019 | 0.942 |
| Never smoking, % | 50.6 | 63.0 | 12.4 (7.3 to 17.5) | OR | 1.25 (1.18–1.32) | <0.001 | 0.453 |
| Quit ratio (former/ever smokers), % | 49.3 | 58.6 | 9.2 (4.6 to 13.9) | OR | 1.18 (1.11–1.26) | <0.001 | 0.212 |
| **Alcohol** |  |  |  |  |  |  |  |
| Drinks at least monthly in past 12 months, % | 51.3 | 55.8 | 4.4 (−1.9 to 10.7) | OR | 1.11 (1.03–1.20) | 0.006 | 0.069 |
| Mean drinks per drinking day among past-year drinkers | 2.78 | 2.78 | 0.00 (−0.25 to 0.26) | Difference | −0.03 (−0.11 to 0.04) | 0.364 | 0.590 |
| Past-year drinkers exceeding >2 drinks per drinking day (men) or >1 (women), % | 49.0 | 54.4 | 5.4 (0.5 to 10.3) | OR | 1.08 (1.02–1.15) | 0.013 | 0.632 |
| **Prior cardiovascular disease** |  |  |  |  |  |  |  |
| Prior CVD (any), % | 8.7 | 8.2 | −0.5 (−2.1 to 1.1) | OR | 0.95 (0.89–1.02) | 0.141 | 0.238 |
| Heart failure, % | 2.2 | 2.5 | 0.3 (−0.4 to 1.0) | OR | 0.99 (0.90–1.09) | 0.838 | 0.581 |
| Coronary heart disease, % | 3.4 | 3.3 | −0.0 (−0.8 to 0.7) | OR | 0.97 (0.87–1.07) | 0.507 | 0.082 |
| Angina, % | 3.1 | 1.4 | −1.6 (−2.5 to −0.8) | OR | 0.72 (0.64–0.80) | <0.001 | 0.221 |
| Myocardial infarction, % | 3.6 | 2.8 | −0.8 (−1.6 to 0.0) | OR | 0.89 (0.82–0.98) | 0.017 | 0.746 |
| Stroke, % | 2.5 | 3.1 | 0.6 (−0.3 to 1.4) | OR | 1.07 (0.98–1.17) | 0.116 | 0.675 |
| **Awareness, treatment, and control among adults with each condition** |  |  |  |  |  |  |  |
| Hypertension awareness, % | 45.6 | 54.0 | 8.4 (4.0 to 12.8) | OR | 1.20 (1.13–1.26) | <0.001 | <0.001 |
| Hypertension treatment, % | 27.8 | 38.0 | 10.3 (6.5 to 14.0) | OR | 1.25 (1.18–1.32) | <0.001 | <0.001 |
| Hypertension control (BP <130/80 mm Hg), % | 14.2 | 21.5 | 7.3 (4.7 to 9.9) | OR | 1.27 (1.21–1.34) | <0.001 | <0.001 |
| Dyslipidemia awareness, % | 42.7 | 57.2 | 14.5 (10.6 to 18.3) | OR | 1.33 (1.26–1.40) | <0.001 | 0.007 |
| Dyslipidemia treatment, % | 13.8 | 28.4 | 14.6 (12.3 to 16.9) | OR | 1.36 (1.29–1.43) | <0.001 | <0.001 |
| Dyslipidemia control (fasting LDL <100 mg/dL), % | 17.0 | 34.2 | 17.3 (12.8 to 21.7) | OR | 1.44 (1.34–1.55) | <0.001 | <0.001 |
| Diabetes awareness, % | 78.2 | 81.3 | 3.0 (−6.4 to 12.5) | OR | 1.13 (1.02–1.26) | 0.018 | 0.739 |
| Diabetes treatment, % | 57.6 | 68.5 | 10.9 (0.3 to 21.5) | OR | 1.08 (0.99–1.18) | 0.082 | 0.461 |
| Diabetes control (HbA1c <7%), % | 36.4 | 41.9 | 5.6 (−8.6 to 19.8) | OR | 1.02 (0.93–1.12) | 0.704 | <0.001 |
| **PREVENT predicted risk** |  |  |  |  |  |  |  |
| 10-year total CVD risk, mean % | 5.66 | 5.28 | −0.39 (−0.74 to −0.03) | Difference | −0.10 (−0.21 to −0.00) | 0.045 | 0.551 |
| 10-year ASCVD risk, mean % | 3.66 | 3.20 | −0.45 (−0.70 to −0.21) | Difference | −0.16 (−0.22 to −0.09) | <0.001 | 0.241 |
| 10-year heart failure risk, mean % | 3.05 | 3.19 | 0.14 (−0.09 to 0.37) | Difference | 0.10 (0.03–0.17) | 0.004 | 0.588 |
| 10-year total CVD risk <5%, % | 64.5 | 65.4 | 0.9 (−2.4 to 4.2) | OR | 0.99 (0.92–1.06) | 0.722 | 0.959 |
| 10-year total CVD risk 5 to <7.5%, % | 10.3 | 10.4 | 0.1 (−2.2 to 2.4) | OR | 1.03 (0.96–1.11) | 0.398 | 0.333 |
| 10-year total CVD risk 7.5 to <20%, % | 20.1 | 20.0 | −0.2 (−1.9 to 1.6) | OR | 1.03 (0.98–1.09) | 0.267 | 0.251 |
| 10-year total CVD risk ≥20%, % | 5.0 | 4.2 | −0.8 (−2.0 to 0.4) | OR | 0.89 (0.82–0.98) | 0.013 | 0.997 |
| 10-year total CVD risk ≥7.5%, % | 25.2 | 24.2 | −1.0 (−2.9 to 0.9) | OR | 0.99 (0.92–1.06) | 0.696 | 0.305 |
| 30-year total CVD risk, mean % (ages 30–59) | 15.32 | 14.14 | −1.18 (−2.19 to −0.17) | Difference | −0.37 (−0.63 to −0.12) | 0.004 | 0.857 |
| 30-year ASCVD risk, mean % (ages 30–59) | 9.95 | 8.91 | −1.04 (−1.77 to −0.31) | Difference | −0.40 (−0.58 to −0.22) | <0.001 | 0.808 |
| 30-year heart failure risk, mean % (ages 30–59) | 7.94 | 8.36 | 0.42 (−0.21 to 1.05) | Difference | 0.31 (0.14–0.48) | <0.001 | 0.661 |

Obesity increased across all subgroups with data available throughout the study period. However, the magnitude of the increase varied by age (P for interaction = 0.006), with the largest increase in odds among adults aged 20–39 years (OR, 1.27). Trends also differed by race and Hispanic origin (P = 0.045) and education (P = 0.048). Mexican American adults had the largest increase in odds (OR, 1.42), as did adults with some college education (OR, 1.32) within their respective subgroups. No significant differences in trends were observed by sex or income **(Figure 4; eTable 4; eFigures 5–9)**.

### 3.3. Diabetes

Diabetes prevalence increased from 8.2% to 12.7% (OR, 1.26; 95% CI, 1.19–1.34). Most of this increase was attributable to diagnosed diabetes, which accounted for 4.1 of the 4.5 percentage-point increase. In contrast, undiagnosed diabetes changed little, from 1.7% to 2.1%, with no statistically significant trend (P = 0.060) **(Figure 1**; **Table 2; eFigure 4)**.

Diabetes prevalence increased significantly in most subgroups, although trends differed significantly only by race and Hispanic origin (P for interaction = 0.016). Mexican American adults had the largest increase in odds (OR, 1.33), whereas non-Hispanic Black adults had the highest prevalence in 2021–2023 (21.4%). No significant differences in trends were found by sex, age, income, or education **(Figure 4; eTable 4; eFigure 7)**.

Among adults with diabetes, awareness increased from 78.2% to 81.3% (OR, 1.13; 95% CI, 1.02–1.26). Glycemic control followed a different pattern. The proportion with HbA1c <7% initially increased from 36.4% to 51.2% in 2003–2004, before declining to 41.9% in 2021–2023 (P for nonlinearity <0.001) **(Figure 2**; **Table 2; eTable 3)**.

**Figure 2.**
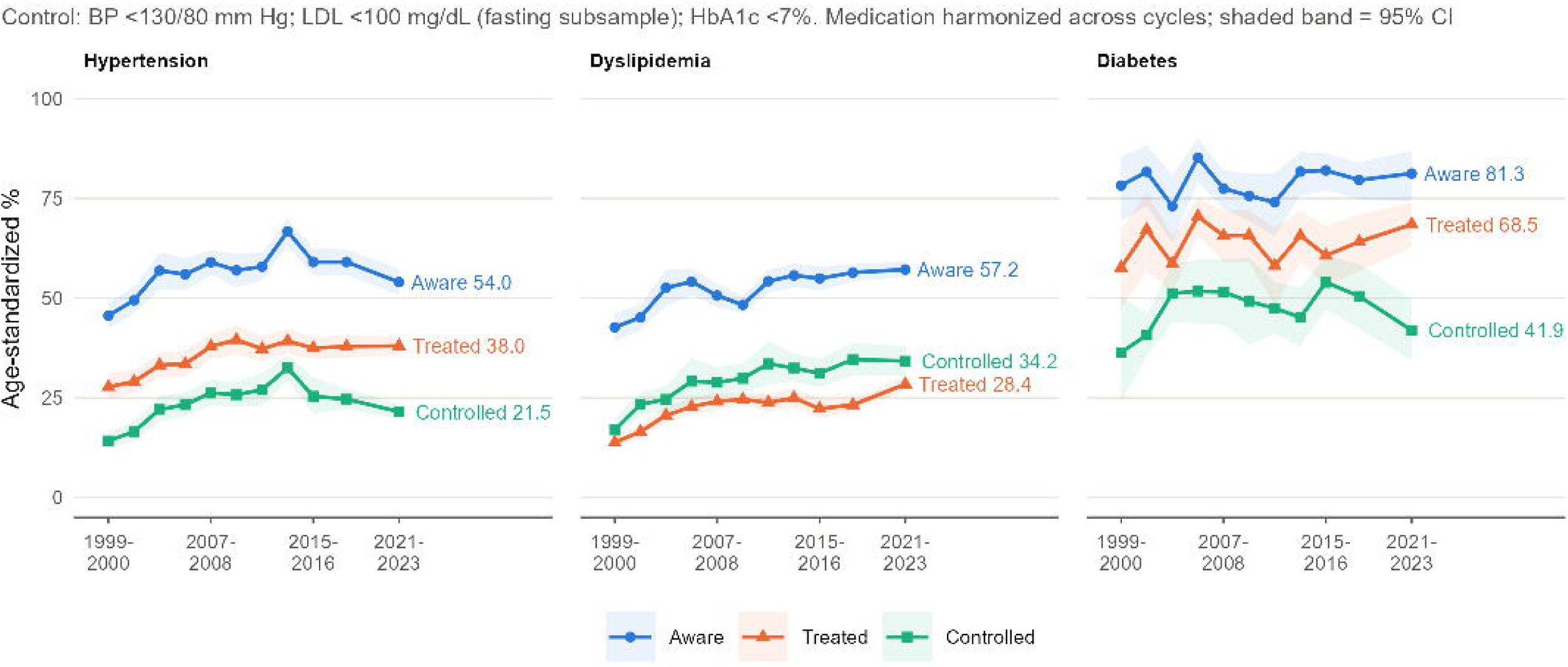
Trends in awareness, treatment, and control of hypertension, dyslipidemia, and diabetes among US adults, NHANES 1999–2000 to 2021–2023. Estimates were age-standardized to the 2000 US population and restricted to adults with each condition. Awareness was based on self-reported diagnosis and treatment on medication use. Control was defined as blood pressure below 130/80 mm Hg for hypertension, fasting low-density lipoprotein (LDL) cholesterol below 100 mg/dL for dyslipidemia, and glycated hemoglobin (HbA1c) below 7% for diabetes. Dyslipidemia control was assessed in the fasting subsample. Shaded bands indicate 95% CIs.

### 3.4. Hypertension

Hypertension prevalence showed no significant linear trend, decreasing from 50.6% to 48.1% (OR, 0.96; 95% CI, 0.92–1.02) **(Figure 1**; **Table 2)**. However, the proportion of adults with measured blood pressure ≥130/80 mm Hg declined from 44.0% to 37.2%, while antihypertensive medication use increased from 17.6% to 22.2% **(eFigure 4)**. Mean systolic blood pressure also decreased, from 124.2 to 120.2 mm Hg **(eFigure 3)**.

**Figure 3.**
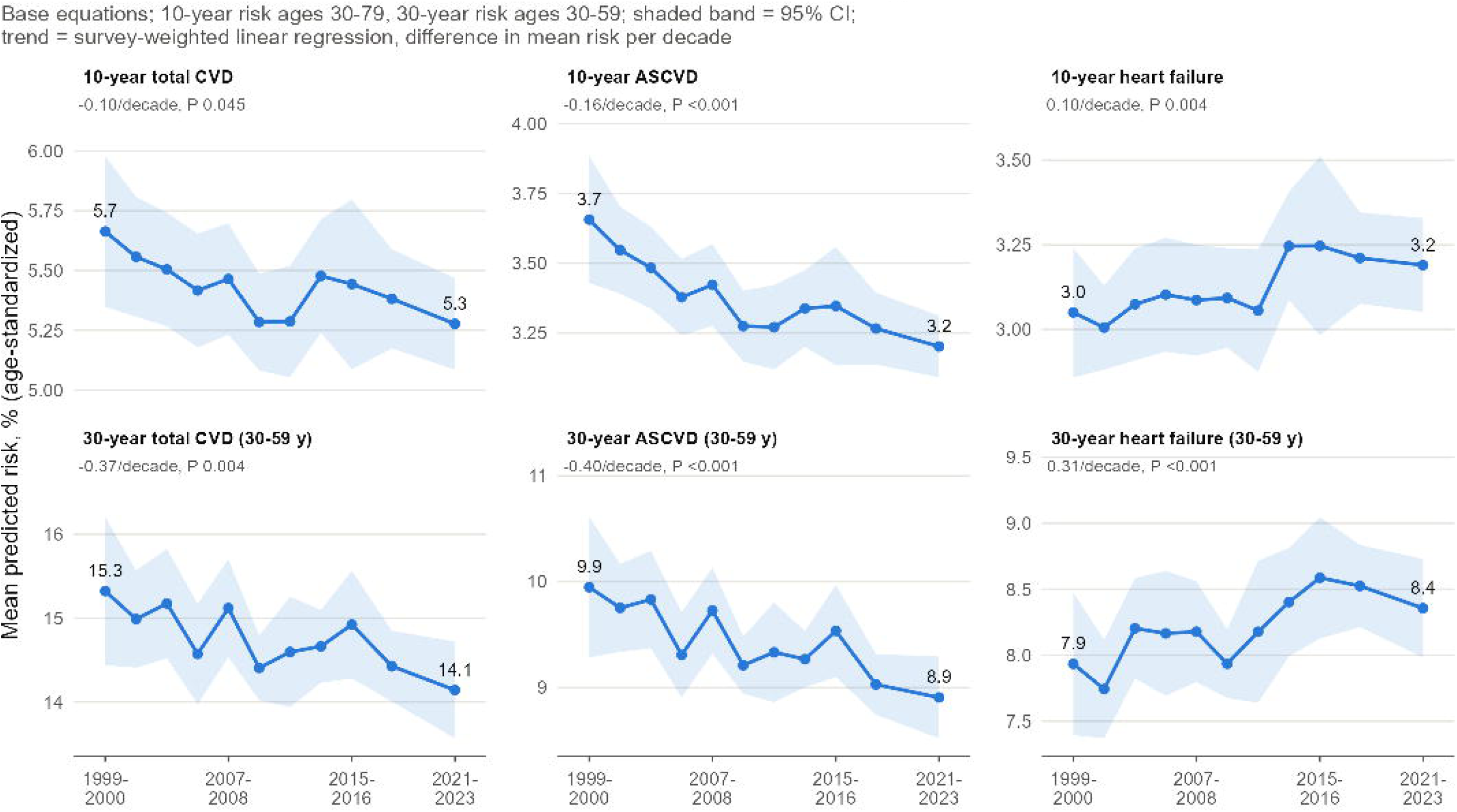
Trends in PREVENT-predicted cardiovascular risk among US adults without self-reported CVD, NHANES 1999–2000 to 2021–2023. Upper panels show mean predicted 10-year risks of total CVD, atherosclerotic CVD (ASCVD), and heart failure (HF) among adults aged 30–79 years. Lower panels show the corresponding 30-year risks among adults aged 30–59 years. Risk estimates were calculated using the base PREVENT equations and age-standardized within the eligible age ranges. Shaded bands indicate 95% CIs. Changes per decade and P values were obtained from survey-weighted linear regression adjusted for age group.

Hypertension trends varied by age (P for interaction <0.001) and education (P = 0.005). The largest decline in odds occurred among adults aged ≥60 years (OR, 0.85), whose prevalence fell from 80.2% to 73.0%. By education, odds declined most among college graduates (OR, 0.91) but increased among adults with some college education or an associate degree (OR, 1.07). No significant differences in trends were observed by sex, race and Hispanic origin, or income **(Figure 4; eTable 4; eFigures 6 and 9).**

**Figure 4.**
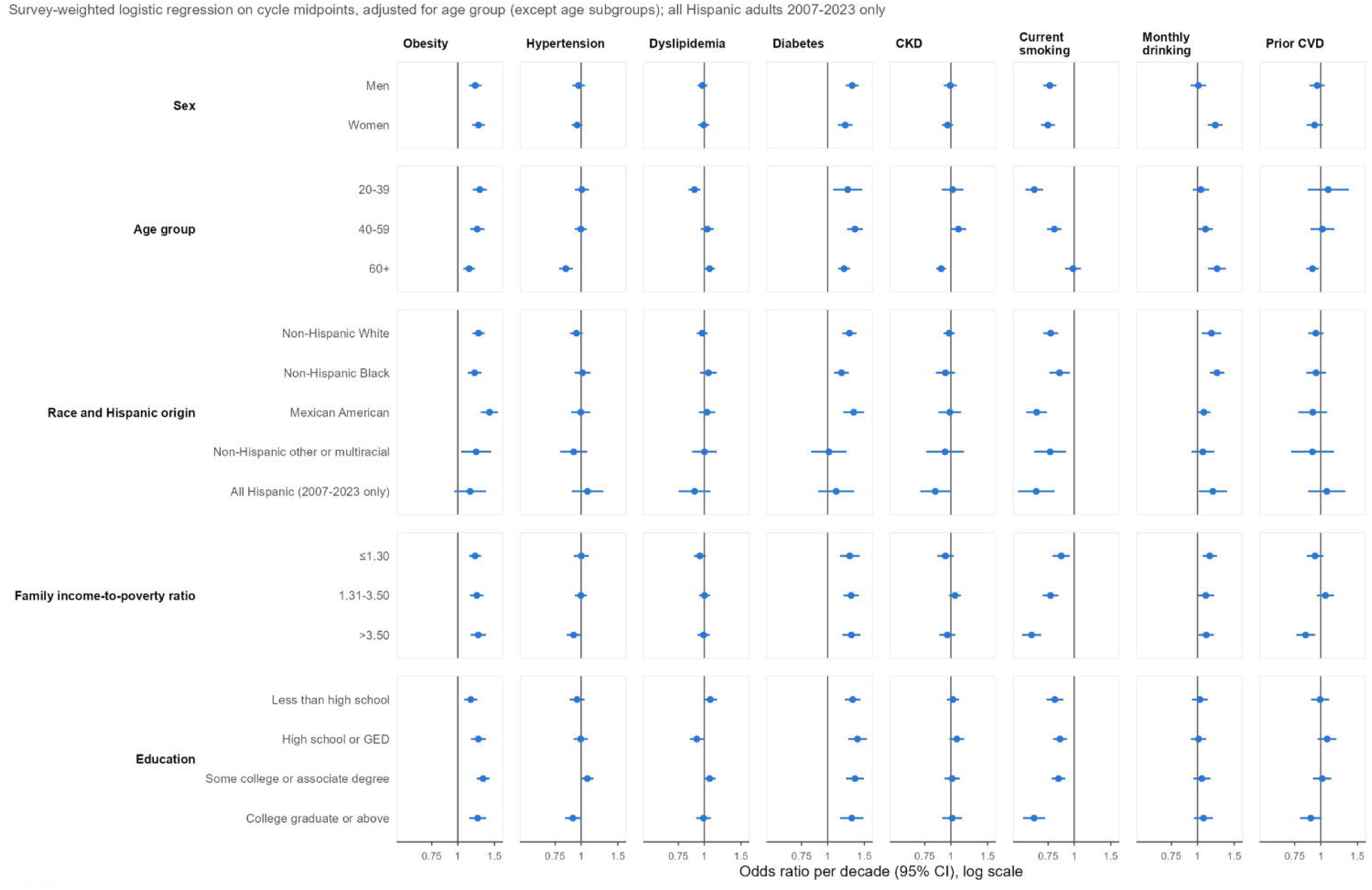
Subgroup trends in cardiovascular risk factors among US adults, NHANES 1999–2000 to 2021–2023. Points and horizontal lines represent ORs per decade and 95% CIs for obesity, hypertension, dyslipidemia, diabetes, CKD, current smoking, monthly alcohol consumption, and prior CVD. Estimates were obtained from survey-weighted logistic regression adjusted for age group, except in age-stratified analyses, which present age-specific estimates. The vertical reference line indicates an OR of 1.0. Analyses of all Hispanic adults were restricted to 2007–2008 through 2021–2023. P values for trends and interactions are reported in eTable 4.

Among adults with hypertension, blood pressure control below 130/80 mm Hg initially improved from 14.2% to 32.5% in 2013–2014, before declining to 21.5% in 2021–2023. After 2013–2014, control decreased by an estimated 1.17 percentage points annually (95% CI, 0.70–1.64).

Awareness followed a similar pattern, increasing from 45.6% to 66.8% before falling to 54.0%, whereas treatment remained relatively stable at approximately 38% after 2009–2010 **(Figure 2**; **Table 2; eTable 6)**. When hypertension was defined using the higher threshold of 140/90 mm Hg, prevalence was 34.3% in both the first and last survey cycles **(eTable 5; eFigure 10)**.

### 3.5. Dyslipidemia

Overall dyslipidemia prevalence showed no significant trend, declining from 57.5% to 53.4% (OR, 0.98; 95% CI, 0.94–1.03; P = 0.51) **(Figure 1**; **Table 2)**. However, individual lipid measures improved over the study period. The prevalence of high total cholesterol (≥240 mg/dL) fell from 17.8% to 11.0%, and low HDL cholesterol from 39.3% to 26.7%, while lipid-lowering medication use doubled from 8.8% to 17.8% **(eFigure 4)**. Mean total cholesterol decreased from 203.6 to 188.1 mg/dL, and mean fasting LDL cholesterol from 126.0 to 110.5 mg/dL **(eFigure 3)**.

Among adults with dyslipidemia, treatment increased from 13.8% to 28.4%. LDL cholesterol control below 100 mg/dL also improved, from 17.0% to 34.2% in the fasting subsample, with most of the improvement occurring by 2005–2006, with slower gains thereafter **(Figure 2**; **Table 2)**. When low HDL cholesterol was excluded from the definition, dyslipidemia prevalence instead increased from 33.6% to 38.2% (OR, 1.10; 95% CI, 1.06–1.14) **(eTable 5; eFigures 10–11)**.

Dyslipidemia trends differed significantly by age (P for interaction <0.001) and education (P = 0.011). Odds declined most among adults aged 20–39 years (OR, 0.90) but showed a borderline increase among those aged ≥60 years (OR, 1.06; P = 0.050). Across education groups, adults with a high school diploma or GED had the largest decline in odds (OR, 0.92). No significant differences in trends were found by sex, race and Hispanic origin, or income **(Figure 4; eTable 4; eFigures 6 and 9)**.

### 3.6. Smoking and alcohol use

Current smoking declined from 24.4% to 14.9% (OR, 0.76; 95% CI, 0.71–0.81), representing the largest absolute decline among the primary outcomes **(Figure 1**; **Table 2)**. Significant declines were observed across all subgroups except adults aged ≥60 years. However, the magnitude of the decline varied by age (P for interaction <0.001), race and Hispanic origin (P = 0.011), income (P <0.001), and education (P <0.001). Adults aged 20–39 years had the largest decline in odds (OR, 0.64), whereas smoking remained relatively unchanged among those aged ≥60 years (OR, 0.99; P = 0.773). Among racial and ethnic groups, Mexican American adults had the largest decline (OR, 0.66). Declines were also greatest among adults in the highest-income group (OR, 0.62) and college graduates (OR, 0.64). Trends did not differ significantly by sex **(Figure 4; eTable 4; eFigures 5–9)**.

Monthly drinking increased from 51.3% to 55.8% (OR, 1.11; 95% CI, 1.03–1.20), although the difference between the first and last cycles was not statistically significant (4.4 percentage points; 95% CI, −1.9 to 10.7) **(Figure 1**; **Table 2)**. Most of the increase occurred through 2011–2012, with no significant change thereafter **(eTable 6; eFigure 13)**. Trends varied by sex (P for interaction <0.001), age (P <0.001), and race and Hispanic origin (P = 0.017). Monthly drinking increased significantly among women (OR, 1.22) but not men (OR, 1.01). Adults aged ≥60 years had the largest increase in odds among age groups (OR, 1.24), while non-Hispanic Black adults had the largest increase among racial and ethnic groups (OR, 1.24). No significant differences in trends were observed by income or education **(Figure 4; eTable 4; eFigures 5–9)**.

Among past-year drinkers, the proportion consuming more than 2 drinks per drinking day for men or more than 1 for women increased from 49.0% to 54.4% (OR, 1.08; 95% CI, 1.02–1.15). However, mean consumption remained unchanged at 2.78 drinks per drinking day in both the first and last survey cycles **(Table 2; eFigure 15)**.

### 3.7. Chronic kidney disease and prior cardiovascular disease

Overall CKD prevalence showed no significant trend, declining from 14.7% to 13.5% **(Figure 1**; **Table 2)**. However, eGFR-defined CKD declined significantly (OR, 0.91; 95% CI, 0.85–0.97), while self-reported weak or failing kidneys increased (OR, 1.19; 95% CI, 1.07–1.32) **(eTable 3; eFigure 4)**. CKD trends differed by age (P for interaction <0.001), with a significant decline observed only among adults aged ≥60 years (OR, 0.90). Trends in younger age groups were not significant. No significant differences in trends were observed by sex, race and Hispanic origin, income, or education **(Figure 4; eTable 4; eFigure 6)**.

Prior CVD prevalence also remained relatively stable, decreasing from 8.7% to 8.2% **(Figure 1**; **Table 2)**. Trends differed significantly by income (P for interaction = 0.005), with a significant decline observed only in the highest-income group (OR, 0.85). Prior CVD also declined among adults aged ≥60 years (OR, 0.91; P = 0.009), although trends did not differ significantly across age groups (P for interaction = 0.148). No significant differences in trends were found by sex, race and Hispanic origin, or education **(Figure 4; eTable 4; eFigure 8)**. Among individual CVD conditions, angina declined from 3.1% to 1.4% (OR, 0.72; P <0.001). Myocardial infarction also showed a significant declining trend (OR, 0.89; P = 0.017), although the difference between the first and last cycles was borderline significant (3.6% to 2.8%; P for change = 0.051). No significant linear trends were observed for heart failure, coronary heart disease, or stroke **(Table 2; eTable 3; eFigure 15)**.

### 3.8. Predicted cardiovascular risk

Among 35,782 adults without reported CVD, mean predicted 10-year total CVD risk declined from 5.66% to 5.28% (−0.10 percentage points per decade; P = 0.045). Predicted 10-year ASCVD risk also decreased, from 3.66% to 3.20%, whereas predicted heart failure risk increased from 3.05% to 3.19%. By 2021–2023, predicted heart failure risk was nearly equal to predicted ASCVD risk **(Figure 3**; **Table 2)**.

A similar pattern emerged for 30-year risk estimates among adults aged 30–59 years. Predicted ASCVD risk declined from 9.95% to 8.91% (−0.40 percentage points per decade; P <0.001), while predicted heart failure risk increased from 7.94% to 8.36% (0.31 percentage points per decade; P <0.001) **(Figure 3**; **Table 2)**. The proportion of adults with predicted 10-year total CVD risk ≥20% also declined significantly in the trend model (OR per decade, 0.89; 95% CI, 0.82–0.98), although the difference between the first and last cycles was not statistically significant (5.0% to 4.2%; P for change = 0.181) **(Table 2; eTable 3; eFigure 14)**.

## 4. DISCUSSION

In this nationally representative analysis of US adults spanning 11 NHANES survey periods from 1999–2000 to 2021–2023, trends in cardiovascular risk factors varied considerably.

Obesity, severe obesity, and diabetes became more prevalent, while current smoking and several lipid measures declined. Although measured blood pressure and cholesterol levels decreased and medication use increased, the overall prevalence of hypertension and dyslipidemia remained largely unchanged. Hypertension control improved until 2013–2014 but declined thereafter. Meanwhile, mean predicted 10-year atherosclerotic cardiovascular disease (ASCVD) risk decreased modestly, whereas predicted heart failure (HF) risk increased.

These findings build on earlier NHANES studies documenting increases in obesity and diabetes alongside declines in smoking and cholesterol ^9,10,40–43^ . Our obesity estimates for 2021–2023 were also consistent with those reported by the National Center for Health Statistics ^12,13^ . A recent study examining PREVENT risk trends through 2017–2018 found that age-standardized total cardiovascular disease (CVD) risk remained stable, while ASCVD risk declined and heart failure (HF) risk increased ^19^. We extend these findings through 2021–2023, incorporate 30-year risk estimates, and assess changes in risk factors and their management within the same analysis. Another study, using a hypertension threshold of 140/90 mm Hg, reported a decline in blood pressure control through 2017–2020, with no significant change between 2017–2020 and 2021–2023 ^44^. Differences in hypertension definitions and analytic methods may help explain variations between these findings and our trend estimates.

Examining individual components helps explain why overall prevalence remained stable despite improvements in some measures. Hypertension prevalence changed little despite declining blood pressure and increasing medication use. Similarly, dyslipidemia prevalence remained stable, although high total cholesterol and low HDL cholesterol declined and treatment increased.

Excluding low HDL cholesterol changed the dyslipidemia trend, highlighting the influence of its definition. Rising diabetes prevalence was driven mainly by diagnosed cases, while undiagnosed diabetes changed little. Glycemic control also declined after an early peak. These findings underscore the continued need for prevention, detection, and sustained management.

Subgroup analyses revealed important differences in risk-factor trends. Obesity increased across all subgroups, with larger increases in odds among younger adults, Mexican American adults, and those with some college education. Diabetes trends differed by race and Hispanic origin.

Smoking declined overall but changed little among adults aged 60 years or older, with varying declines across racial, income, and education groups. Hypertension trends differed by age and education, while monthly drinking increased among women and older adults, with differences by race and Hispanic origin. These findings show that overall improvements can mask important differences across population groups.

A recent analysis of national mortality records also documented a nearly fourfold increase in metabolic multimorbidity among US decedents aged 25–44 years between 1999 and 2024, with higher prevalence among female and Black decedents ^45^. Although based on death certificate data rather than population prevalence estimates, these findings provide complementary evidence of a growing burden of coexisting metabolic conditions among younger adults.

The diverging trends in predicted ASCVD and HF risk highlight the need for prevention strategies that address obesity and diabetes alongside traditional atherosclerotic risk factors. However, PREVENT estimates reflect predicted risk rather than observed events and should be interpreted accordingly ^15,19,46–48^.

Strengths of this study include nationally representative data spanning 11 survey periods, appropriate survey-design methods, harmonized definitions, assessment of composite outcomes and their components, and subgroup and sensitivity analyses. Several limitations warrant consideration. The repeated cross-sectional design cannot establish individual trajectories or causality. Some outcomes relied on self-reported diagnoses and behaviors, while single-occasion kidney measurements could not confirm persistent chronic kidney disease. Changes in laboratory assays, measurement devices, questionnaires, and the pandemic-related interruption may have affected comparisons despite harmonization and sensitivity analyses. PREVENT estimates were also limited by equation inputs and eligible age ranges and do not represent observed cardiovascular events.

In conclusion, declines in smoking, cholesterol, and measured blood pressure were accompanied by rising obesity and diabetes, worsening hypertension control after its peak, and increasing predicted HF risk. These findings underscore the need for sustained cardiovascular prevention and management that addresses subgroup differences and both ASCVD and HF risk.

## Acknowledgments

None.

## Ethics Approval and Informed Consent

This study used publicly available, deidentified data from the National Health and Nutrition Examination Survey (NHANES). NHANES protocols were approved by the National Center for Health Statistics Research Ethics Review Board, and participants provided informed consent. This secondary analysis did not require additional institutional review board approval.

## Data Availability

All data used in this study are publicly available through the National Health and Nutrition Examination Survey (NHANES), conducted by the National Center for Health Statistics, Centers for Disease Control and Prevention (https://www.cdc.gov/nchs/nhanes/). No new datasets were generated for this study.

## Author Contributions

**Hafiz M Ahmed**

Conceptualization, Methodology, Project administration, Supervision, Investigation, Data curation, Formal analysis, Software, Visualization, Writing – original draft, Writing –review & editing.

**Nawal Younas**

Investigation, Data curation, Visualization, Writing – original draft, Writing – review & editing.

**Muhammad Owais**

Investigation, Data curation, Validation, Writing – original draft, Writing – review & editing.

## Abbreviations

AHA: American Heart Association
ASCVD: atherosclerotic cardiovascular disease
BMI: body mass index
BP: blood pressure
CI: confidence interval
CKD: chronic kidney disease
COVID-19: coronavirus disease 2019
CVD: cardiovascular disease
eGFR: estimated glomerular filtration rate
GED: General Educational Development
HbA1c: glycated hemoglobin
HDL: high-density lipoprotein
HF: heart failure
LDL: low-density lipoprotein
MEC: mobile examination center
NCHS: National Center for Health Statistics
NHANES: National Health and Nutrition Examination Survey
OR: odds ratio
PREVENT: Predicting Risk of Cardiovascular Disease Events
RECORD: Reporting of studies Conducted using Observational Routinely-collected health Data
STROBE: Strengthening the Reporting of Observational Studies in Epidemiology
US: United States.

