## Supplementary Material for "Trends in Cardiovascular Risk Factors, Disease Management, and Predicted Cardiovascular Risk Among US Adults, 1999–2023"

Supplementary Online Content

**Supplementary Methods and Selected Results for Trends in Cardiovascular Risk Factors Among US Adults, 1999–2023**

This supplement provides additional methods and selected results for the NHANES analysis.

**Contents**

1. eMethods. Additional details on study population, outcome definitions, and statistical analysis
2. eTable 1. Analytic sample and PREVENT eligibility by survey cycle
3. eTable 2. Unweighted sample sizes for each outcome, by cycle
4. eTable 3. Endpoint estimates and trends across the study period
5. eTable 4. Subgroup trends and tests for interaction
6. eTable 5. Sensitivity analyses
7. eTable 6. Joinpoint regression of age-standardized estimates
8. eTable 7. Obesity and overweight among non-Hispanic Asian adults using standard and Asian BMI thresholds, 2011–2023
9. eTable 8. Item missingness among eligible adults, by cycle
10. eTable 9. Nonresponse analyses for items with more than 10% missing data
1. eFigure 1. Selection of the analytic sample
2. eFigure 2. Weight status by BMI class
3. eFigure 3. Mean levels of continuous risk factors
4. eFigure 4. Components of the composite risk-factor definitions
5. eFigure 5. Primary risk factors by sex
6. eFigure 6. Primary risk factors by age group (crude within age group)
7. eFigure 7. Primary risk factors by race and Hispanic origin
8. eFigure 8. Primary risk factors by family income-to-poverty ratio
9. eFigure 9. Primary risk factors by education
10. eFigure 10. Sensitivity analyses: odds ratios per decade for primary and alternative analyses
11. eFigure 11. HDL cholesterol assay changes and the dyslipidemia trend
12. eFigure 12. Effect of harmonizing medication questions across cycles
13. eFigure 13. Joinpoint regression of the primary outcomes
14. eFigure 14. Distribution of 10-year PREVENT total CVD risk
15. eFigure 15. Smoking, alcohol use, and prior CVD components
16. eFigure 16. Item missingness among eligible adults, by cycle
17. eFigure 17. Primary and extended fasting definitions of dyslipidemia and diabetes

**eMethods. Additional Details on Study Population, Outcome Definitions, and Statistical Analysis**

**1. Survey Design, Eligibility, and Sample Selection**

The National Health and Nutrition Examination Survey (NHANES) uses a stratified, multistage probability sampling design to obtain nationally representative estimates of the civilian, noninstitutionalized US population. Participants complete household interviews and undergo standardized examinations at mobile examination centers (MECs). Survey periods were treated as repeated cross-sectional samples rather than longitudinal follow-up.

The analysis included nonpregnant adults aged 20 years or older who completed an MEC examination. For each outcome, analyses were restricted to participants with the necessary measurements or questionnaire data, resulting in outcome-specific sample sizes. Fasting measurements were analyzed only among eligible participants in the fasting subsample. No separate prospective sample-size calculation was performed. Sample selection and eligibility for the PREVENT risk equations across survey periods are summarized in **eTable 1**.

Eligibility restrictions were applied through survey domain analysis to preserve the survey design structure during variance estimation.

**2. Survey Weights and Variance Estimation**

For each outcome, we used the survey weight corresponding to the smallest subsample containing all required variables. Weights were selected according to National Center for Health Statistics (NCHS) recommendations for the 1999–2010, 2011–2016, 2017–March 2020, and August 2021–August 2023 survey periods.

| **Analysis** | **1999–2002** | **2003–2016** | **2017–Mar 2020** | **2021–2023** |
| --- | --- | --- | --- | --- |
| Interview and examination outcomes | 2 × WTMEC4YR | WTMEC2YR | WTMECPRP | WTMEC2YR |
| Blood tests (TC, HDL, HbA1c, creatinine, UACR); PREVENT | 2 × WTMEC4YR | WTMEC2YR | WTMECPRP | WTPH2YR (phlebotomy) |
| Fasting LDL, triglycerides, glucose | 2 × WTSAF4YR | WTSAF2YR | WTSAFPRP | WTSAF2YR |

For 1999–2002, the NCHS-recommended 4-year weights were multiplied by 2 to make them comparable with subsequent 2-year weights, given differences in the census populations used to construct the original weights. For pooled temporal analyses, cycle-specific weights were then scaled according to each cycle's share of the 23.2-year study period. The 2017–March 2020 cycle was assigned 3.2 years, while all other cycles were assigned 2 years. Time was modeled using survey-cycle midpoints (2000, 2002, 2004, 2006, 2008, 2010, 2012, 2014, 2016, 2018.6, and 2022.7) to account for unequal intervals between cycles, including the interruption in data collection from April 2020 through July 2021.

Variances were estimated using Taylor series linearization, accounting for masked variance strata (SDMVSTRA) and primary sampling units (SDMVPSU). Primary sampling units were nested within strata, and strata containing a single primary sampling unit were centered at the grand mean. Separate survey designs were constructed for each cycle. Domain-specific survey degrees of freedom were calculated as the number of primary sampling units minus the number of strata represented within each analytic domain.

**3. Outcome Definitions and Harmonization**

Definitions of the primary outcomes are provided in the main Methods section. Additional outcome definitions, component measures, and analytic considerations are described below.

Composite outcomes (obesity, hypertension, dyslipidemia, diabetes, and chronic kidney disease [CKD]) were considered present if any component criterion was met. Outcomes were classified as absent if at least one component was available and all available components were negative, and as missing if no component data were available. Responses of "refused" or "don't know" were treated as missing.

| **Outcome** | **Definition** | **Sample and weight** |
| --- | --- | --- |
| Weight status | Mutually exclusive BMI categories (kg/m²): underweight, <18.5; normal weight, 18.5–24.9; overweight, 25.0–29.9; obesity class I, 30.0–34.9; class II, 35.0–39.9; class III, ≥40.0 | MEC |
| Obesity | BMI ≥30 kg/m² or waist circumference >102 cm in men or >88 cm in women | MEC |
| Hypertension | Mean BP ≥130/80 mm Hg, current antihypertensive medication use, or self-reported diagnosis | MEC |
| Dyslipidemia (primary definition) | Total cholesterol ≥240 mg/dL, HDL cholesterol <40 mg/dL in men or <50 mg/dL in women, lipid-lowering medication use, or self-reported high cholesterol | Blood test |
| Dyslipidemia (extended definition) | Primary criteria plus fasting LDL cholesterol ≥160 mg/dL or triglycerides ≥150 mg/dL | Fasting |
| Diabetes (primary definition) | Self-reported diagnosis, or HbA1c ≥6.5%, or insulin or oral glucose-lowering medication | Blood test |
| Diabetes (extended definition) | Primary criteria plus fasting glucose ≥126 mg/dL | Fasting |
| Undiagnosed diabetes | HbA1c ≥6.5% without a self-reported diabetes diagnosis | Blood test |
| Chronic kidney disease | eGFR <60 mL/min/1.73 m² (2021 race-free CKD-EPI equation), urine albumin-to-creatinine ratio ≥30 mg/g, self-reported weak or failing kidneys, or dialysis in the past 12 months (not assessed in 1999–2000) | Blood test |
| Smoking | Current: ≥100 lifetime cigarettes and currently smoking every day or some days; former: ≥100 lifetime cigarettes but not currently smoking; never: <100 lifetime cigarettes. Quit ratio: former smokers divided by ever smokers | MEC |
| Alcohol | Drinking at least monthly in the past year; mean drinks per drinking day among past-year drinkers; consumption exceeding 2 drinks/day in men or 1 drink/day in women, based on drinking days | MEC |
| Prior CVD | Self-reported heart failure, coronary heart disease, angina, myocardial infarction, or stroke | MEC |
| Disease awareness | Self-reported diagnosis among participants meeting the corresponding primary disease definition | Disease-specific |
| Disease treatment | Current medication use among participants meeting the corresponding primary disease definition | Disease-specific |
| Disease control | BP <130/80 mm Hg, fasting LDL cholesterol <100 mg/dL, or HbA1c <7%, as applicable | Disease-specific |
| Continuous measures | Mean BMI, waist circumference, systolic and diastolic BP, total, HDL and non-HDL cholesterol, HbA1c, and eGFR; mean fasting LDL cholesterol, triglycerides, and glucose; geometric mean fasting triglycerides | As the measure |

Blood pressure was calculated as the mean of up to four auscultatory readings through 2015–2016 and up to three oscillometric readings from 2017 onward. For non-Hispanic Asian adults, we performed a sensitivity analysis using Asian-specific BMI thresholds of 23.0–27.4 kg/m² for overweight and ≥27.5 kg/m² for obesity, restricted to the 2011–2023 survey periods.

**4. Predicted cardiovascular risk**

We used the base PREVENT equations to estimate 10-year risks of total cardiovascular disease (CVD), atherosclerotic cardiovascular disease (ASCVD), and heart failure among adults aged 30–79 years without self-reported CVD and with complete equation inputs. Thirty-year risks were estimated among adults aged 30–59 years.

The equations included age, sex, systolic blood pressure, antihypertensive and lipid-lowering medication use, total and HDL cholesterol, diabetes, current smoking, estimated glomerular filtration rate (eGFR), and BMI. Lipid-lowering medication use served as a proxy for statin treatment.

To avoid extrapolation beyond the equations' specified ranges, continuous input values outside the permitted limits were capped at the corresponding boundaries, as detailed below.

| **Input** | **Lower limit** | **Upper limit** |
| --- | --- | --- |
| Systolic BP, mm Hg | 90 | 180 |
| Total cholesterol, mg/dL | 130 | 320 |
| HDL cholesterol, mg/dL | 20 | 100 |
| BMI, kg/m² | 18.5 | 39.9 |
| eGFR, mL/min/1.73 m² | 15 | 140 |

**5. Measurement changes across survey cycles**

Measurement protocols and questionnaire items changed over the study period. We addressed these changes as outlined below.

| **Item** | **Change** | **How we handled it** |
| --- | --- | --- |
| HDL cholesterol | HDL cholesterol was measured primarily by heparin–manganese precipitation in 1999–2002 and by direct immunoassay from 2003 onward. Additional analyzer and laboratory changes occurred in 2005–2006 and 2007–2008, respectively. NCHS cautions that these changes may affect trends from 1999 to 2008. Age-standardized mean HDL increased from 50.3 to 54.2 mg/dL by 2005–2006 before declining to 51.8 mg/dL in 2007–2008. NCHS corrected selected released values but did not provide a cross-cycle conversion equation. | We used the released HDL values without further adjustment. In sensitivity analyses, we excluded low HDL cholesterol from the dyslipidemia definition and restricted HDL-dependent outcomes to 2007–2023. |
| Serum creatinine | Creatinine measurements were not standardized to isotope-dilution mass spectrometry (IDMS) before 2007. | We applied NCHS-recommended corrections: 1.013 × creatinine + 0.147 for 1999–2000 and 0.978 × creatinine − 0.016 for 2005–2006. Values from 2001–2004 and 2007 onward were used without adjustment. |
| Blood pressure | Blood pressure was measured by mercury auscultation (up to four readings) through 2015–2016 and by oscillometric devices (up to three readings) beginning in 2017. | Because no NCHS conversion equation was available, we repeated hypertension and mean systolic blood pressure analyses using only data from 1999–2016. |
| Glucose-lowering medication | During 1999–2004, oral medication use was assessed only among adults with diagnosed diabetes. From 2005 onward, questions also included adults with prediabetes or increased diabetes risk. Beginning in 2017, insulin use was assessed only among adults with diagnosed diabetes. | To maintain a consistent definition, oral medication and insulin use were counted only among adults reporting diagnosed diabetes. Sensitivity analyses used medication responses as originally collected. |
| Lipid-lowering medication | During 1999–2010, medication use was assessed only among adults reporting a diagnosis of high cholesterol. Eligibility expanded during 2011–2020, and all participants were asked about medication use in 2021–2023. | We counted medication use only among adults reporting a high-cholesterol diagnosis. Sensitivity analyses removed this restriction. |
| Alcohol | Through 2016, drinking frequency was reported as an open-ended number of occasions per week, month, or year. Fixed response categories were introduced in 2017. | Earlier responses were converted to drinking days per year and mapped to the categories used from 2017 onward. |
| Kidney questions | In 1999–2000, participants were asked about weak or failing kidneys, but dialysis was not assessed. Both questions were available from 2001 onward. | Dialysis status was treated as missing in 1999–2000 and was not required to establish CKD completeness for that cycle. |
| Race and Hispanic origin | Mexican American participants were oversampled during 1999–2006, followed by broader Hispanic oversampling beginning in 2007. Non-Hispanic Asian participants were separately identified from 2011 onward. Race- and income-based oversampling was not used in 2021–2023. | We used Mexican American participants for trends spanning 1999–2023 and restricted analyses of all Hispanic adults to 2007–2023, consistent with NCHS recommendations. |

**6. Statistical Analysis**

***Age Standardization***

For each survey period, prevalence and mean estimates were directly age-standardized to the 2000 US standard population using age groups of 20–39, 40–59, and ≥60 years, with corresponding weights of 0.3966, 0.3718, and 0.2316. Estimates stratified by age were presented without further standardization.

PREVENT risk estimates were age-standardized within the eligible age ranges for each equation. For 10-year risk (ages 30–79 years), we used age groups of 30–39, 40–59, and 60–79 years, with weights of 0.2766, 0.4831, and 0.2402, respectively. For 30-year risk (ages 30–59 years), the corresponding age groups were 30–39, 40–49, and 50–59 years, with weights of 0.3641, 0.3693, and 0.2666.

We also calculated crude estimates of the primary outcomes for comparison with published National Center for Health Statistics (NCHS) estimates. Key estimates were independently checked against the corresponding analytic outputs and published NCHS benchmarks.

***Confidence Intervals and Estimate Reliability***

For proportions, 95% confidence intervals (CIs) were calculated using the Korn–Graubard method, which adapts the Clopper–Pearson interval for complex survey data using effective sample sizes and domain-specific degrees of freedom.

Following NCHS data presentation standards, proportion estimates were flagged as unreliable if they met any of the following criteria:

- Unweighted or effective sample size <30.
- Absolute 95% CI width ≥30 percentage points.
- Absolute 95% CI width >5 percentage points and relative CI width >130%.

Estimates with fewer than 8 degrees of freedom were also flagged for review.

For means, 95% CIs were calculated using the t distribution with domain-specific degrees of freedom. Mean estimates with relative standard errors ≥30% were flagged. Triglyceride geometric means were estimated by averaging log-transformed values and back-transforming the results.

***Temporal Trend Models***

We assessed temporal trends using pooled individual-level survey-weighted regression models, with survey-cycle midpoint expressed in decades and centered at 2011. All models adjusted for age group.

Binary outcomes were analyzed using logistic regression, with trends reported as odds ratios per decade. Continuous outcomes were analyzed using linear regression and expressed as mean differences per decade. For geometric mean outcomes, we used log-linear regression and reported ratios per decade.

To assess nonlinearity, we added a quadratic time term to the models. Absolute changes were calculated by subtracting the 1999–2000 age-standardized estimate from the corresponding 2021–2023 estimate. Changes in prevalence were expressed in percentage points, while changes in continuous outcomes were reported in their original measurement units.

Differences between the first and last survey-cycle estimates were tested using a t statistic, with degrees of freedom equal to the sum of the two independent cycle-specific degrees of freedom.

***Joinpoint Regression***

We used inverse-variance-weighted segmented regression to identify changes in temporal trends across the 11 age-standardized survey-period estimates. Weights were defined as 1/SE², following the National Cancer Institute Joinpoint methodology used in NCHS reports.

Models allowed a maximum of one joinpoint, consistent with recommendations for analyses containing 7–11 time points. Candidate joinpoints were limited to observed survey-cycle midpoints, with at least three observations required in each segment.

A one-joinpoint model was selected over a single linear trend only if a permutation test based on 4,999 standardized-residual permutations yielded P<.05. Otherwise, a single linear trend was retained for the full study period.

For each retained segment, we reported the annual slope and its 95% CI. Slopes represented percentage-point changes per year for prevalence outcomes and changes in the original measurement units per year for continuous outcomes.

***Subgroup Analyses***

We examined trends by sex, age, race and Hispanic origin, family income-to-poverty ratio, and education. Race and income categories followed NCHS recommendations, with subgroup definitions summarized below.

| **Subgroup** | **Categories** |
| --- | --- |
| Sex | Men; women |
| Age | 20–39, 40–59, ≥60 years (the NCHS standard-population groups) |
| Race and Hispanic origin | Non-Hispanic White; non-Hispanic Black; Mexican American; non-Hispanic other or multiracial. All Hispanic adults were analyzed separately for 2007–2023 only. |
| Family income-to-poverty ratio | ≤1.30; 1.31–3.50; >3.50 (cutoffs corresponding to SNAP eligibility categories in the 1999–2010 NCHS guidelines) |
| Education | Less than high school (grade 11 or below, or grade 12 without a diploma); high school graduate or GED; some college or associate degree; college graduate or higher |

Subgroup estimates were age-standardized, except for age-stratified estimates, which were presented without further standardization. Differences in temporal trends across subgroups were assessed using time-by-subgroup interaction terms in pooled survey-weighted logistic regression models adjusted for age group. Interaction P values were obtained from design-based Wald tests.

Participants with missing family income-to-poverty ratio (approximately 7%–14% per cycle) or education (<0.3%) were excluded only from analyses involving the corresponding subgroup.

***Sensitivity Analyses***

We performed sensitivity analyses to assess the potential influence of changes in measurement methods, outcome definitions, and survey design. For each alternative definition or restricted study period, we recalculated age-standardized estimates, absolute changes, and temporal trends.

| **Threat** | **Sensitivity analysis** |
| --- | --- |
| Hypertension threshold | Repeated analyses using a hypertension threshold of ≥140/90 mm Hg. |
| BP measurement change (2017) | Restricted hypertension and mean systolic blood pressure analyses to 1999–2016. |
| HDL assay changes | Excluded low HDL cholesterol from the dyslipidemia definition and restricted analyses of dyslipidemia, low HDL cholesterol, mean HDL cholesterol, and mean non-HDL cholesterol to 2007–2023. |
| Medication questionnaire changes | Repeated diabetes and dyslipidemia analyses using medication responses as originally collected, without harmonization. |
| Incomplete composite criteria | Repeated obesity, hypertension, dyslipidemia, diabetes, and CKD analyses using complete cases with all component criteria available. |
| BMI thresholds in Asian adults | Applied BMI thresholds of 23.0–27.4 kg/m² for overweight and ≥27.5 kg/m² for obesity among non-Hispanic Asian adults during 2011–2023 and compared results with those obtained using standard thresholds over the same period. |
| Pandemic interruption and 2021–2023 survey design | Restricted analyses of the eight primary outcomes to pre-pandemic survey cycles (1999–March 2020). |
| Fasting criteria | Applied expanded dyslipidemia criteria (LDL cholesterol ≥160 mg/dL or triglycerides ≥150 mg/dL) and diabetes criteria (fasting glucose ≥126 mg/dL), using the fasting subsample and corresponding survey weights. |

For each outcome and survey cycle, we estimated the proportion of missing data among eligible adults using the appropriate survey weights. When missingness exceeded 10%, we compared respondents and nonrespondents by age (≥60 years), sex, non-Hispanic Black race, and Mexican American origin using survey-weighted percentages.

We then recalculated age-standardized prevalence using weights adjusted for item nonresponse within age, sex, and race/Hispanic-origin groups and compared these estimates with the original results. Missing values were not imputed because composite outcomes were classified using all available component criteria.

**Comparison of Age-Standardized Prevalence Estimates with Published NCHS Estimates**

| **Measure** | **Survey cycle** | **NCHS (%)** | **Our estimate (%)** | **Difference (pp)** |
| --- | --- | --- | --- | --- |
| High total cholesterol ≥240 mg/dL (DB515) | 1999–2000 | 18.3 | 17.8 | −0.5 |
|  | 2001–2002 | 16.5 | 16.1 | −0.4 |
|  | 2003–2004 | 16.8 | 16.6 | −0.2 |
|  | 2005–2006 | 15.7 | 15.3 | −0.4 |
|  | 2007–2008 | 14.3 | 14.2 | −0.1 |
|  | 2009–2010 | 13.4 | 13.1 | −0.3 |
|  | 2011–2012 | 12.9 | 12.6 | −0.3 |
|  | 2013–2014 | 11.0 | 10.8 | −0.2 |
|  | 2015–2016 | 12.4 | 12.1 | −0.3 |
|  | 2017–March 2020 | 10.0 | 9.8 | −0.2 |
|  | 2021–2023 | 11.2 | 11.0 | −0.2 |
| Low HDL cholesterol <40 mg/dL, all adults (DB515) | 2007–2008 | 22.2 | 22.4 | +0.2 |
|  | 2009–2010 | 21.3 | 21.5 | +0.2 |
|  | 2011–2012 | 17.4 | 17.6 | +0.2 |
|  | 2013–2014 | 19.9 | 20.2 | +0.3 |
|  | 2015–2016 | 18.4 | 18.7 | +0.3 |
|  | 2017–March 2020 | 16.9 | 17.0 | +0.1 |
|  | 2021–2023 | 14.2 | 14.3 | +0.1 |
| Obesity, BMI ≥30 kg/m² (DB508) | 2013–2014 | 37.7 | 37.7 | 0.0 |
|  | 2015–2016 | 39.6 | 39.6 | 0.0 |
|  | 2017–March 2020 | 41.9 | 41.9 | 0.0 |
|  | 2021–2023 | 40.3 | 40.3 | 0.0 |
| Severe obesity, BMI ≥40 kg/m² (DB508) | 2013–2014 | 7.7 | 7.7 | 0.0 |
|  | 2015–2016 | 7.7 | 7.7 | 0.0 |
|  | 2017–March 2020 | 9.2 | 9.2 | 0.0 |
|  | 2021–2023 | 9.7 | 9.7 | 0.0 |

**Note:** Estimates were compared with published values from NCHS Data Briefs 515 and 508. Differences were calculated by subtracting the published NCHS estimates from the corresponding study estimates and are expressed in percentage points. Small differences in cholesterol estimates may partly reflect the inclusion of pregnant women in Data Brief 515, whereas the present study excluded them. For obesity comparisons with Data Brief 508, both analyses excluded pregnant participants.

**eTable 1. Analytic Sample and PREVENT Eligibility by Survey Cycle**

| **Survey cycle** | **Total NHANES participants** | **Adults aged ≥20 y** | **Adults examined at MEC** | **Pregnant participants excluded** | **Final analytic sample** | **PREVENT-eligible adults** | **Complete PREVENT inputs** |
| --- | --- | --- | --- | --- | --- | --- | --- |
| 1999–2000 | 9,965 | 4,880 | 4,444 | 257 | 4,187 | 2,854 | 2,470 |
| 2001–2002 | 11,039 | 5,411 | 5,027 | 296 | 4,731 | 3,159 | 2,658 |
| 2003–2004 | 10,122 | 5,041 | 4,742 | 219 | 4,523 | 2,873 | 2,444 |
| 2005–2006 | 10,348 | 4,979 | 4,773 | 325 | 4,448 | 2,914 | 2,549 |
| 2007–2008 | 10,149 | 5,935 | 5,707 | 57 | 5,650 | 3,898 | 3,379 |
| 2009–2010 | 10,537 | 6,218 | 6,059 | 68 | 5,991 | 4,122 | 3,596 |
| 2011–2012 | 9,756 | 5,560 | 5,319 | 57 | 5,262 | 3,588 | 3,138 |
| 2013–2014 | 10,175 | 5,769 | 5,588 | 65 | 5,523 | 3,873 | 3,526 |
| 2015–2016 | 9,971 | 5,719 | 5,474 | 70 | 5,404 | 3,694 | 3,352 |
| 2017–Mar 2020 | 15,560 | 9,232 | 8,544 | 87 | 8,457 | 5,805 | 4,835 |
| 2021–2023 | 11,933 | 7,809 | 6,064 | 41 | 6,023 | 4,227 | 3,835 |
| Total | 119,555 | 66,553 | 61,741 | 1,542 | 60,199 | 41,007 | 35,782 |

**Note:** Values represent unweighted participant counts. The analytic sample comprised nonpregnant adults aged 20 years or older who completed a mobile examination center (MEC) examination. PREVENT analyses were restricted to adults aged 30–79 years without self-reported cardiovascular disease (CVD) and with complete equation inputs.

**Abbreviations:** CVD, cardiovascular disease; MEC, Mobile Examination Center; PREVENT, Predicting Risk of Cardiovascular Disease EVENTs.

**eTable 2. Unweighted sample sizes for each outcome, by cycle**

| **Outcome** | **1999–2000** | **2001–2002** | **2003–2004** | **2005–2006** | **2007–2008** | | **2009–2010** | **2011–2012** | **2013–2014** | **2015–2016** | **2017–Mar 2020** | **2021–2023** |
| --- | --- | --- | --- | --- | --- | --- | --- | --- | --- | --- | --- | --- |
| **Adiposity** | | | | | | | | | | | | |
| Obesity, (BMI ≥30 or waist >102/88 cm) | 4123 | 4526 | 4440 | 4366 | 5561 | 5937 | | 5199 | 5469 | 5346 | 8313 | 5946 |
| Obesity by BMI ≥30 alone | 4117 | 4413 | 4431 | 4356 | 5550 | 5926 | | 5181 | 5455 | 5337 | 8295 | 5929 |
| Abdominal obesity by waist alone | 4026 | 4460 | 4256 | 4188 | 5322 | 5640 | | 4924 | 5210 | 5053 | 7981 | 5726 |
| Underweight (BMI <18.5) | 4117 | 4413 | 4431 | 4356 | 5550 | 5926 | | 5181 | 5455 | 5337 | 8295 | 5929 |
| Normal weight (BMI 18.5–24.9) | 4117 | 4413 | 4431 | 4356 | 5550 | 5926 | | 5181 | 5455 | 5337 | 8295 | 5929 |
| Overweight (BMI 25.0–29.9) | 4117 | 4413 | 4431 | 4356 | 5550 | 5926 | | 5181 | 5455 | 5337 | 8295 | 5929 |
| Obesity class I (BMI 30.0–34.9) | 4117 | 4413 | 4431 | 4356 | 5550 | 5926 | | 5181 | 5455 | 5337 | 8295 | 5929 |
| Obesity class II (BMI 35.0–39.9) | 4117 | 4413 | 4431 | 4356 | 5550 | 5926 | | 5181 | 5455 | 5337 | 8295 | 5929 |
| Obesity class III (BMI ≥40) | 4117 | 4413 | 4431 | 4356 | 5550 | 5926 | | 5181 | 5455 | 5337 | 8295 | 5929 |
| Mean BMI, kg/m2 | 4117 | 4413 | 4431 | 4356 | 5550 | 5926 | | 5181 | 5455 | 5337 | 8295 | 5929 |
| Mean waist circumference, cm | 4026 | 4460 | 4256 | 4188 | 5322 | 5640 | | 4924 | 5210 | 5053 | 7981 | 5726 |
| **Hypertension and blood pressure** | | | | | | | | | | | | |
| Hypertension, (≥130/80, medication or self-report) | 4186 | 4722 | 4519 | 4446 | 5650 | 5990 | | 5262 | 5523 | 5404 | 8456 | 6023 |
| Measured BP ≥130/80 mm Hg | 4041 | 4486 | 4198 | 4210 | 5377 | 5718 | | 5021 | 5333 | 5248 | 7572 | 5824 |
| Antihypertensive medication | 4137 | 4675 | 4469 | 4430 | 5643 | 5981 | | 5250 | 5516 | 5398 | 8442 | 6016 |
| Self-reported diagnosis of hypertension | 4137 | 4678 | 4472 | 4435 | 5645 | 5981 | | 5254 | 5519 | 5399 | 8445 | 6019 |
| Mean SBP, mm Hg | 4041 | 4486 | 4198 | 4210 | 5377 | 5718 | | 5021 | 5333 | 5248 | 7572 | 5824 |
| Mean DBP, mm Hg | 4000 | 4451 | 4162 | 4184 | 5347 | 5699 | | 5013 | 5317 | 5234 | 7572 | 5824 |
| **Dyslipidemia and lipids** | | | | | | | | | | | | |
| Dyslipidemia, (TC ≥240, low HDL, medication or self-report) | 4170 | 4714 | 4506 | 4431 | 5638 | 5975 | | 5256 | 5522 | 5403 | 8449 | 5754 |
| Total cholesterol ≥240 mg/dL | 3878 | 4414 | 4277 | 4173 | 5282 | 5632 | | 4863 | 5281 | 5097 | 7845 | 5462 |
| Low HDL (<40 men, <50 women) | 3877 | 4414 | 4276 | 4173 | 5282 | 5632 | | 4863 | 5281 | 5097 | 7845 | 5462 |
| Lipid-lowering medication (harmonized) | 4026 | 4519 | 4333 | 4264 | 5426 | 5809 | | 5224 | 5488 | 5366 | 8383 | 5711 |
| Self-reported high cholesterol | 4029 | 4521 | 4336 | 4268 | 5431 | 5811 | | 5232 | 5492 | 5370 | 8398 | 5721 |
| Dyslipidemia, extended (adds LDL ≥160 or TG ≥150; fasting) | 1744 | 2084 | 1886 | 1834 | 2337 | 2594 | | 2294 | 2370 | 2246 | 3767 | 2940 |
| LDL ≥160 mg/dL (fasting) | 1666 | 1976 | 1814 | 1766 | 2274 | 2522 | | 2223 | 2312 | 2206 | 3673 | 2788 |
| Triglycerides ≥150 mg/dL (fasting) | 1712 | 2043 | 1865 | 1814 | 2325 | 2567 | | 2265 | 2345 | 2230 | 3705 | 2820 |
| Mean total cholesterol, mg/dL | 3878 | 4414 | 4277 | 4173 | 5282 | 5632 | | 4863 | 5281 | 5097 | 7845 | 5462 |
| Mean HDL cholesterol, mg/dL | 3877 | 4414 | 4276 | 4173 | 5282 | 5632 | | 4863 | 5281 | 5097 | 7845 | 5462 |
| Mean non-HDL cholesterol, mg/dL | 3876 | 4414 | 4276 | 4173 | 5282 | 5632 | | 4863 | 5281 | 5097 | 7845 | 5462 |
| Mean LDL cholesterol, mg/dL (fasting) | 1666 | 1976 | 1814 | 1766 | 2274 | 2522 | | 2223 | 2312 | 2206 | 3673 | 2788 |
| Mean triglycerides, mg/dL (fasting) | 1712 | 2043 | 1865 | 1814 | 2325 | 2567 | | 2265 | 2345 | 2230 | 3705 | 2820 |
| Geometric mean triglycerides, mg/dL (fasting) | 1712 | 2043 | 1865 | 1814 | 2325 | 2567 | | 2265 | 2345 | 2230 | 3705 | 2820 |
| **Diabetes and glycemia** | | | | | | | | | | | | |
| Diabetes, (self-report, HbA1c ≥6.5% or medication) | 4186 | 4731 | 4523 | 4448 | 5650 | 5991 | | 5262 | 5523 | 5404 | 8457 | 5755 |
| Diagnosed diabetes (self-report) | 4183 | 4730 | 4522 | 4442 | 5644 | 5988 | | 5258 | 5520 | 5401 | 8452 | 5755 |
| HbA1c ≥6.5% | 3962 | 4493 | 4317 | 4195 | 5306 | 5687 | | 4970 | 5331 | 5141 | 8005 | 5730 |
| Insulin or oral glucose-lowering medication (harmonized) | 4183 | 4730 | 4522 | 4440 | 5641 | 5988 | | 5257 | 5519 | 5400 | 8449 | 5753 |
| Undiagnosed diabetes (no diagnosis, HbA1c ≥6.5%) | 3996 | 4510 | 4342 | 4218 | 5351 | 5726 | | 5002 | 5353 | 5159 | 8064 | 5733 |
| Diabetes, extended (adds fasting glucose ≥126; fasting) | 1746 | 2085 | 1887 | 1836 | 2337 | 2595 | | 2294 | 2370 | 2246 | 3769 | 2940 |
| Fasting glucose ≥126 mg/dL (fasting) | 1739 | 2080 | 1875 | 1836 | 2337 | 2595 | | 2294 | 2370 | 2246 | 3769 | 2940 |
| Mean HbA1c, % | 3962 | 4493 | 4317 | 4195 | 5306 | 5687 | | 4970 | 5331 | 5141 | 8005 | 5730 |
| Mean fasting glucose, mg/dL (fasting) | 1739 | 2080 | 1875 | 1836 | 2337 | 2595 | | 2294 | 2370 | 2246 | 3769 | 2940 |
| **Chronic kidney disease** | | | | | | | | | | | | |
| CKD (eGFR <60, UACR ≥30, weak/failing kidneys or dialysis) | 4187 | 4730 | 4522 | 4448 | 5650 | 5991 | | 5262 | 5522 | 5404 | 8457 | 5755 |
| eGFR <60 mL/min/1.73 m2 | 3881 | 4421 | 4256 | 4155 | 5264 | 5619 | | 4839 | 5267 | 5088 | 7806 | 5407 |
| UACR ≥30 mg/g | 4067 | 4577 | 4379 | 4305 | 5468 | 5861 | | 5144 | 5404 | 5279 | 8234 | 5643 |
| Self-reported weak or failing kidneys | 4178 | 4714 | 4515 | 4435 | 5641 | 5979 | | 5257 | 5516 | 5397 | 8443 | 5744 |
| Dialysis in past 12 months (2001+) |  | 4714 | 4515 | 4435 | 5641 | 5979 | | 5257 | 5516 | 5396 | 8443 | 5744 |
| Mean eGFR, mL/min/1.73 m2 | 3881 | 4421 | 4256 | 4155 | 5264 | 5619 | | 4839 | 5267 | 5088 | 7806 | 5407 |
| **Smoking** | | | | | | | | | | | | |
| Current smoking | 4178 | 4724 | 4517 | 4444 | 5644 | 5991 | | 5255 | 5521 | 5394 | 8453 | 6016 |
| Former smoking | 4178 | 4724 | 4517 | 4444 | 5644 | 5991 | | 5255 | 5521 | 5394 | 8453 | 6016 |
| Never smoking | 4178 | 4724 | 4517 | 4444 | 5644 | 5991 | | 5255 | 5521 | 5394 | 8453 | 6016 |
| Quit ratio (former / ever smokers) | 2007 | 2352 | 2285 | 2145 | 2686 | 2775 | | 2262 | 2424 | 2272 | 3538 | 2502 |
| **Alcohol** | | | | | | | | | | | | |
| Drinks at least once a month (past 12 months) | 3914 | 4358 | 4104 | 4034 | 5160 | 5268 | | 4634 | 5035 | 4893 | 7897 | 5195 |
| Drinks per drinking day, past-year drinkers (mean) | 2511 | 2844 | 2583 | 2645 | 3274 | 3624 | | 3116 | 3416 | 3219 | 5553 | 3896 |
| Exceeds >2 (men) / >1 (women) drinks per drinking day, drinkers | 2511 | 2844 | 2583 | 2645 | 3274 | 3624 | | 3116 | 3416 | 3219 | 5553 | 3896 |
| **Prior cardiovascular disease** | | | | | | | | | | | | |
| Prior CVD (any) | 4187 | 4731 | 4521 | 4448 | 5649 | 5990 | | 5260 | 5522 | 5404 | 8456 | 6023 |
| Heart failure | 4165 | 4712 | 4501 | 4434 | 5625 | 5974 | | 5247 | 5516 | 5393 | 8436 | 6014 |
| Coronary heart disease | 4157 | 4699 | 4500 | 4424 | 5622 | 5965 | | 5242 | 5507 | 5380 | 8431 | 5998 |
| Angina | 4164 | 4705 | 4504 | 4432 | 5628 | 5975 | | 5246 | 5517 | 5388 | 8419 | 6008 |
| Myocardial infarction | 4173 | 4724 | 4514 | 4440 | 5640 | 5976 | | 5256 | 5520 | 5397 | 8444 | 6013 |
| Stroke | 4185 | 4724 | 4517 | 4441 | 5632 | 5985 | | 5258 | 5518 | 5400 | 8442 | 6012 |
| **Awareness, treatment, and control among adults with each condition** | | | | | | | | | | | | |
| Hypertension: aware | 2386 | 2578 | 2509 | 2314 | 3041 | 3135 | | 2801 | 2926 | 2967 | 4836 | 3447 |
| Hypertension: treated | 2386 | 2575 | 2506 | 2309 | 3039 | 3135 | | 2797 | 2923 | 2966 | 4833 | 3444 |
| Hypertension: controlled (<130/80) | 2315 | 2486 | 2367 | 2208 | 2927 | 3032 | | 2725 | 2835 | 2910 | 4508 | 3368 |
| Dyslipidemia: aware | 2309 | 2461 | 2391 | 2254 | 3170 | 3319 | | 2887 | 3175 | 3014 | 4691 | 3385 |
| Dyslipidemia: treated | 2306 | 2459 | 2388 | 2250 | 3165 | 3317 | | 2879 | 3171 | 3010 | 4676 | 3375 |
| Dyslipidemia: LDL <100 mg/dL (fasting) | 1003 | 1086 | 997 | 888 | 1269 | 1428 | | 1210 | 1312 | 1265 | 2099 | 1667 |
| Diabetes: aware | 543 | 573 | 614 | 587 | 912 | 904 | | 845 | 853 | 957 | 1605 | 974 |
| Diabetes: treated | 543 | 573 | 614 | 585 | 909 | 904 | | 844 | 852 | 956 | 1602 | 972 |
| Diabetes: HbA1c <7% | 506 | 555 | 588 | 559 | 861 | 863 | | 809 | 828 | 938 | 1541 | 971 |
| **PREVENT predicted risk** | | | | | | | | | | | | |
| 10-year total CVD risk, mean % | 2470 | 2658 | 2444 | 2549 | 3379 | 3596 | | 3138 | 3526 | 3352 | 4835 | 3835 |
| 10-year ASCVD risk, mean % | 2470 | 2658 | 2444 | 2549 | 3379 | 3596 | | 3138 | 3526 | 3352 | 4835 | 3835 |
| 10-year heart failure risk, mean % | 2470 | 2658 | 2444 | 2549 | 3379 | 3596 | | 3138 | 3526 | 3352 | 4835 | 3835 |
| 10-year total CVD risk <5% | 2470 | 2658 | 2444 | 2549 | 3379 | 3596 | | 3138 | 3526 | 3352 | 4835 | 3835 |
| 10-year total CVD risk 5 to <7.5% | 2470 | 2658 | 2444 | 2549 | 3379 | 3596 | | 3138 | 3526 | 3352 | 4835 | 3835 |
| 10-year total CVD risk 7.5 to <20% | 2470 | 2658 | 2444 | 2549 | 3379 | 3596 | | 3138 | 3526 | 3352 | 4835 | 3835 |
| 10-year total CVD risk ≥20% | 2470 | 2658 | 2444 | 2549 | 3379 | 3596 | | 3138 | 3526 | 3352 | 4835 | 3835 |
| 10-year total CVD risk ≥7.5% | 2470 | 2658 | 2444 | 2549 | 3379 | 3596 | | 3138 | 3526 | 3352 | 4835 | 3835 |
| 30-year total CVD risk, mean % (ages 30–59) | 1617 | 1862 | 1619 | 1793 | 2253 | 2481 | | 2192 | 2487 | 2316 | 3174 | 2139 |
| 30-year ASCVD risk, mean % (ages 30–59) | 1617 | 1862 | 1619 | 1793 | 2253 | 2481 | | 2192 | 2487 | 2316 | 3174 | 2139 |
| 30-year heart failure risk, mean % (ages 30–59) | 1617 | 1862 | 1619 | 1793 | 2253 | 2481 | | 2192 | 2487 | 2316 | 3174 | 2139 |

**Note:** Values represent unweighted sample sizes (n) for each outcome and NHANES survey cycle, including only eligible adults with available data. Sample sizes vary across outcomes because of differences in data availability, eligibility criteria, and fasting subsample requirements. Analyses of awareness, treatment, and control were restricted to adults with the corresponding condition, while PREVENT analyses included only adults within the eligible age ranges. Dialysis data were not available for 1999–2000.

**Abbreviations:** ASCVD, atherosclerotic cardiovascular disease; BMI, body mass index; BP, blood pressure; CKD, chronic kidney disease; CVD, cardiovascular disease; DBP, diastolic blood pressure; eGFR, estimated glomerular filtration rate; HbA1c, glycated hemoglobin; HDL, high-density lipoprotein; LDL, low-density lipoprotein; SBP, systolic blood pressure; TC, total cholesterol; TG, triglycerides; UACR, urine albumin-to-creatinine ratio.

**eTable 3. Endpoint estimates and trends across the study period**

| **Outcome** | **1999–2000** | **2021–2023** | **Change (95% CI)** | **P for change** | **Trend measure** | **Per decade (95% CI)** | **P for trend** | **P for nonlinearity** |
| --- | --- | --- | --- | --- | --- | --- | --- | --- |
| **Adiposity** |  |  |  |  |  |  |  |  |
| Obesity, primary (BMI ≥30 or waist >102/88 cm) | 47.4 | 57.2 | 9.8 (4.2 to 15.3) | 0.001 | OR | 1.23 (1.16–1.30) | <0.001 | 0.072 |
| Obesity by BMI ≥30 alone | 30.2 | 40.3 | 10.2 (5.2 to 15.2) | <0.001 | OR | 1.28 (1.21–1.35) | <0.001 | 0.356 |
| Abdominal obesity by waist alone | 46.6 | 56.2 | 9.6 (4.1 to 15.2) | 0.001 | OR | 1.22 (1.15–1.29) | <0.001 | 0.035 |
| Underweight (BMI <18.5) | 2.1 | 1.6 | −0.4 (−1.2 to 0.3) | 0.218 | OR | 0.88 (0.78–1.00) | 0.046 | 0.457 |
| Normal weight (BMI 18.5–24.9) | 33.7 | 26.3 | −7.5 (−11.8 to −3.1) | 0.001 | OR | 0.83 (0.79–0.88) | <0.001 | 0.672 |
| Overweight (BMI 25.0–29.9) | 34.0 | 31.7 | −2.3 (−5.2 to 0.7) | 0.127 | OR | 0.93 (0.90–0.97) | <0.001 | 0.922 |
| Obesity class I (BMI 30.0–34.9) | 17.1 | 20.3 | 3.2 (0.5 to 5.9) | 0.022 | OR | 1.11 (1.06–1.16) | <0.001 | 0.022 |
| Obesity class II (BMI 35.0–39.9) | 8.3 | 10.3 | 2.0 (0.2 to 3.8) | 0.034 | OR | 1.21 (1.13–1.28) | <0.001 | 0.920 |
| Obesity class III (BMI ≥40) | 4.7 | 9.7 | 5.0 (3.2 to 6.9) | <0.001 | OR | 1.47 (1.35–1.59) | <0.001 | 0.838 |
| Mean BMI, kg/m2 | 28.0 | 29.7 | 1.7 (1.0 to 2.4) | <0.001 | Difference | 0.89 (0.70–1.08) | <0.001 | 0.940 |
| Mean waist circumference, cm | 95.6 | 100.2 | 4.5 (2.7 to 6.3) | <0.001 | Difference | 2.20 (1.73–2.67) | <0.001 | 0.106 |
| **Hypertension and blood pressure** |  |  |  |  |  |  |  |  |
| Hypertension, primary (≥130/80, medication or self-report) | 50.6 | 48.1 | −2.5 (−6.0 to 1.1) | 0.168 | OR | 0.96 (0.92–1.02) | 0.167 | 0.054 |
| Measured BP ≥130/80 mm Hg | 44.0 | 37.2 | −6.8 (−10.1 to −3.5) | <0.001 | OR | 0.88 (0.84–0.92) | <0.001 | <0.001 |
| Antihypertensive medication | 17.6 | 22.2 | 4.6 (2.2 to 7.1) | <0.001 | OR | 1.14 (1.08–1.20) | <0.001 | <0.001 |
| Self-reported diagnosis of hypertension | 25.5 | 28.8 | 3.3 (0.4 to 6.2) | 0.026 | OR | 1.08 (1.03–1.14) | 0.003 | <0.001 |
| Mean SBP, mm Hg | 124.2 | 120.2 | −4.0 (−5.5 to −2.5) | <0.001 | Difference | −1.52 (−1.92 to −1.13) | <0.001 | 0.100 |
| Mean DBP, mm Hg | 73.1 | 74.9 | 1.8 (0.8 to 2.7) | <0.001 | Difference | 1.02 (0.72–1.31) | <0.001 | <0.001 |
| **Dyslipidemia and lipids** |  |  |  |  |  |  |  |  |
| Dyslipidemia, primary (TC ≥240, low HDL, medication or self-report) | 57.5 | 53.4 | −4.1 (−8.3 to 0.1) | 0.057 | OR | 0.98 (0.94–1.03) | 0.507 | 0.956 |
| Total cholesterol ≥240 mg/dL | 17.8 | 11.0 | −6.7 (−9.2 to −4.2) | <0.001 | OR | 0.73 (0.69–0.78) | <0.001 | 0.101 |
| Low HDL (<40 men, <50 women) | 39.3 | 26.7 | −12.7 (−17.4 to −7.9) | <0.001 | OR | 0.86 (0.82–0.91) | <0.001 | 0.383 |
| Lipid-lowering medication | 8.8 | 17.8 | 9.0 (7.5 to 10.5) | <0.001 | OR | 1.30 (1.24–1.36) | <0.001 | <0.001 |
| Self-reported high cholesterol | 25.9 | 33.3 | 7.3 (5.1 to 9.5) | <0.001 | OR | 1.19 (1.14–1.24) | <0.001 | 0.002 |
| Dyslipidemia, extended (adds LDL ≥160 or TG ≥150; fasting) | 67.8 | 57.4 | −10.5 (−14.8 to −6.1) | <0.001 | OR | 0.87 (0.83–0.92) | <0.001 | 0.019 |
| LDL ≥160 mg/dL (fasting) | 15.9 | 8.6 | −7.3 (−10.8 to −3.8) | <0.001 | OR | 0.75 (0.68–0.83) | <0.001 | 0.027 |
| Triglycerides ≥150 mg/dL (fasting) | 33.1 | 20.9 | −12.2 (−16.0 to −8.4) | <0.001 | OR | 0.68 (0.64–0.72) | <0.001 | 0.156 |
| Mean total cholesterol, mg/dL | 203.6 | 188.1 | −15.5 (−18.9 to −12.1) | <0.001 | Difference | −8.02 (−9.05 to −6.99) | <0.001 | 0.049 |
| Mean HDL cholesterol, mg/dL | 50.3 | 53.3 | 3.0 (1.3 to 4.7) | 0.001 | Difference | 0.92 (0.50–1.34) | <0.001 | 0.011 |
| Mean non-HDL cholesterol, mg/dL | 153.3 | 134.8 | −18.5 (−22.3 to −14.7) | <0.001 | Difference | −8.93 (−9.97 to −7.89) | <0.001 | 0.002 |
| Mean LDL cholesterol, mg/dL (fasting) | 126.0 | 110.5 | −15.5 (−18.8 to −12.2) | <0.001 | Difference | −5.78 (−6.78 to −4.77) | <0.001 | <0.001 |
| Mean triglycerides, mg/dL (fasting) | 143.4 | 118.7 | −24.6 (−35.0 to −14.3) | <0.001 | Difference | −18.45 (−21.53 to −15.37) | <0.001 | 0.018 |
| Geometric mean triglycerides, mg/dL (fasting) | 121.5 | 99.9 | −21.6 (−28.0 to −15.2) | <0.001 | Ratio | 0.87 (0.86–0.88) | <0.001 | 0.002 |
| **Diabetes and glycemia** |  |  |  |  |  |  |  |  |
| Diabetes, primary (self-report, HbA1c ≥6.5% or medication) | 8.2 | 12.7 | 4.5 (2.7 to 6.3) | <0.001 | OR | 1.26 (1.19–1.34) | <0.001 | 0.211 |
| Diagnosed diabetes (self-report) | 6.6 | 10.6 | 4.1 (2.5 to 5.7) | <0.001 | OR | 1.28 (1.21–1.37) | <0.001 | 0.262 |
| HbA1c ≥6.5% | 6.4 | 9.7 | 3.2 (1.5 to 5.0) | <0.001 | OR | 1.29 (1.20–1.38) | <0.001 | 0.782 |
| Insulin or oral glucose-lowering medication | 5.4 | 8.9 | 3.6 (2.0 to 5.2) | <0.001 | OR | 1.28 (1.19–1.37) | <0.001 | 0.151 |
| Undiagnosed diabetes (no diagnosis, HbA1c ≥6.5%) | 1.7 | 2.1 | 0.4 (−0.2 to 1.0) | 0.209 | OR | 1.10 (1.00–1.22) | 0.060 | 0.291 |
| Diabetes, extended (adds fasting glucose ≥126; fasting) | 8.9 | 14.3 | 5.4 (2.4 to 8.4) | <0.001 | OR | 1.27 (1.17–1.37) | <0.001 | 0.174 |
| Fasting glucose ≥126 mg/dL (fasting) | 6.9 | 10.4 | 3.4 (1.1 to 5.8) | 0.006 | OR | 1.28 (1.18–1.39) | <0.001 | 0.315 |
| Mean HbA1c, % | 5.43 | 5.68 | 0.25 (0.14 to 0.36) | <0.001 | Difference | 0.11 (0.09–0.13) | <0.001 | 0.071 |
| Mean fasting glucose, mg/dL (fasting) | 100.9 | 108.0 | 7.1 (4.0 to 10.2) | <0.001 | Difference | 3.83 (2.98–4.68) | <0.001 | 0.413 |
| **Chronic kidney disease** |  |  |  |  |  |  |  |  |
| CKD (eGFR <60, UACR ≥30, weak/failing kidneys or dialysis) | 14.7 | 13.5 | −1.2 (−3.2 to 0.9) | 0.246 | OR | 0.98 (0.93–1.03) | 0.339 | 0.559 |
| eGFR <60 mL/min/1.73 m2 | 5.4 | 5.3 | −0.1 (−1.2 to 1.0) | 0.899 | OR | 0.91 (0.85–0.97) | 0.006 | 0.975 |
| UACR ≥30 mg/g | 10.7 | 9.4 | −1.3 (−2.7 to 0.1) | 0.066 | OR | 0.98 (0.93–1.03) | 0.476 | 0.155 |
| Self-reported weak or failing kidneys | 2.4 | 2.5 | 0.0 (−1.0 to 1.0) | 0.946 | OR | 1.19 (1.07–1.32) | 0.002 | 0.965 |
| Dialysis in past 12 months (first cycle 2001–2002) | 0.2 | 0.1 | −0.1 (−0.2 to 0.1) | 0.463 | OR | 1.10 (0.88–1.39) | 0.400 | 0.025 |
| Mean eGFR, mL/min/1.73 m2 | 95.1 | 97.3 | 2.2 (1.0 to 3.5) | <0.001 | Difference | 1.30 (0.89–1.70) | <0.001 | 0.068 |
| **Smoking** |  |  |  |  |  |  |  |  |
| Current smoking | 24.4 | 14.9 | −9.5 (−13.2 to −5.8) | <0.001 | OR | 0.76 (0.71–0.81) | <0.001 | 0.158 |
| Former smoking | 25.0 | 22.1 | −2.9 (−5.7 to −0.1) | 0.042 | OR | 0.95 (0.90–0.99) | 0.019 | 0.942 |
| Never smoking | 50.6 | 63.0 | 12.4 (7.3 to 17.5) | <0.001 | OR | 1.25 (1.18–1.32) | <0.001 | 0.453 |
| Quit ratio (former / ever smokers) | 49.3 | 58.6 | 9.2 (4.6 to 13.9) | <0.001 | OR | 1.18 (1.11–1.26) | <0.001 | 0.212 |
| **Alcohol** |  |  |  |  |  |  |  |  |
| Drinks at least once a month (past 12 months) | 51.3 | 55.8 | 4.4 (−1.9 to 10.7) | 0.161 | OR | 1.11 (1.03–1.20) | 0.006 | 0.069 |
| Drinks per drinking day, past-year drinkers (mean) | 2.78 | 2.78 | 0.00 (−0.25 to 0.26) | 0.976 | Difference | −0.03 (−0.11–0.04) | 0.364 | 0.590 |
| Exceeds >2 (men) / >1 (women) drinks per drinking day, drinkers | 49.0 | 54.4 | 5.4 (0.5 to 10.3) | 0.033 | OR | 1.08 (1.02–1.15) | 0.013 | 0.632 |
| **Prior cardiovascular disease** |  |  |  |  |  |  |  |  |
| Prior CVD (any) | 8.7 | 8.2 | −0.5 (−2.1 to 1.1) | 0.531 | OR | 0.95 (0.89–1.02) | 0.141 | 0.238 |
| Heart failure | 2.2 | 2.5 | 0.3 (−0.4 to 1.0) | 0.383 | OR | 0.99 (0.90–1.09) | 0.838 | 0.581 |
| Coronary heart disease | 3.4 | 3.3 | −0.0 (−0.8 to 0.7) | 0.933 | OR | 0.97 (0.87–1.07) | 0.507 | 0.082 |
| Angina | 3.1 | 1.4 | −1.6 (−2.5 to −0.8) | <0.001 | OR | 0.72 (0.64–0.80) | <0.001 | 0.221 |
| Myocardial infarction | 3.6 | 2.8 | −0.8 (−1.6 to 0.0) | 0.051 | OR | 0.89 (0.82–0.98) | 0.017 | 0.746 |
| Stroke | 2.5 | 3.1 | 0.6 (−0.3 to 1.4) | 0.167 | OR | 1.07 (0.98–1.17) | 0.116 | 0.675 |
| **Awareness, treatment, and control among adults with each condition** |  |  |  |  |  |  |  |  |
| Hypertension: aware | 45.6 | 54.0 | 8.4 (4.0 to 12.8) | <0.001 | OR | 1.20 (1.13–1.26) | <0.001 | <0.001 |
| Hypertension: treated | 27.8 | 38.0 | 10.3 (6.5 to 14.0) | <0.001 | OR | 1.25 (1.18–1.32) | <0.001 | <0.001 |
| Hypertension: controlled (<130/80) | 14.2 | 21.5 | 7.3 (4.7 to 9.9) | <0.001 | OR | 1.27 (1.21–1.34) | <0.001 | <0.001 |
| Dyslipidemia: aware | 42.7 | 57.2 | 14.5 (10.6 to 18.3) | <0.001 | OR | 1.33 (1.26–1.40) | <0.001 | 0.007 |
| Dyslipidemia: treated | 13.8 | 28.4 | 14.6 (12.3 to 16.9) | <0.001 | OR | 1.36 (1.29–1.43) | <0.001 | <0.001 |
| Dyslipidemia: LDL <100 mg/dL (fasting) | 17.0 | 34.2 | 17.3 (12.8 to 21.7) | <0.001 | OR | 1.44 (1.34–1.55) | <0.001 | <0.001 |
| Diabetes: aware | 78.2 | 81.3 | 3.0 (−6.4 to 12.5) | 0.519 | OR | 1.13 (1.02–1.26) | 0.018 | 0.739 |
| Diabetes: treated | 57.6 | 68.5 | 10.9 (0.3 to 21.5) | 0.045 | OR | 1.08 (0.99–1.18) | 0.082 | 0.461 |
| Diabetes: HbA1c <7% | 36.4 | 41.9 | 5.6 (−8.6 to 19.8) | 0.430 | OR | 1.02 (0.93–1.12) | 0.704 | <0.001 |
| **PREVENT predicted risk** |  |  |  |  |  |  |  |  |
| 10-year total CVD risk, mean % | 5.66 | 5.28 | −0.39 (−0.74 to −0.03) | 0.032 | Difference | −0.10 (−0.21 to −0.00) | 0.045 | 0.551 |
| 10-year ASCVD risk, mean % | 3.66 | 3.20 | −0.45 (−0.70 to −0.21) | <0.001 | Difference | −0.16 (−0.22 to −0.09) | <0.001 | 0.241 |
| 10-year heart failure risk, mean % | 3.05 | 3.19 | 0.14 (−0.09 to 0.37) | 0.214 | Difference | 0.10 (0.03–0.17) | 0.004 | 0.588 |
| 10-year total CVD risk <5% | 64.5 | 65.4 | 0.9 (−2.4 to 4.2) | 0.579 | OR | 0.99 (0.92–1.06) | 0.722 | 0.959 |
| 10-year total CVD risk 5 to <7.5% | 10.3 | 10.4 | 0.1 (−2.2 to 2.4) | 0.947 | OR | 1.03 (0.96–1.11) | 0.398 | 0.333 |
| 10-year total CVD risk 7.5 to <20% | 20.1 | 20.0 | −0.2 (−1.9 to 1.6) | 0.849 | OR | 1.03 (0.98–1.09) | 0.267 | 0.251 |
| 10-year total CVD risk ≥20% | 5.0 | 4.2 | −0.8 (−2.0 to 0.4) | 0.181 | OR | 0.89 (0.82–0.98) | 0.013 | 0.997 |
| 10-year total CVD risk ≥7.5% | 25.2 | 24.2 | −1.0 (−2.9 to 0.9) | 0.301 | OR | 0.99 (0.92–1.06) | 0.696 | 0.305 |
| 30-year total CVD risk, mean % (ages 30–59) | 15.32 | 14.14 | −1.18 (−2.19 to −0.17) | 0.023 | Difference | −0.37 (−0.63 to −0.12) | 0.004 | 0.857 |
| 30-year ASCVD risk, mean % (ages 30–59) | 9.95 | 8.91 | −1.04 (−1.77 to −0.31) | 0.007 | Difference | −0.40 (−0.58 to −0.22) | <0.001 | 0.808 |
| 30-year heart failure risk, mean % (ages 30–59) | 7.94 | 8.36 | 0.42 (−0.21 to 1.05) | 0.180 | Difference | 0.31 (0.14–0.48) | <0.001 | 0.661 |

**Note:** Estimates are age-standardized unless otherwise specified. Changes represent absolute differences between 1999–2000 and 2021–2023, expressed in percentage points for prevalence outcomes and original units for continuous measures. Trends per decade are reported as odds ratios (ORs), absolute differences, or ratios, as appropriate. P values correspond to first-to-last differences, linear trends, and quadratic nonlinearity, respectively. PREVENT risk estimates are restricted to the age groups eligible for each equation.

**Abbreviations:** ASCVD, atherosclerotic cardiovascular disease; BMI, body mass index; BP, blood pressure; CI, confidence interval; CKD, chronic kidney disease; CVD, cardiovascular disease; DBP, diastolic blood pressure; eGFR, estimated glomerular filtration rate; HbA1c, glycated hemoglobin; HDL, high-density lipoprotein; LDL, low-density lipoprotein; OR, odds ratio; SBP, systolic blood pressure; TC, total cholesterol; TG, triglycerides; UACR, urine albumin-to-creatinine ratio.

**eTable 4. Subgroup trends and tests for interaction**

| **Subgroup / level** | **1999–2000** | **2021–2023** | **Change (95% CI)** | **OR per decade (95% CI)** | **P for trend** | **P for interaction** |
| --- | --- | --- | --- | --- | --- | --- |
| **Obesity, (BMI ≥30 or waist >102/88 cm)** |  |  |  |  |  |  |
| **Sex:** Men | 38.8 | 48.9 | 10.1 (4.5 to 15.7) | 1.21 (1.13–1.30) | <0.001 | 0.296 |
| Women | 55.7 | 65.3 | 9.6 (3.1 to 16.2) | 1.26 (1.17–1.35) | <0.001 | 0.296 |
| **Age group:** 20–39 | 35.3 | 46.0 | 10.7 (4.2 to 17.2) | 1.27 (1.18–1.38) | <0.001 | 0.006 |
| 40–59 | 52.0 | 62.5 | 10.5 (3.1 to 17.8) | 1.24 (1.15–1.34) | <0.001 | 0.006 |
| 60+ | 61.0 | 68.1 | 7.1 (1.8 to 12.4) | 1.13 (1.06–1.21) | <0.001 | 0.006 |
| **Race and Hispanic origin:** Non-Hispanic White | 46.4 | 57.5 | 11.0 (4.9 to 17.2) | 1.26 (1.18–1.34) | <0.001 | 0.045 |
| Non-Hispanic Black | 54.4 | 64.4 | 9.9 (3.5 to 16.4) | 1.20 (1.12–1.30) | <0.001 | 0.045 |
| Mexican American | 50.5 | 62.2 | 11.8 (2.6 to 20.9) | 1.42 (1.29–1.56) | <0.001 | 0.045 |
| Non-Hispanic other or multiracial | 39.5 | 44.0 | 4.6 (−10.9 to 20.0) | 1.22 (1.04–1.44) | 0.017 | 0.045 |
| All Hispanic (2007–2023 only) † | 56.4 | 60.5 | 4.1 (−4.2 to 12.4) | 1.15 (0.96–1.36) | 0.126 |  |
| **Family income-to-poverty ratio:** ≤1.30 | 51.9 | 61.1 | 9.2 (4.0 to 14.5) | 1.21 (1.13–1.29) | <0.001 | 0.735 |
| 1.31–3.50 | 52.2 | 60.0 | 7.8 (1.1 to 14.4) | 1.23 (1.14–1.33) | <0.001 | 0.735 |
| >3.50 | 42.2 | 53.0 | 10.8 (3.9 to 17.8) | 1.25 (1.15–1.36) | <0.001 | 0.735 |
| **Education:** Less than high school | 52.1 | 62.6 | 10.5 (2.9 to 18.1) | 1.15 (1.07–1.24) | <0.001 | 0.048 |
| High school or GED | 49.9 | 61.6 | 11.7 (4.4 to 19.0) | 1.26 (1.16–1.36) | <0.001 | 0.048 |
| Some college or associate degree | 48.5 | 61.3 | 12.8 (6.3 to 19.2) | 1.32 (1.23–1.42) | <0.001 | 0.048 |
| College graduate or above | 37.0 | 49.1 | 12.1 (4.2 to 20.0) | 1.25 (1.13–1.37) | <0.001 | 0.048 |
| **Hypertension, (≥130/80, medication or self-report)** |  |  |  |  |  |  |
| **Sex:** Men | 55.2 | 52.5 | −2.7 (−7.4 to 2.1) | 0.97 (0.91–1.04) | 0.436 | 0.780 |
| Women | 45.8 | 43.7 | −2.1 (−5.9 to 1.7) | 0.96 (0.90–1.02) | 0.144 | 0.780 |
| **Age group:** 20–39 | 28.1 | 27.4 | −0.7 (−6.2 to 4.8) | 1.01 (0.93–1.09) | 0.824 | <0.001 |
| 40–59 | 56.1 | 54.7 | −1.4 (−6.0 to 3.1) | 1.00 (0.93–1.06) | 0.916 | <0.001 |
| 60+ | 80.2 | 73.0 | −7.2 (−11.8 to −2.5) | 0.85 (0.78–0.91) | <0.001 | <0.001 |
| **Race and Hispanic origin:** Non-Hispanic White | 49.2 | 47.2 | −2.0 (−6.7 to 2.7) | 0.95 (0.89–1.02) | 0.143 | 0.443 |
| Non-Hispanic Black | 60.4 | 61.5 | 1.0 (−5.3 to 7.4) | 1.02 (0.93–1.11) | 0.712 | 0.443 |
| Mexican American | 47.8 | 45.6 | −2.2 (−10.6 to 6.1) | 1.00 (0.90–1.11) | 0.950 | 0.443 |
| Non-Hispanic other or multiracial | 55.2 | 45.7 | −9.5 (−23.2 to 4.1) | 0.92 (0.79–1.07) | 0.283 | 0.443 |
| All Hispanic (2007–2023 only) † | 43.9 | 45.6 | 1.8 (−4.6 to 8.1) | 1.07 (0.90–1.28) | 0.421 |  |
| **Family income-to-poverty ratio:** ≤1.30 | 54.3 | 52.1 | −2.2 (−8.6 to 4.2) | 1.00 (0.92–1.09) | 0.964 | 0.071 |
| 1.31–3.50 | 51.7 | 50.3 | −1.4 (−5.9 to 3.1) | 1.00 (0.93–1.06) | 0.935 | 0.071 |
| >3.50 | 49.2 | 45.1 | −4.0 (−9.2 to 1.1) | 0.92 (0.85–0.99) | 0.036 | 0.071 |
| **Education:** Less than high school | 52.0 | 52.9 | 1.0 (−5.8 to 7.7) | 0.96 (0.88–1.04) | 0.290 | 0.005 |
| High school or GED | 54.2 | 52.3 | −1.9 (−8.2 to 4.4) | 1.00 (0.92–1.08) | 0.901 | 0.005 |
| Some college or associate degree | 51.0 | 49.5 | −1.5 (−6.0 to 2.9) | 1.07 (1.00–1.15) | 0.041 | 0.005 |
| College graduate or above | 44.7 | 42.4 | −2.2 (−7.3 to 2.8) | 0.91 (0.84–1.00) | 0.040 | 0.005 |
| **Dyslipidemia, (TC ≥240, low HDL, medication or self-report)** |  |  |  |  |  |  |
| Sex: Men | 55.4 | 51.1 | −4.3 (−8.4 to −0.2) | 0.98 (0.93–1.03) | 0.430 | 0.525 |
| Women | 59.6 | 55.6 | −4.0 (−9.5 to 1.5) | 0.99 (0.93–1.06) | 0.790 | 0.525 |
| **Age group:** 20–39 | 49.7 | 38.6 | −11.2 (−17.2 to −5.2) | 0.90 (0.84–0.96) | 0.001 | <0.001 |
| 40–59 | 59.6 | 59.4 | −0.2 (−5.8 to 5.3) | 1.03 (0.96–1.11) | 0.379 | <0.001 |
| 60+ | 67.4 | 69.2 | 1.9 (−1.7 to 5.4) | 1.06 (1.00–1.12) | 0.050 | <0.001 |
| **Race and Hispanic origin:** Non-Hispanic White | 59.1 | 53.6 | −5.5 (−10.7 to −0.3) | 0.98 (0.92–1.04) | 0.446 | 0.607 |
| Non-Hispanic Black | 47.1 | 51.2 | 4.1 (−4.3 to 12.5) | 1.05 (0.95–1.14) | 0.339 | 0.607 |
| Mexican American | 54.9 | 52.9 | −2.1 (−9.0 to 4.8) | 1.03 (0.94–1.13) | 0.518 | 0.607 |
| Non-Hispanic other or multiracial | 57.0 | 54.6 | −2.5 (−15.5 to 10.5) | 1.00 (0.87–1.15) | 0.984 | 0.607 |
| All Hispanic (2007–2023 only) † | 57.3 | 53.2 | −4.1 (−11.8 to 3.5) | 0.90 (0.75–1.07) | 0.224 |  |
| **Family income-to-poverty ratio:** ≤1.30 | 60.5 | 55.4 | −5.1 (−9.6 to −0.6) | 0.95 (0.89–1.02) | 0.136 | 0.431 |
| 1.31–3.50 | 58.6 | 54.6 | −4.0 (−8.6 to 0.5) | 1.00 (0.94–1.06) | 0.977 | 0.431 |
| >3.50 | 55.8 | 51.1 | −4.8 (−11.6 to 2.1) | 0.99 (0.93–1.06) | 0.792 | 0.431 |
| **Education:** Less than high school | 59.2 | 58.5 | −0.7 (−8.3 to 6.8) | 1.07 (0.99–1.15) | 0.087 | 0.011 |
| High school or GED | 62.3 | 53.8 | −8.5 (−14.5 to −2.4) | 0.92 (0.85–0.99) | 0.031 | 0.011 |
| Some college or associate degree | 55.8 | 57.4 | 1.5 (−4.1 to 7.2) | 1.06 (0.99–1.13) | 0.082 | 0.011 |
| College graduate or above | 52.7 | 48.8 | −3.9 (−12.6 to 4.8) | 0.99 (0.91–1.07) | 0.811 | 0.011 |
| **Diabetes, (self-report, HbA1c ≥6.5% or medication)** |  |  |  |  |  |  |
| **Sex:** Men | 8.8 | 14.0 | 5.1 (2.7 to 7.5) | 1.31 (1.22–1.41) | <0.001 | 0.095 |
| Women | 7.7 | 11.6 | 3.9 (1.5 to 6.3) | 1.21 (1.12–1.32) | <0.001 | 0.095 |
| **Age group:** 20–39 | 2.0 | 3.6 | 1.6 (0.4 to 2.9) | 1.25 (1.06–1.46) | 0.007 | 0.061 |
| 40–59 | 8.4 | 15.3 | 6.8 (4.1 to 9.6) | 1.35 (1.24–1.47) | <0.001 | 0.061 |
| 60+ | 18.6 | 24.3 | 5.7 (1.5 to 9.9) | 1.20 (1.12–1.28) | <0.001 | 0.061 |
| **Race and Hispanic origin:** Non-Hispanic White | 6.5 | 10.4 | 3.9 (1.8 to 6.0) | 1.27 (1.18–1.38) | <0.001 | 0.016 |
| Non-Hispanic Black | 14.7 | 21.4 | 6.7 (2.9 to 10.4) | 1.17 (1.08–1.26) | <0.001 | 0.016 |
| Mexican American | 12.6 | 19.6 | 7.0 (2.1 to 11.9) | 1.33 (1.19–1.50) | <0.001 | 0.016 |
| Non-Hispanic other or multiracial | 15.5 | 14.1 | −1.3 (−10.7 to 8.0) | 1.01 (0.83–1.23) | 0.886 | 0.016 |
| All Hispanic (2007–2023 only) † | 14.2 | 16.1 | 2.0 (−2.3 to 6.3) | 1.10 (0.90–1.34) | 0.350 |  |
| **Family income-to-poverty ratio:** ≤1.30 | 12.0 | 17.6 | 5.7 (0.5 to 10.8) | 1.28 (1.14–1.42) | <0.001 | 0.958 |
| 1.31–3.50 | 10.2 | 15.2 | 5.0 (2.2 to 7.8) | 1.30 (1.19–1.41) | <0.001 | 0.958 |
| >3.50 | 4.7 | 8.3 | 3.6 (1.3 to 5.9) | 1.30 (1.18–1.44) | <0.001 | 0.958 |
| **Education:** Less than high school | 12.4 | 21.8 | 9.4 (5.1 to 13.7) | 1.32 (1.21–1.44) | <0.001 | 0.859 |
| High school or GED | 8.2 | 14.0 | 5.8 (2.7 to 8.9) | 1.39 (1.25–1.54) | <0.001 | 0.859 |
| Some college or associate degree | 7.5 | 13.7 | 6.2 (2.7 to 9.6) | 1.35 (1.23–1.49) | <0.001 | 0.859 |
| College graduate or above | 3.1 | 8.0 | 4.8 (2.6 to 7.1) | 1.31 (1.15–1.49) | <0.001 | 0.859 |
| **Current smoking** |  |  |  |  |  |  |
| **Sex:** Men | 27.1 | 18.3 | −8.8 (−13.3 to −4.3) | 0.76 (0.71–0.82) | <0.001 | 0.512 |
| Women | 21.7 | 11.5 | −10.2 (−14.1 to −6.4) | 0.75 (0.69–0.81) | <0.001 | 0.512 |
| **Age group:** 20–39 | 31.3 | 12.9 | −18.4 (−24.1 to −12.7) | 0.64 (0.58–0.71) | <0.001 | <0.001 |
| 40–59 | 24.4 | 18.3 | −6.1 (−11.1 to −1.1) | 0.80 (0.74–0.87) | <0.001 | <0.001 |
| 60+ | 12.7 | 12.9 | 0.2 (−3.0 to 3.4) | 0.99 (0.90–1.08) | 0.773 | <0.001 |
| **Race and Hispanic origin:** Non-Hispanic White | 25.4 | 15.4 | −10.0 (−14.7 to −5.4) | 0.77 (0.71–0.84) | <0.001 | 0.011 |
| Non-Hispanic Black | 25.7 | 18.7 | −6.9 (−14.2 to 0.3) | 0.85 (0.76–0.95) | 0.005 | 0.011 |
| Mexican American | 19.3 | 10.1 | −9.2 (−14.1 to −4.4) | 0.66 (0.59–0.74) | <0.001 | 0.011 |
| Non-Hispanic other or multiracial | 24.1 | 15.2 | −8.9 (−15.7 to −2.2) | 0.77 (0.64–0.91) | 0.003 | 0.011 |
| All Hispanic (2007–2023 only) † | 18.3 | 11.3 | −7.0 (−11.6 to −2.4) | 0.66 (0.54–0.81) | <0.001 |  |
| **Family income-to-poverty ratio:** ≤1.30 | 33.0 | 27.2 | −5.8 (−12.1 to 0.5) | 0.87 (0.79–0.95) | 0.003 | <0.001 |
| 1.31–3.50 | 27.2 | 18.0 | −9.2 (−14.5 to −4.0) | 0.77 (0.71–0.84) | <0.001 | <0.001 |
| >3.50 | 17.1 | 7.3 | −9.8 (−13.2 to −6.4) | 0.62 (0.56–0.69) | <0.001 | <0.001 |
| **Education:** Less than high school | 33.9 | 26.7 | −7.3 (−15.2 to 0.7) | 0.81 (0.74–0.89) | <0.001 | <0.001 |
| High school or GED | 31.6 | 24.6 | −7.0 (−13.2 to −0.9) | 0.85 (0.79–0.92) | <0.001 | <0.001 |
| Some college or associate degree | 21.7 | 15.6 | −6.0 (−11.0 to −1.1) | 0.84 (0.78–0.91) | <0.001 | <0.001 |
| College graduate or above | 11.4 | 3.8 | −7.6 (−10.9 to −4.3) | 0.64 (0.57–0.73) | <0.001 | <0.001 |
| **Drinks at least once a month (past 12 months)** |  |  |  |  |  |  |
| **Sex:** Men | 62.2 | 62.2 | 0.1 (−6.1 to 6.3) | 1.01 (0.93–1.10) | 0.818 | <0.001 |
| Women | 40.7 | 49.4 | 8.7 (1.6 to 15.8) | 1.22 (1.12–1.32) | <0.001 | <0.001 |
| **Age group:** 20–39 | 59.5 | 60.9 | 1.4 (−4.4 to 7.2) | 1.04 (0.95–1.14) | 0.393 | <0.001 |
| 40–59 | 52.5 | 55.6 | 3.1 (−4.8 to 11.0) | 1.09 (1.01–1.19) | 0.031 | <0.001 |
| 60+ | 35.6 | 47.3 | 11.8 (3.5 to 20.1) | 1.24 (1.12–1.37) | <0.001 | <0.001 |
| **Race and Hispanic origin:** Non-Hispanic White | 54.4 | 60.3 | 5.8 (−2.8 to 14.4) | 1.17 (1.05–1.30) | 0.005 | 0.017 |
| Non-Hispanic Black | 37.7 | 48.0 | 10.4 (3.4 to 17.3) | 1.24 (1.15–1.34) | <0.001 | 0.017 |
| Mexican American | 44.7 | 50.5 | 5.8 (−0.3 to 11.9) | 1.07 (1.00–1.15) | 0.057 | 0.017 |
| Non-Hispanic other or multiracial | 43.8 | 43.6 | −0.1 (−9.9 to 9.6) | 1.06 (0.94–1.20) | 0.347 | 0.017 |
| All Hispanic (2007–2023 only) † | 45.5 | 51.5 | 6.0 (−1.4 to 13.4) | 1.19 (1.01–1.39) | 0.032 |  |
| **Family income-to-poverty ratio:** ≤1.30 | 38.4 | 44.5 | 6.1 (−0.5 to 12.7) | 1.15 (1.06–1.24) | <0.001 | 0.648 |
| 1.31–3.50 | 44.3 | 50.2 | 5.9 (−2.8 to 14.6) | 1.10 (1.00–1.20) | 0.053 | 0.648 |
| >3.50 | 64.1 | 65.5 | 1.4 (−4.1 to 6.9) | 1.10 (1.01–1.20) | 0.023 | 0.648 |
| **Education:** Less than high school | 39.5 | 42.1 | 2.6 (−4.7 to 9.9) | 1.03 (0.94–1.12) | 0.541 | 0.465 |
| High school or GED | 47.8 | 48.2 | 0.3 (−5.3 to 6.0) | 1.01 (0.93–1.10) | 0.779 | 0.465 |
| Some college or associate degree | 54.7 | 53.7 | −1.1 (−8.8 to 6.6) | 1.05 (0.96–1.15) | 0.313 | 0.465 |
| College graduate or above | 65.1 | 66.0 | 0.9 (−8.4 to 10.3) | 1.07 (0.96–1.19) | 0.207 | 0.465 |
| **CKD (eGFR <60, UACR ≥30, weak/failing kidneys or dialysis)** |  |  |  |  |  |  |
| **Sex:** Men | 13.0 | 12.8 | −0.2 (−2.8 to 2.4) | 0.99 (0.92–1.07) | 0.855 | 0.312 |
| Women | 16.4 | 14.3 | −2.0 (−4.7 to 0.6) | 0.96 (0.91–1.02) | 0.235 | 0.312 |
| **Age group:** 20–39 | 7.1 | 5.8 | −1.4 (−4.3 to 1.5) | 1.02 (0.90–1.15) | 0.756 | <0.001 |
| 40–59 | 11.7 | 12.0 | 0.3 (−2.5 to 3.0) | 1.09 (1.00–1.18) | 0.052 | <0.001 |
| 60+ | 32.5 | 29.4 | −3.1 (−7.2 to 0.9) | 0.90 (0.85–0.95) | <0.001 | <0.001 |
| **Race and Hispanic origin:** Non-Hispanic White | 13.0 | 12.2 | −0.7 (−2.9 to 1.5) | 0.98 (0.92–1.04) | 0.547 | 0.808 |
| Non-Hispanic Black | 21.4 | 19.1 | −2.2 (−7.6 to 3.2) | 0.94 (0.85–1.05) | 0.261 | 0.808 |
| Mexican American | 15.9 | 13.0 | −2.9 (−8.5 to 2.7) | 0.99 (0.87–1.12) | 0.848 | 0.808 |
| Non-Hispanic other or multiracial | 21.0 | 15.6 | −5.4 (−16.3 to 5.5) | 0.94 (0.76–1.15) | 0.541 | 0.808 |
| All Hispanic (2007–2023 only) † | 16.6 | 12.2 | −4.4 (−7.3 to −1.5) | 0.84 (0.71–0.99) | 0.042 |  |
| **Family income-to-poverty ratio:** ≤1.30 | 21.6 | 17.5 | −4.0 (−8.7 to 0.6) | 0.94 (0.86–1.03) | 0.186 | 0.056 |
| 1.31–3.50 | 14.3 | 15.1 | 0.8 (−2.1 to 3.7) | 1.05 (0.98–1.12) | 0.184 | 0.056 |
| >3.50 | 10.7 | 9.6 | −1.1 (−3.9 to 1.7) | 0.96 (0.88–1.05) | 0.369 | 0.056 |
| **Education:** Less than high school | 19.0 | 21.9 | 2.8 (−0.9 to 6.5) | 1.02 (0.96–1.09) | 0.499 | 0.762 |
| High school or GED | 13.8 | 15.7 | 1.9 (−1.1 to 4.8) | 1.07 (0.98–1.16) | 0.123 | 0.762 |
| Some college or associate degree | 14.3 | 12.9 | −1.3 (−4.7 to 2.1) | 1.01 (0.93–1.10) | 0.755 | 0.762 |
| College graduate or above | 10.8 | 9.8 | −1.0 (−5.1 to 3.1) | 1.01 (0.91–1.13) | 0.797 | 0.762 |
| **Prior CVD (any)** |  |  |  |  |  |  |
| **Sex:** Men | 10.4 | 9.7 | −0.8 (−3.3 to 1.8) | 0.96 (0.88–1.04) | 0.341 | 0.387 |
| Women | 7.2 | 6.9 | −0.3 (−2.1 to 1.4) | 0.93 (0.86–1.02) | 0.134 | 0.387 |
| **Age group:** 20–39 | 0.7 | 1.0 | 0.3 (−0.6 to 1.2) | 1.09 (0.87–1.36) | 0.470 | 0.148 |
| 40–59 | 7.8 | 7.2 | −0.6 (−3.2 to 2.0) | 1.02 (0.89–1.16) | 0.783 | 0.148 |
| 60+ | 23.6 | 22.0 | −1.6 (−5.9 to 2.6) | 0.91 (0.85–0.98) | 0.009 | 0.148 |
| **Race and Hispanic origin:** Non-Hispanic White | 8.9 | 8.3 | −0.6 (−2.6 to 1.4) | 0.95 (0.87–1.03) | 0.197 | 0.962 |
| Non-Hispanic Black | 8.5 | 8.7 | 0.2 (−2.8 to 3.3) | 0.95 (0.85–1.06) | 0.342 | 0.962 |
| Mexican American | 6.6 | 6.2 | −0.4 (−2.6 to 1.8) | 0.92 (0.78–1.07) | 0.278 | 0.962 |
| Non-Hispanic other or multiracial **‡** | 9.0 | 7.7 | −1.4 (−6.8 to 4.1) | 0.91 (0.72–1.16) | 0.448 | 0.962 |
| All Hispanic (2007–2023 only) **†** | 6.5 | 7.7 | 1.2 (−0.7 to 3.1) | 1.07 (0.87–1.31) | 0.520 |  |
| **Family income-to-poverty ratio:** ≤1.30 | 13.1 | 12.7 | −0.4 (−3.2 to 2.5) | 0.94 (0.86–1.03) | 0.183 | 0.005 |
| 1.31–3.50 | 8.5 | 8.6 | 0.1 (−2.4 to 2.6) | 1.05 (0.96–1.16) | 0.279 | 0.005 |
| >3.50 | 7.4 | 5.7 | −1.8 (−3.0 to −0.5) | 0.85 (0.76–0.94) | 0.002 | 0.005 |
| **Education:** Less than high school | 10.6 | 13.6 | 3.1 (−0.1 to 6.3) | 0.99 (0.90–1.10) | 0.889 | 0.340 |
| High school or GED | 9.4 | 10.0 | 0.6 (−1.8 to 3.0) | 1.07 (0.97–1.19) | 0.176 | 0.340 |
| Some college or associate degree | 8.7 | 8.6 | −0.1 (−2.8 to 2.6) | 1.02 (0.92–1.12) | 0.757 | 0.340 |
| College graduate or above | 5.5 | 4.6 | −0.9 (−3.3 to 1.4) | 0.90 (0.80–1.01) | 0.071 | 0.340 |

**Note:** Estimates are age-standardized prevalence percentages, except for age-stratified analyses, which are presented as age-specific estimates. Changes represent absolute percentage-point differences between the first and last survey cycles, and odds ratios (ORs) describe trends per decade. P values for trend assess temporal changes within each subgroup, whereas P values for interaction assess whether trends differ across subgroups. ‡ Does not meet the NCHS data presentation standards (relative CI width >130%); interpret with caution. † Analyses of all Hispanic adults were restricted to 2007–2008 through 2021–2023.

**Abbreviations:** BMI, body mass index; CI, confidence interval; CKD, chronic kidney disease; CVD, cardiovascular disease; eGFR, estimated glomerular filtration rate; GED, General Educational Development; HbA1c, glycated hemoglobin; HDL, high-density lipoprotein; OR, odds ratio; TC, total cholesterol; UACR, urine albumin-to-creatinine ratio.

**eTable 5. Sensitivity analyses**

| **Analysis** | **First cycle** | **First estimate** | **Last cycle** | **Last estimate** | **Change (95% CI)** | **Effect per decade (95% CI)** | **P for trend** | **P for nonlinearity** |
| --- | --- | --- | --- | --- | --- | --- | --- | --- |
| **Hypertension Definition** | | | | | | | | |
| Hypertension with ≥140/90 mm Hg threshold | 1999–2000 | 34.3 | 2021–2023 | 34.3 | 0.0 (−3.8 to 3.8) | OR 1.01 (0.95–1.07) | 0.749 | 0.061 |
| **Hypertension Measurement change** | | | | | | | | |
| Hypertension, 1999–2016 only (single BP device) | 1999–2000 | 50.6 | 2015–2016 | 48.1 | −2.5 (−6.1 to 1.0) | OR 0.93 (0.86–1.00) | 0.042 | 0.057 |
| Mean SBP, 1999–2016 only (single BP device) | 1999–2000 | 124.2 | 2015–2016 | 123.0 | −1.2 (−2.8 to 0.4) | Δ −1.53 (−2.18 to −0.89) | <0.001 | <0.001 |
| **HDL assay** | | | | | | | | |
| Dyslipidemia without the low-HDL criterion | 1999–2000 | 33.6 | 2021–2023 | 38.2 | 4.5 (2.0 to 7.1) | OR 1.10 (1.06–1.14) | <0.001 | 0.089 |
| Dyslipidemia, primary, 2007–2023 only | 2007–2008 | 55.8 | 2021–2023 | 53.4 | −2.3 (−6.3 to 1.7) | OR 0.94 (0.86–1.02) | 0.129 | 0.667 |
| Low HDL, 2007–2023 only | 2007–2008 | 34.0 | 2021–2023 | 26.7 | −7.3 (−11.3 to −3.4) | OR 0.80 (0.73–0.88) | <0.001 | 0.715 |
| Mean HDL, 2007–2023 only | 2007–2008 | 51.8 | 2021–2023 | 53.3 | 1.5 (0.1 to 2.9) | Δ 1.05 (0.32–1.78) | 0.005 | 0.011 |
| Mean non-HDL cholesterol, 2007–2023 only | 2007–2008 | 144.9 | 2021–2023 | 134.8 | −10.0 (−12.7 to −7.4) | Δ −8.08 (−9.65 to −6.52) | <0.001 | <0.001 |
| **Medication harmonization** | | | | | | | | |
| Dyslipidemia, lipid medication as asked | 1999–2000 | 57.5 | 2021–2023 | 55.0 | −2.5 (−6.7 to 1.7) | OR 1.03 (0.98–1.07) | 0.254 | 0.707 |
| Diabetes, glucose-lowering medication as asked | 1999–2000 | 8.2 | 2021–2023 | 13.5 | 5.3 (3.4 to 7.1) | OR 1.30 (1.23–1.38) | <0.001 | 0.154 |
| **Complete case** | | | | | | | | |
| Obesity, complete case (BMI and waist measured) | 1999–2000 | 47.5 | 2021–2023 | 57.7 | 10.2 (4.7 to 15.7) | OR 1.23 (1.16–1.31) | <0.001 | 0.031 |
| Hypertension, complete case | 1999–2000 | 51.4 | 2021–2023 | 48.7 | −2.7 (−6.5 to 1.1) | OR 0.96 (0.91–1.02) | 0.187 | 0.059 |
| Dyslipidemia, complete case | 1999–2000 | 59.5 | 2021–2023 | 54.2 | −5.4 (−9.5 to −1.2) | OR 0.97 (0.92–1.01) | 0.129 | 0.863 |
| Diabetes, complete case | 1999–2000 | 8.1 | 2021–2023 | 12.7 | 4.6 (2.7 to 6.5) | OR 1.27 (1.20–1.35) | <0.001 | 0.156 |
| CKD, complete case | 1999–2000 | 14.5 | 2021–2023 | 13.4 | −1.2 (−3.3 to 0.9) | OR 0.98 (0.93–1.03) | 0.395 | 0.693 |
| **Asian BMI cutoffs** | | | | | | | | |
| Obesity with Asian BMI cutoff ≥27.5 (2011–2023) | 2011–2012 | 54.8 | 2021–2023 | 57.8 | 3.0 (−1.7 to 7.7) | OR 1.13 (0.96–1.32) | 0.130 | 0.043 |
| Obesity, standard cutoffs (2011–2023, comparator) | 2011–2012 | 54.6 | 2021–2023 | 57.2 | 2.7 (−2.1 to 7.5) | OR 1.11 (0.95–1.31) | 0.193 | 0.049 |
| Overweight with Asian cutoffs 23–27.4 (2011–2023) | 2011–2012 | 34.2 | 2021–2023 | 32.1 | −2.1 (−5.3 to 1.0) | OR 0.91 (0.82–1.01) | 0.083 | 0.128 |
| **Pandemic gap** | | | | | | | | |
| Obesity, pre-pandemic cycles only (1999-March 2020) | 1999–2000 | 47.4 | 2017–Mar 2020 | 59.5 | 12.0 (7.3 to 16.8) | OR 1.28 (1.20–1.37) | <0.001 | 0.905 |
| Hypertension, pre-pandemic cycles only | 1999–2000 | 50.6 | 2017–Mar 2020 | 48.3 | −2.2 (−6.1 to 1.6) | OR 0.96 (0.89–1.02) | 0.177 | 0.055 |
| Dyslipidemia, pre-pandemic cycles only | 1999–2000 | 57.5 | 2017–Mar 2020 | 53.2 | −4.3 (−8.2 to −0.5) | OR 0.98 (0.93–1.04) | 0.575 | 0.954 |
| Diabetes, pre-pandemic cycles only | 1999–2000 | 8.2 | 2017–Mar 2020 | 12.7 | 4.5 (2.9 to 6.1) | OR 1.30 (1.22–1.40) | <0.001 | 0.884 |
| CKD, pre-pandemic cycles only | 1999–2000 | 14.7 | 2017–Mar 2020 | 14.1 | −0.7 (−2.7 to 1.4) | OR 0.98 (0.92–1.04) | 0.540 | 0.280 |
| Current smoking, pre-pandemic cycles only | 1999–2000 | 24.4 | 2017–Mar 2020 | 17.4 | −7.0 (−10.5 to −3.5) | OR 0.77 (0.71–0.83) | <0.001 | 0.227 |
| Monthly drinking, pre-pandemic cycles only | 1999–2000 | 51.3 | 2017–Mar 2020 | 53.9 | 2.5 (−3.5 to 8.6) | OR 1.13 (1.03–1.24) | 0.009 | 0.055 |
| Prior CVD, pre-pandemic cycles only | 1999–2000 | 8.7 | 2017–Mar 2020 | 8.8 | 0.1 (−1.5 to 1.7) | OR 0.95 (0.87–1.03) | 0.188 | 0.189 |

**Note:** Estimates are age-standardized. Changes represent differences between the first and last survey cycles, expressed in percentage points for prevalence outcomes and original units for continuous measures. Trends per decade are reported as odds ratios (ORs) for binary outcomes and regression coefficients (Δ) for continuous outcomes. P values for trend and nonlinearity correspond to the linear and quadratic time terms, respectively. Sensitivity analyses examined alternative outcome definitions, measurement changes, medication harmonization, complete-case restrictions, Asian-specific BMI thresholds, and trends restricted to the pre-pandemic period.

**Abbreviations:** BMI, body mass index; BP, blood pressure; CI, confidence interval; CKD, chronic kidney disease; CVD, cardiovascular disease; HDL, high-density lipoprotein; OR, odds ratio; SBP, systolic blood pressure.

**eTable 6. Joinpoint regression of age-standardized estimates**

| **Outcome** | **Joinpoint (cycle)** | **P (permutation, 0 vs 1 joinpoint)** | **Segment** | **Slope per year (95% CI)** | **P slope** |
| --- | --- | --- | --- | --- | --- |
| **Adiposity** |  |  |  |  |  |
| Obesity, (BMI ≥30 or waist >102/88 cm) | None | 0.177 | 1999–2000 to 2021–2023 | 0.528 (0.391 to 0.665) | <0.001 |
| Obesity by BMI ≥30 alone | None | 0.807 | 1999–2000 to 2021–2023 | 0.594 (0.476 to 0.711) | <0.001 |
| Obesity class III (BMI ≥40) | 2007–2008 | 0.036 | 1999–2000 to 2007–2008 | 0.124 (0.032 to 0.216) | 0.016 |
|  |  |  | 2007–2008 to 2021–2023 | 0.288 (0.227 to 0.348) | <0.001 |
| **Hypertension** |  |  |  |  |  |
| Hypertension, (≥130/80, medication or self-report) | 2007–2008 | 0.011 | 1999–2000 to 2007–2008 | −0.450 (−0.676 to −0.223) | 0.002 |
|  |  |  | 2007–2008 to 2021–2023 | 0.100 (−0.014 to 0.215) | 0.076 |
| **Dyslipidemia** |  |  |  |  |  |
| Dyslipidemia, (TC ≥240, low HDL, medication or self-report) | None | 0.721 | 1999–2000 to 2021–2023 | 0.033 (−0.140 to 0.207) | 0.677 |
| Dyslipidemia without the low-HDL criterion | None | 0.336 | 1999–2000 to 2021–2023 | 0.181 (0.033 to 0.329) | 0.022 |
| **Diabetes and kidney disease** |  |  |  |  |  |
| Diabetes, (self-report, HbA1c ≥6.5% or medication) | None | 0.793 | 1999–2000 to 2021–2023 | 0.218 (0.180 to 0.256) | <0.001 |
| CKD (eGFR <60, UACR ≥30, weak/failing kidneys or dialysis) | None | 0.582 | 1999–2000 to 2021–2023 | −0.019 (−0.103 to 0.064) | 0.616 |
| **Behavioral risk factors and prior CVD** |  |  |  |  |  |
| Current smoking | None | 0.062 | 1999–2000 to 2021–2023 | −0.471 (−0.575 to −0.367) | <0.001 |
| Drinks at least once a month (past 12 months) | 2011–2012 | 0.030 | 1999–2000 to 2011–2012 | 0.697 (0.247 to 1.146) | 0.008 |
|  |  |  | 2011–2012 to 2021–2023 | −0.304 (−0.706 to 0.097) | 0.116 |
| Prior CVD (any) | None | 0.222 | 1999–2000 to 2021–2023 | −0.031 (−0.084 to 0.023) | 0.230 |
| **Hypertension awareness, treatment, and control** |  |  |  |  |  |
| Hypertension: aware | 2013–2014 | 0.005 | 1999–2000 to 2013–2014 | 1.161 (0.691 to 1.630) | <0.001 |
|  |  |  | 2013–2014 to 2021–2023 | −1.158 (−2.017 to −0.300) | 0.015 |
| Hypertension: treated | 2009–2010 | <0.001 | 1999–2000 to 2009–2010 | 1.130 (0.863 to 1.396) | <0.001 |
|  |  |  | 2009–2010 to 2021–2023 | −0.092 (−0.291 to 0.107) | 0.311 |
| Hypertension: controlled (<130/80) | 2013–2014 | <0.001 | 1999–2000 to 2013–2014 | 1.163 (0.879 to 1.446) | <0.001 |
|  |  |  | 2013–2014 to 2021–2023 | −1.169 (−1.638 to −0.700) | <0.001 |
| **Dyslipidemia awareness, treatment, and control** |  |  |  |  |  |
| Dyslipidemia: aware | None | 0.316 | 1999–2000 to 2021–2023 | 0.549 (0.299 to 0.799) | <0.001 |
| Dyslipidemia: treated | 2005–2006 | 0.008 | 1999–2000 to 2005–2006 | 1.579 (0.931 to 2.227) | <0.001 |
|  |  |  | 2005–2006 to 2021–2023 | 0.176 (−0.067 to 0.418) | 0.131 |
| Dyslipidemia: LDL <100 mg/dL (fasting) | 2005–2006 | 0.004 | 1999–2000 to 2005–2006 | 1.867 (1.292 to 2.441) | <0.001 |
|  |  |  | 2005–2006 to 2021–2023 | 0.333 (0.097 to 0.569) | 0.013 |
| **Diabetes awareness, treatment, and control** |  |  |  |  |  |
| Diabetes: aware | None | 0.806 | 1999–2000 to 2021–2023 | 0.092 (−0.293 to 0.476) | 0.604 |
| Diabetes: treated | None | 0.456 | 1999–2000 to 2021–2023 | 0.087 (−0.375 to 0.549) | 0.680 |
| Diabetes: HbA1c <7% | 2003–2004 | 0.023 | 1999–2000 to 2003–2004 | 4.714 (1.559 to 7.868) | 0.010 |
|  |  |  | 2003–2004 to 2021–2023 | −0.281 (−0.786 to 0.224) | 0.230 |
| **Mean clinical and laboratory measures** |  |  |  |  |  |
| Mean BMI, kg/m2 | None | 0.555 | 1999–2000 to 2021–2023 | 0.092 (0.076 to 0.108) | <0.001 |
| Mean waist circumference, cm | None | 0.159 | 1999–2000 to 2021–2023 | 0.234 (0.190 to 0.277) | <0.001 |
| Mean SBP, mm Hg | None | 0.275 | 1999–2000 to 2021–2023 | −0.133 (−0.218 to −0.047) | 0.007 |
| Mean total cholesterol, mg/dL | None | 0.241 | 1999–2000 to 2021–2023 | −0.821 (−1.013 to −0.630) | <0.001 |
| Mean HDL cholesterol, mg/dL | None | 0.057 | 1999–2000 to 2021–2023 | 0.063 (−0.044 to 0.171) | 0.214 |
| Mean non-HDL cholesterol, mg/dL | None | 0.197 | 1999–2000 to 2021–2023 | −0.865 (−1.089 to −0.641) | <0.001 |
| Mean LDL cholesterol, mg/dL (fasting) | 2003–2004 | 0.013 | 1999–2000 to 2003–2004 | −2.239 (−3.260 to −1.218) | 0.001 |
|  |  |  | 2003–2004 to 2021–2023 | −0.370 (−0.546 to −0.193) | 0.002 |
| Mean HbA1c, % | None | 0.483 | 1999–2000 to 2021–2023 | 0.011 (0.007 to 0.015) | <0.001 |
| **Predicted 10-year cardiovascular risk** |  |  |  |  |  |
| 10-year total CVD risk, mean % | None | 0.186 | 1999–2000 to 2021–2023 | −0.011 (−0.020 to −0.003) | 0.016 |
| 10-year ASCVD risk, mean % | None | 0.050 | 1999–2000 to 2021–2023 | −0.015 (−0.021 to −0.009) | <0.001 |
| 10-year heart failure risk, mean % | None | 0.400 | 1999–2000 to 2021–2023 | 0.009 (0.005 to 0.014) | 0.002 |

**Note:** Joinpoint regression was applied to age-standardized estimates using inverse-variance weights (1/SE²), with a maximum of one joinpoint allowed. A permutation test was used to compare models with and without a joinpoint. When P<.05, the joinpoint was retained; otherwise, a single linear trend was fitted across the study period. Slopes are reported as annual changes in prevalence (percentage points per year) or continuous measures (original units per year), with 95% confidence intervals. P values indicate whether each segment's slope differs significantly from zero. Survey-cycle labels identify the periods in which joinpoints were estimated.

**Abbreviations:** ASCVD, atherosclerotic cardiovascular disease; BMI, body mass index; CI, confidence interval; CKD, chronic kidney disease; CVD, cardiovascular disease; eGFR, estimated glomerular filtration rate; HbA1c, glycated hemoglobin; HDL, high-density lipoprotein; LDL, low-density lipoprotein; SBP, systolic blood pressure; SE, standard error; UACR, urine albumin-to-creatinine ratio.

**eTable 7. Obesity and Overweight Among Non-Hispanic Asian Adults Using Standard and Asian BMI Thresholds, 2011–2023**

| **Measure** | **2011–2012** | **2013–2014** | **2015–2016** | **2017–March 2020** | **2021–2023** |
| --- | --- | --- | --- | --- | --- |
| Obesity, Asian BMI threshold ≥27.5 kg/m² | 20.8 (17.3–24.6) | 23.5 (19.9–27.3) | 25.5 (20.1–31.6) | 32.3 (28.5–36.3) | 27.4 (19.9–36.0) |
| Obesity, standard BMI threshold ≥30 kg/m² | 10.8 (8.0–14.2) | 12.6 (9.9–15.7) | 12.7 (10.0–15.8) | 16.1 (13.5–18.9) | 13.1 (7.0–21.8) |
| Overweight, Asian BMI threshold 23.0–27.4 kg/m² | 40.3 (34.7–46.1) | 39.5 (34.8–44.3) | 39.9 (31.3–49.0) | 40.6 (36.8–44.6) | 38.4 (32.6–44.4) |
| Overweight, standard BMI threshold 25.0–29.9 kg/m² | 27.8 (24.3–31.6) | 29.1 (25.3–33.2) | 30.7 (26.8–34.7) | 38.0 (33.8–42.4) | 32.1 (24.9–40.1) |

**Note:** Values are prevalence percentages (95% CIs). Asian-specific BMI thresholds define overweight as 23.0–27.4 kg/m² and obesity as ≥27.5 kg/m²; standard thresholds define overweight as 25.0–29.9 kg/m² and obesity as ≥30 kg/m². Estimates are restricted to non-Hispanic Asian adults.

**Abbreviations:** BMI, body mass index; CI, confidence interval.

**eTable 8. Item missingness (%) among eligible adults, by cycle**

| **Item** | **1999–2000** | **2001–2002** | **2003–2004** | **2005–2006** | **2007–2008** | **2009–2010** | **2011–2012** | **2013–2014** | **2015–2016** | **2017–Mar 2020** | **2021–2023** |
| --- | --- | --- | --- | --- | --- | --- | --- | --- | --- | --- | --- |
| Alcohol frequency | 6.5 | 7.9 | 9.3 | 9.3 | 8.7 | 12.1 | 11.9 | 8.8 | 9.5 | 6.6 | 13.7 |
| BMI | 1.7 | 6.7 | 2 | 2.1 | 1.8 | 1.1 | 1.5 | 1.2 | 1.2 | 1.9 | 1.6 |
| Blood pressure | 3.5 | 5.2 | 7.2 | 5.4 | 4.8 | 4.6 | 4.6 | 3.4 | 2.9 | 10.5 | 3.3 |
| Creatinine | 7.3 | 6.6 | 5.9 | 6.6 | 6.8 | 6.2 | 8 | 4.6 | 5.8 | 7.7 | 6 |
| Diabetes question | 0.1 | 0 | 0 | 0.1 | 0.1 | 0.1 | 0.1 | 0.1 | 0.1 | 0.1 | 0 |
| Education | 0.3 | 0.2 | 0.2 | 0.2 | 0.1 | 0.2 | 0.1 | 0.1 | 0.1 | 0.2 | 0.1 |
| Fasting glucose | 0.4 | 0.2 | 0.6 | 0 | 0 | 0 | 0 | 0 | 0 | 0 | 0 |
| HDL | 7.4 | 6.7 | 5.5 | 6.2 | 6.5 | 6 | 7.6 | 4.4 | 5.7 | 7.2 | 5.1 |
| HbA1c | 5.4 | 5 | 4.6 | 5.7 | 6.1 | 5.1 | 5.5 | 3.5 | 4.9 | 5.3 | 0.4 |
| Hypertension questions | 1.2 | 1.1 | 1.1 | 0.3 | 0.1 | 0.2 | 0.2 | 0.1 | 0.1 | 0.1 | 0.1 |
| Income-to-poverty ratio | 14.3 | 7.2 | 5.8 | 4.7 | 9.3 | 9.7 | 8.5 | 7.7 | 10.3 | 14 | 13.1 |
| LDL (fasting) | 4.6 | 5.2 | 3.9 | 3.8 | 2.7 | 2.8 | 3.1 | 2.4 | 1.8 | 2.5 | 5.2 |
| Prior CVD questions | 0 | 0 | 0 | 0 | 0 | 0 | 0 | 0 | 0 | 0 | 0 |
| Smoking | 0.2 | 0.1 | 0.1 | 0.1 | 0.1 | 0 | 0.1 | 0 | 0.2 | 0 | 0.1 |
| Total cholesterol | 7.4 | 6.7 | 5.4 | 6.2 | 6.5 | 6 | 7.6 | 4.4 | 5.7 | 7.2 | 5.1 |
| Triglycerides (fasting) | 1.9 | 2 | 1.2 | 1.2 | 0.5 | 1.1 | 1.3 | 1.1 | 0.7 | 1.7 | 4.1 |
| UACR | 2.9 | 3.3 | 3.2 | 3.2 | 3.2 | 2.2 | 2.2 | 2.2 | 2.3 | 2.6 | 1.9 |
| Waist | 3.8 | 5.7 | 5.9 | 5.8 | 5.8 | 5.9 | 6.4 | 5.7 | 6.5 | 5.6 | 4.9 |

**Note:** Values represent the percentage of eligible adults with missing data for each item within the corresponding survey cycle. Missingness was assessed within the applicable analytic sample, including the fasting subsample for fasting glucose, LDL cholesterol, and triglycerides.

**Abbreviations:** BMI, body mass index; CVD, cardiovascular disease; HbA1c, glycated hemoglobin; HDL, high-density lipoprotein; LDL, low-density lipoprotein; NHANES, National Health and Nutrition Examination Survey; UACR, urine albumin-to-creatinine ratio.

**eTable 9. Nonresponse Analyses for Items with More Than 10% Missing Data**

| **Item** | **Survey cycle** | **Missing (%)** | **Outcome assessed** | **Unadjusted estimate (%)** | **Nonresponse-adjusted estimate (%)** | **Respondents: ≥60 y / women / NH Black / Mexican American (%)** | **Nonrespondents: ≥60 y / women / NH Black / Mexican American (%)** |
| --- | --- | --- | --- | --- | --- | --- | --- |
| Blood pressure | 2017–March 2020 | 10.5 | Measured blood pressure ≥130/80 mm Hg | 36.9 | 36.9 | 30.2 / 50.8 / 11.2 / 7.7 | 28.8 / 58.3 / 13.5 / 14.5 |
| Alcohol frequency | 2009–2010 | 12.1 | Monthly drinking | 57.3 | 56.8 | 25.5 / 50.0 / 10.9 / 8.4 | 21.2 / 60.5 / 14.9 / 8.9 |
| Alcohol frequency | 2011–2012 | 11.9 | Monthly drinking | 58.8 | 58.4 | 27.6 / 50.5 / 11.0 / 7.2 | 12.9 / 60.6 / 15.8 / 12.1 |
| Alcohol frequency | 2021–2023 | 13.7 | Monthly drinking | 55.8 | 55.4 | 31.8 / 50.3 / 10.5 / 6.7 | 28.4 / 56.5 / 13.7 / 9.9 |
| Income-to-poverty ratio | 1999–2000 | 14.3 | — | NA | NA | 22.6 / 50.9 / 10.2 / 6.9 | 35.3 / 52.7 / 15.1 / 10.0 |
| Income-to-poverty ratio | 2015–2016 | 10.3 | — | NA | NA | 27.3 / 51.4 / 11.0 / 8.4 | 40.0 / 50.9 / 15.3 / 13.5 |
| Income-to-poverty ratio | 2017–March 2020 | 14.0 | — | NA | NA | 30.1 / 51.4 / 11.0 / 7.8 | 30.0 / 51.5 / 15.0 / 11.7 |
| Income-to-poverty ratio | 2021–2023 | 13.1 | — | NA | NA | 31.4 / 51.1 / 10.2 / 6.8 | 31.5 / 51.1 / 15.5 / 8.9 |

**Note:** Missingness and respondent characteristics are reported as survey-weighted percentages. Respondents and nonrespondents were compared by age (≥60 years), sex (women), non-Hispanic Black race, and Mexican American ethnicity. For blood pressure and alcohol frequency, age-standardized prevalence estimates based on the original survey weights were compared with estimates using weights adjusted for item nonresponse within age, sex, and race/Hispanic-origin groups. Because the income-to-poverty ratio was used only for subgroup analyses, outcome-specific nonresponse-adjusted estimates were not calculated for this variable.

**Abbreviations:** NA, not applicable; NH, non-Hispanic.

**eFigure 1. Selection of the analytic sample**

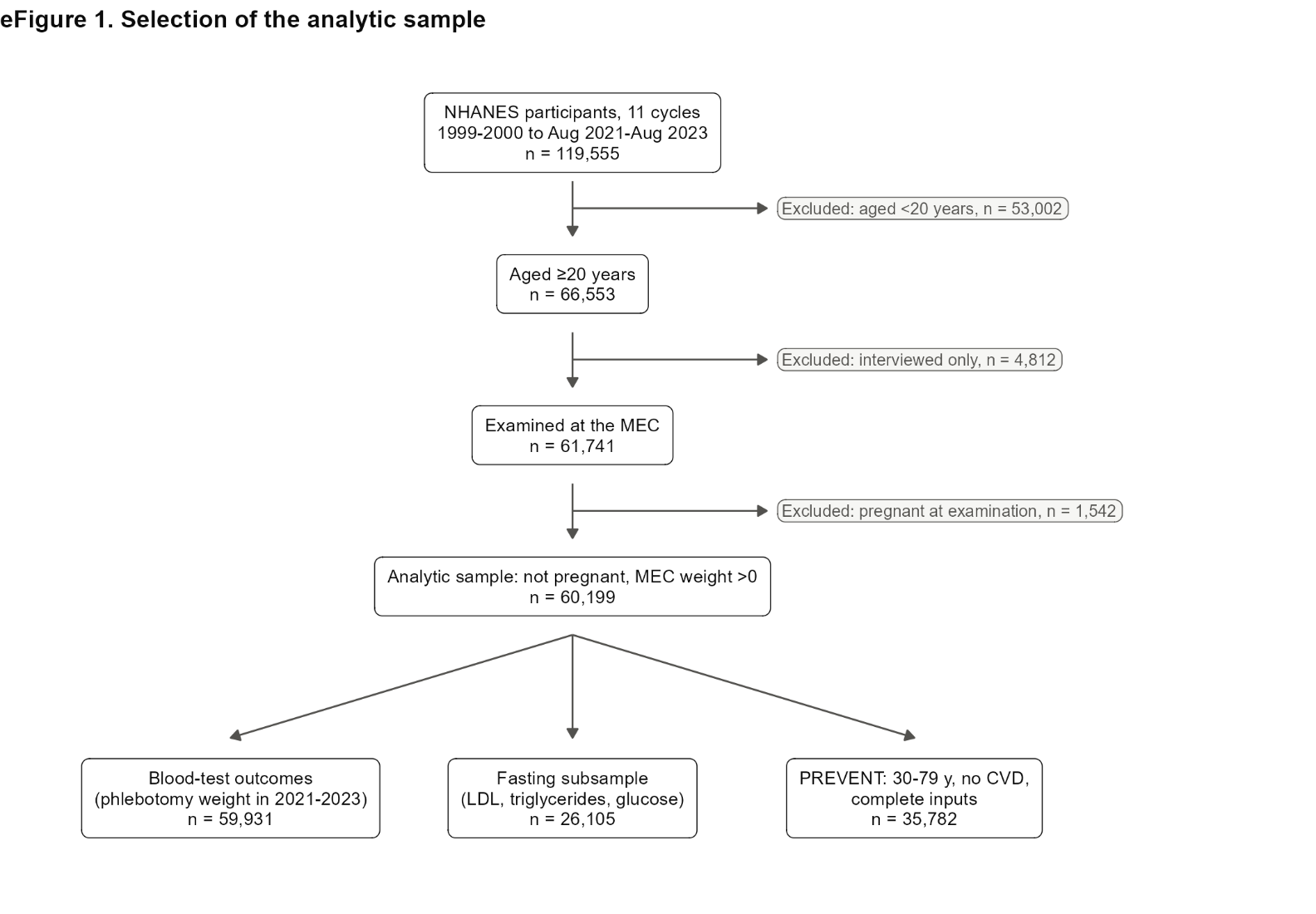

**eFigure 2. Weight status by BMI class**

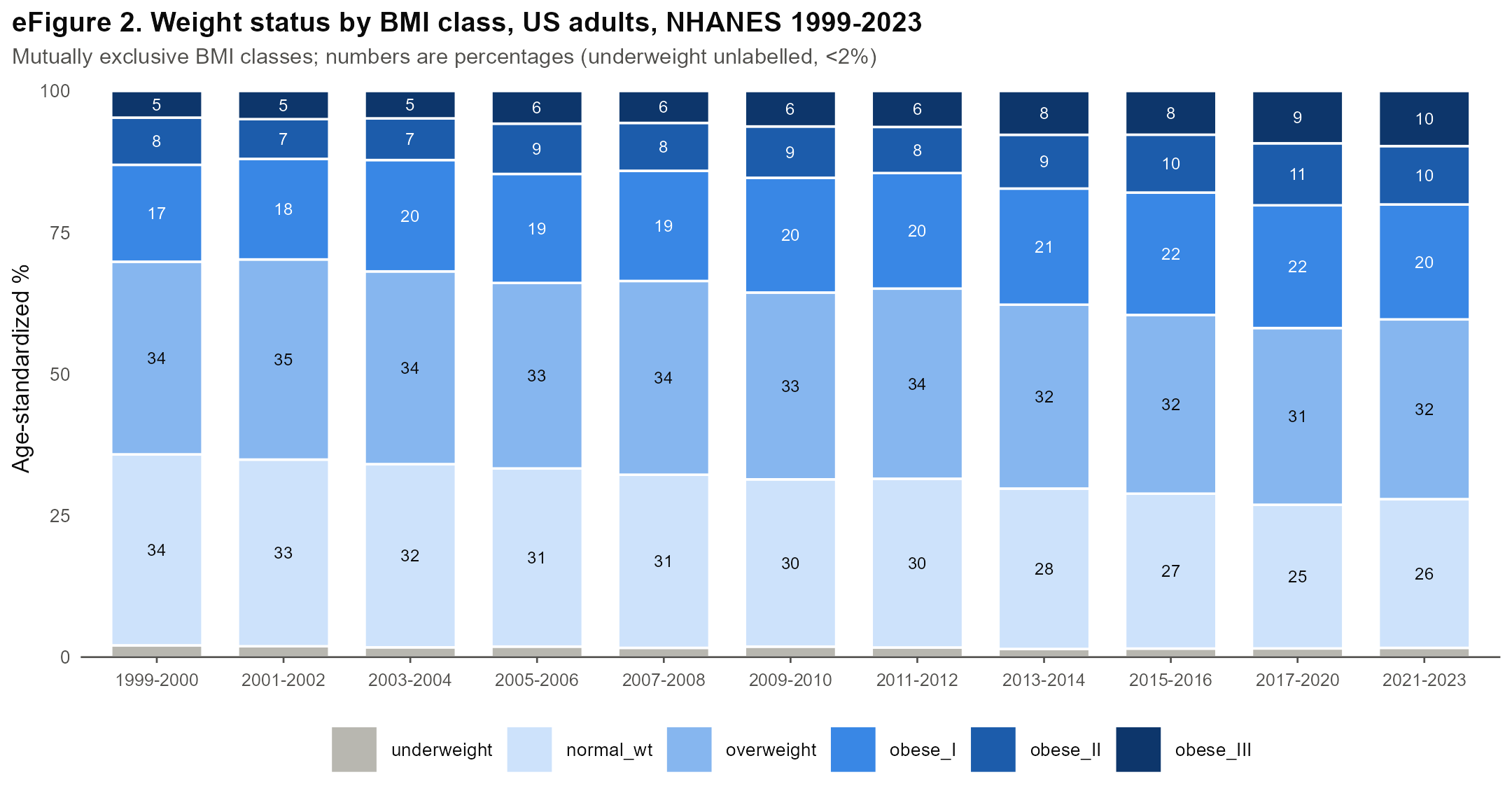

**eFigure 3. Mean levels of continuous risk factors**

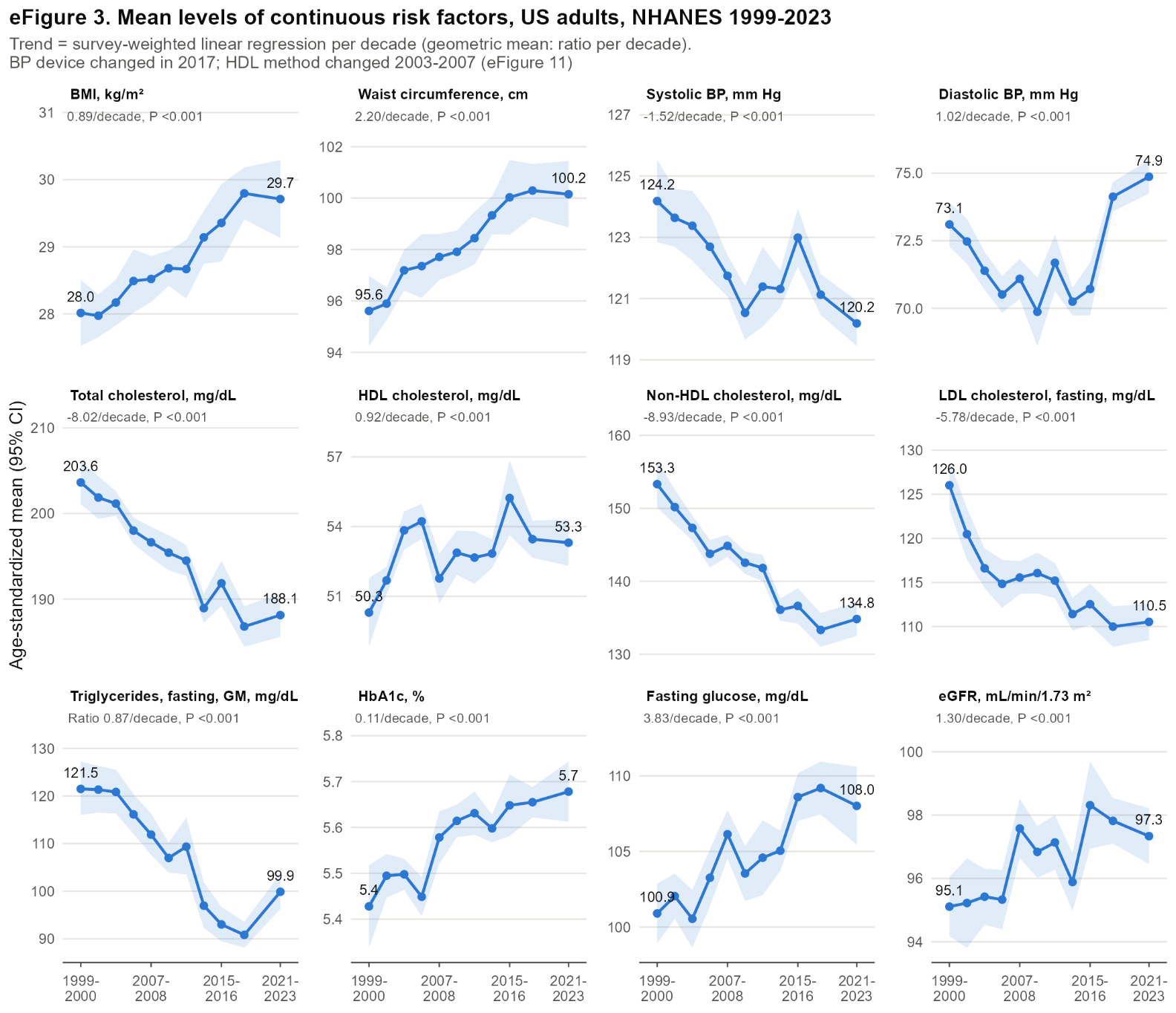

**eFigure 4. Components of the composite risk-factor definitions**

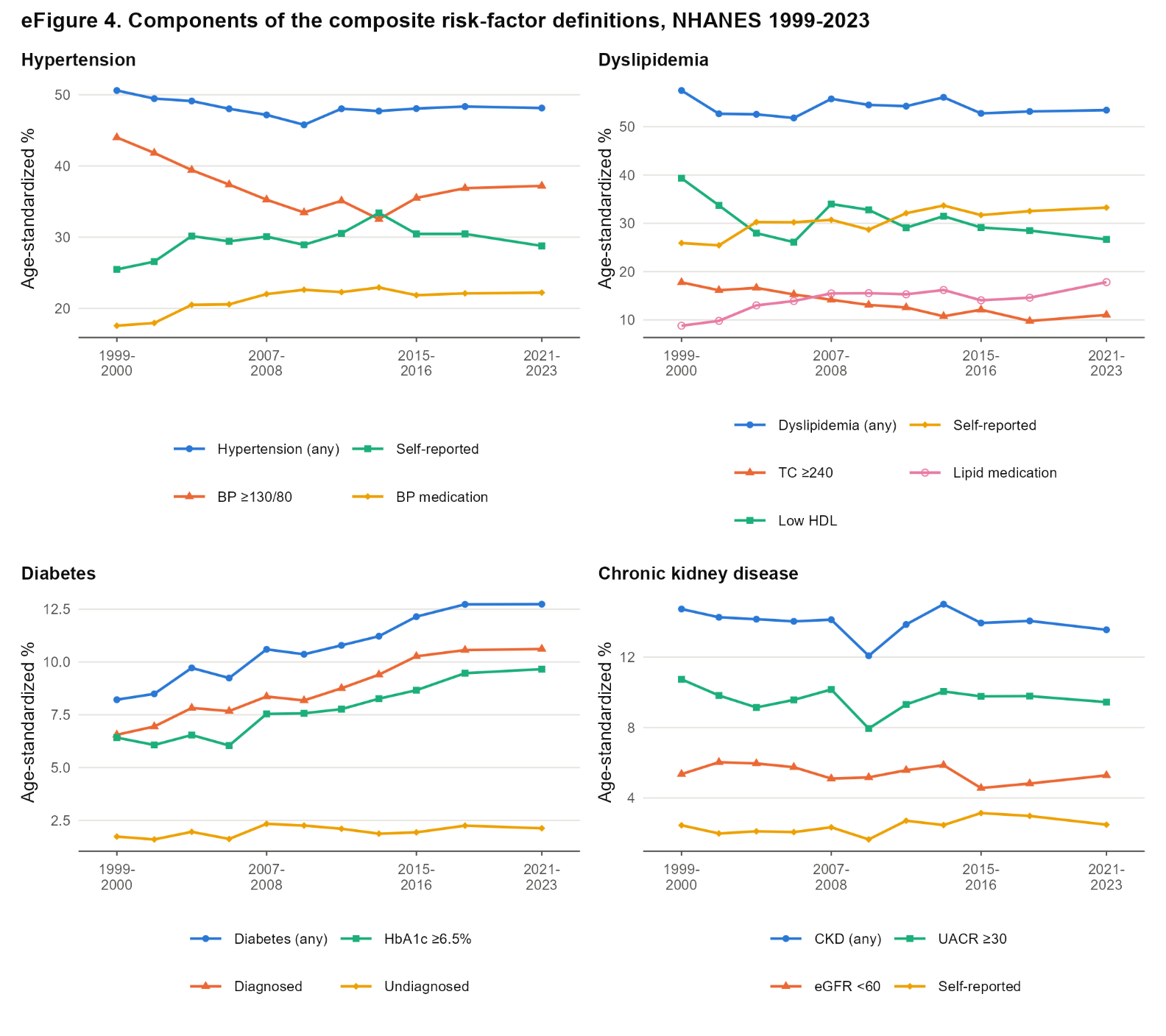

**eFigure 5. Primary risk factors by sex**

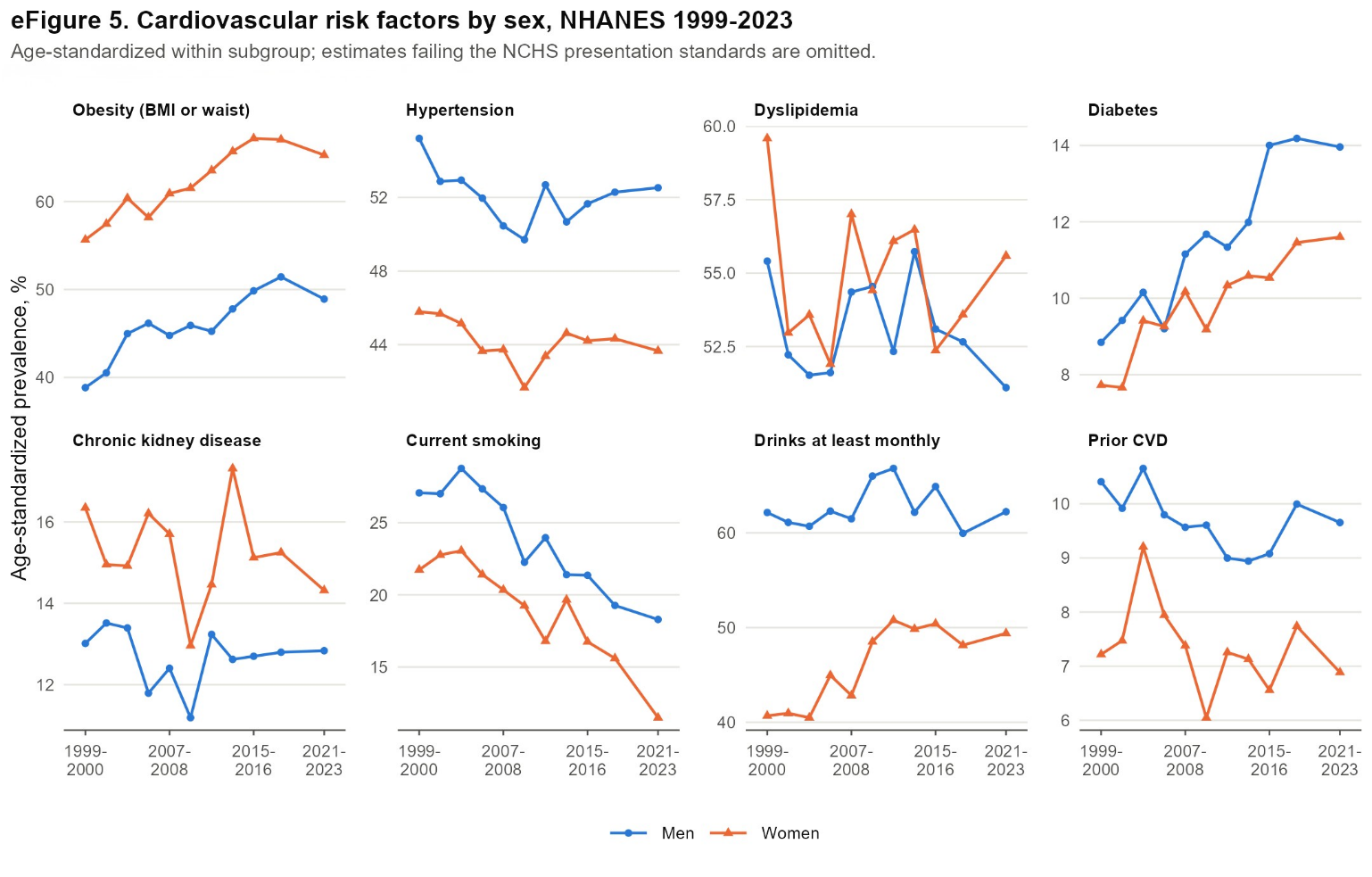

**eFigure 6. Primary risk factors by age group (crude within age group)**

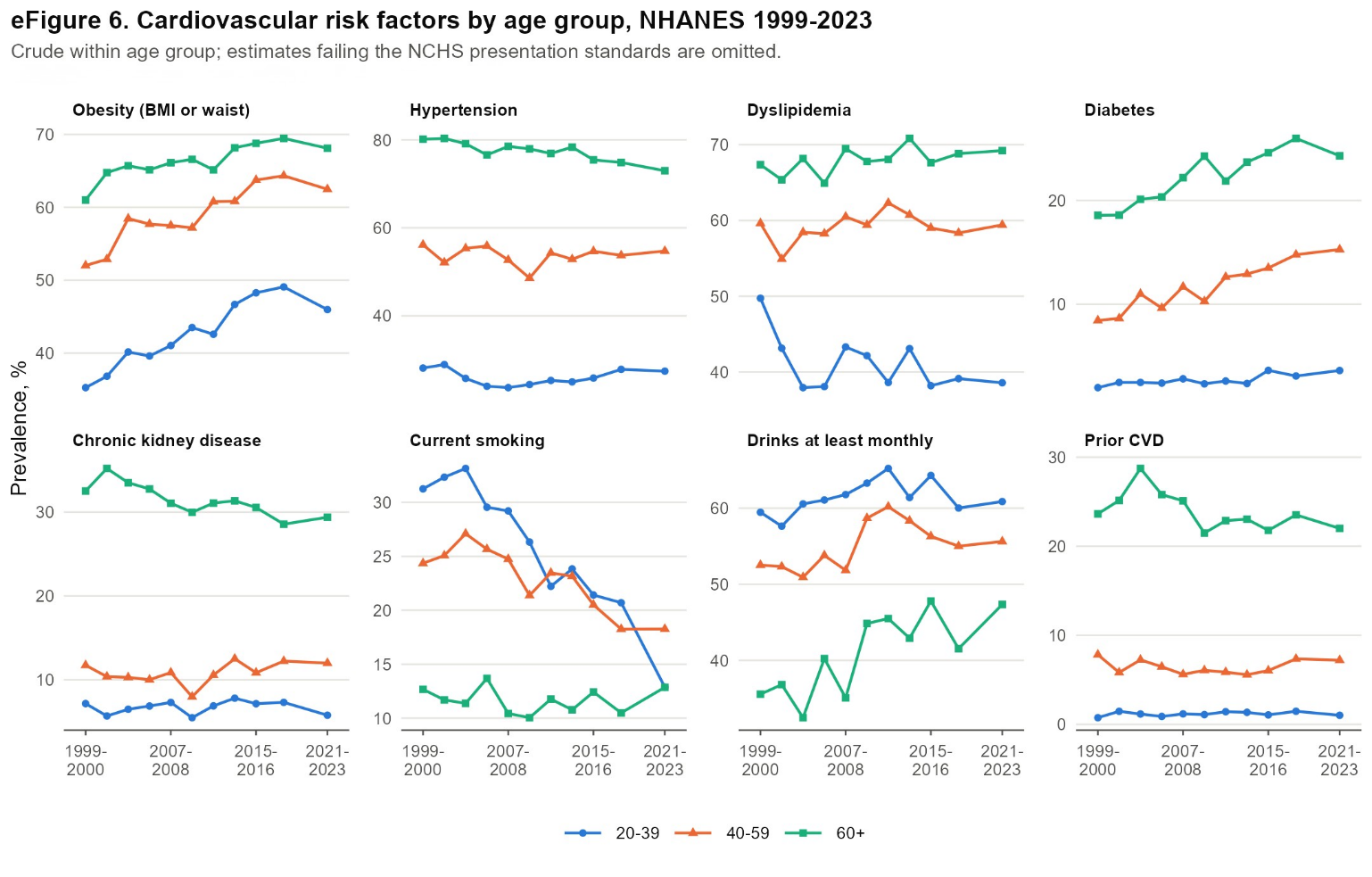

**eFigure 7. Primary risk factors by race and Hispanic origin**

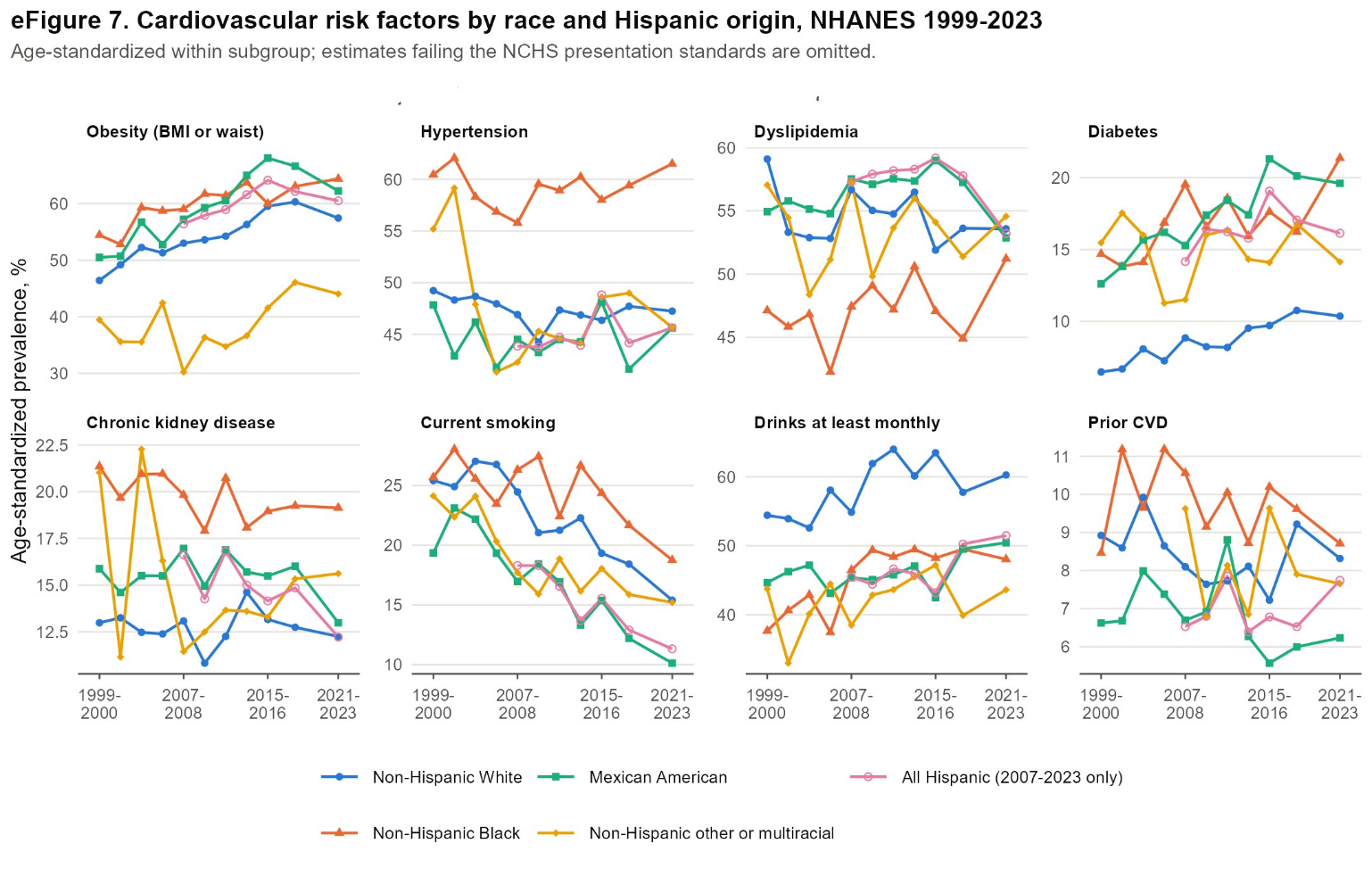

**eFigure 8. Primary risk factors by family income-to-poverty ratio**

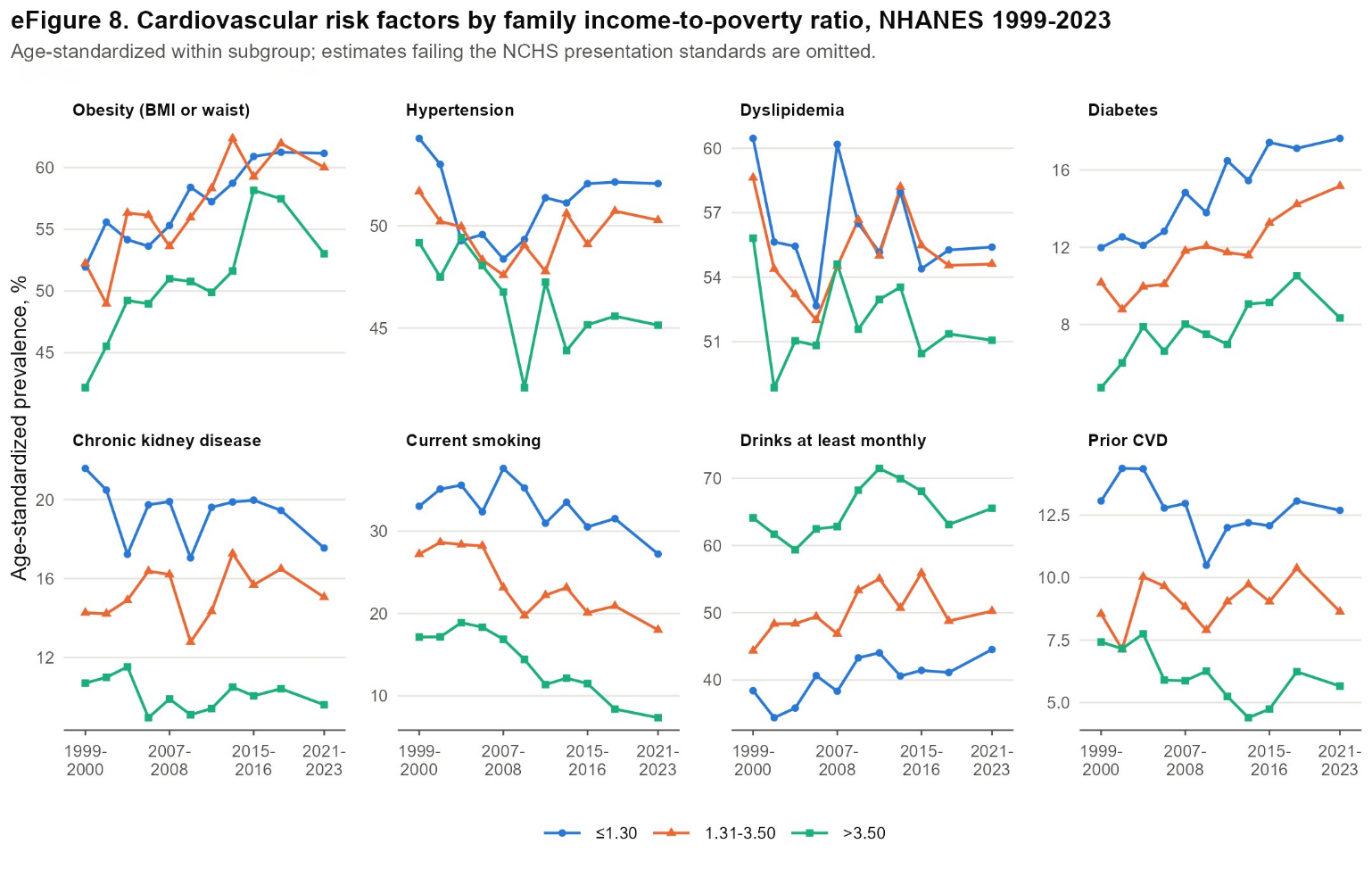

**eFigure 9. Primary risk factors by education**

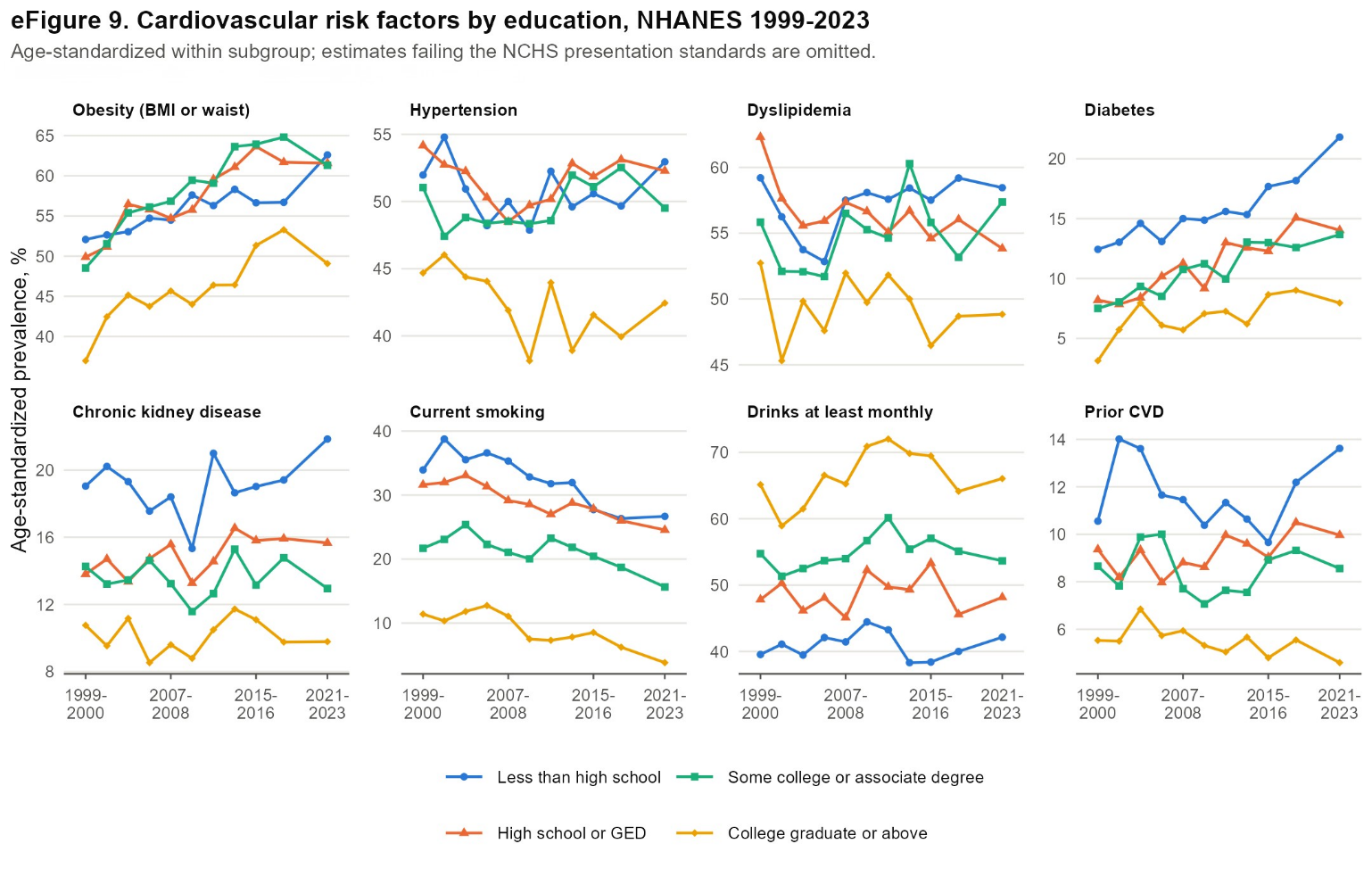

**eFigure 10. Sensitivity analyses: odds ratio per decade for primary and alternative analyses**

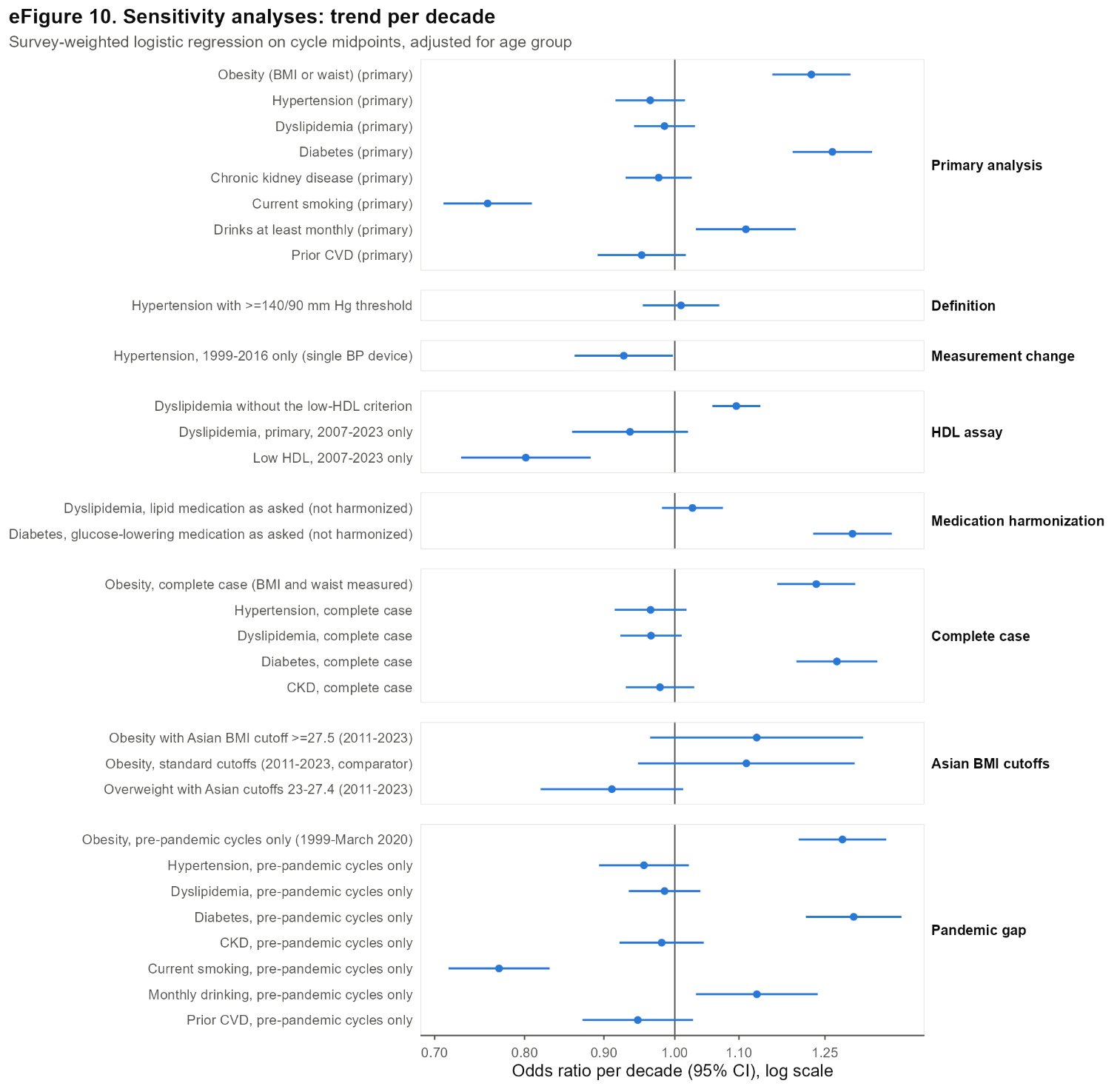

**eFigure 11. HDL cholesterol assay changes and the dyslipidemia trend**

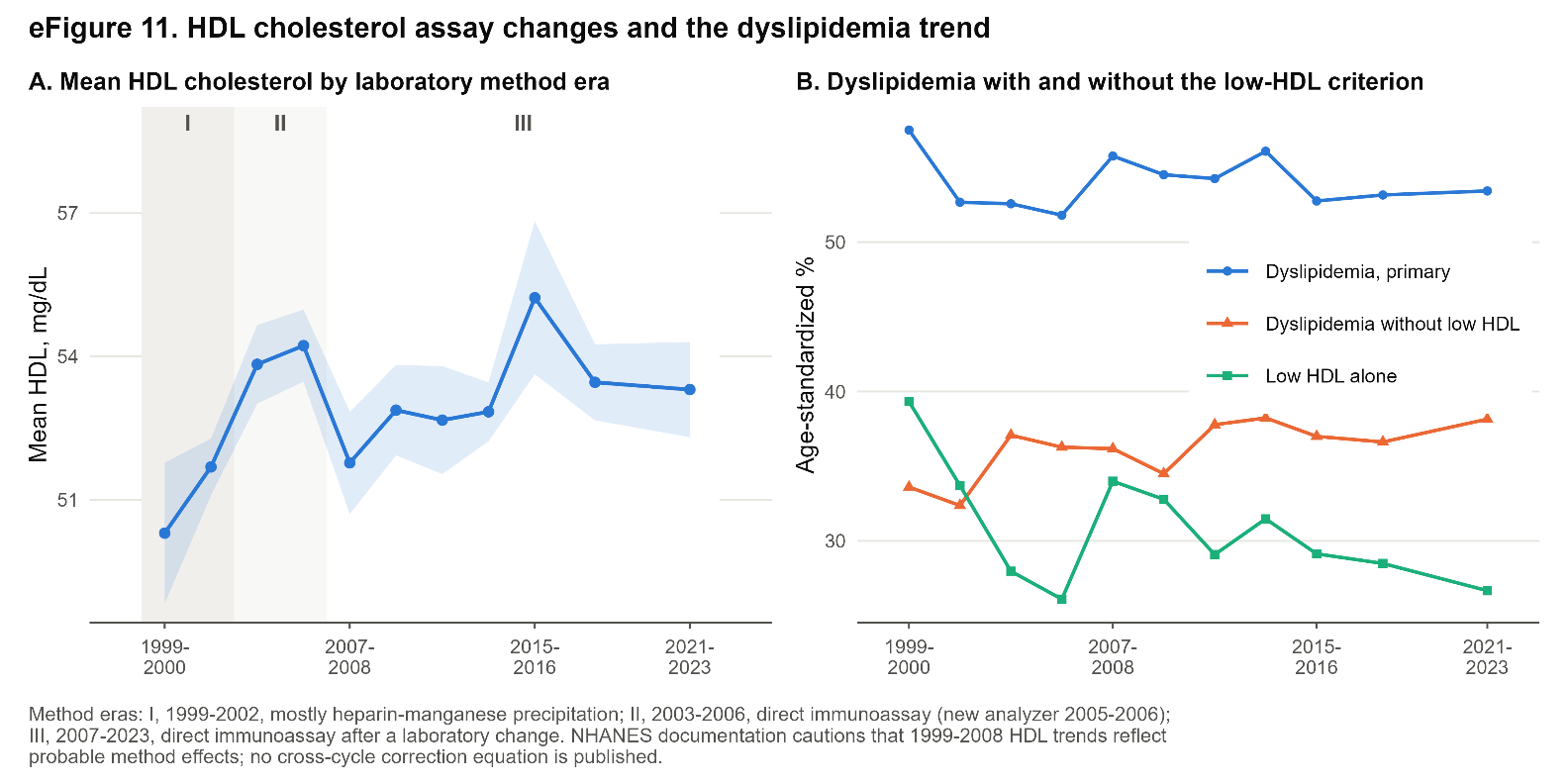

**eFigure 12. Effect of harmonizing the medication questions across cycles**

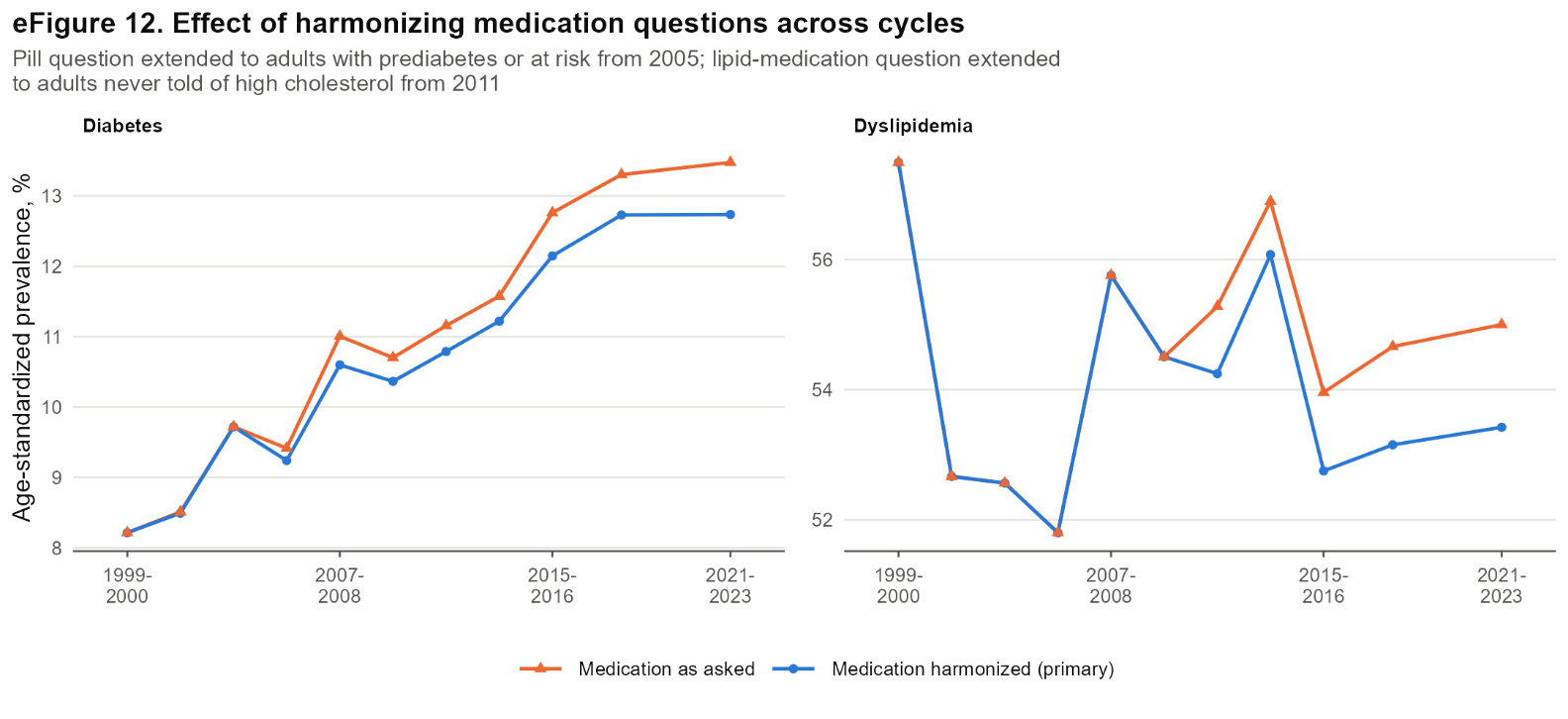

**eFigure 13. Joinpoint regression of the primary outcomes**

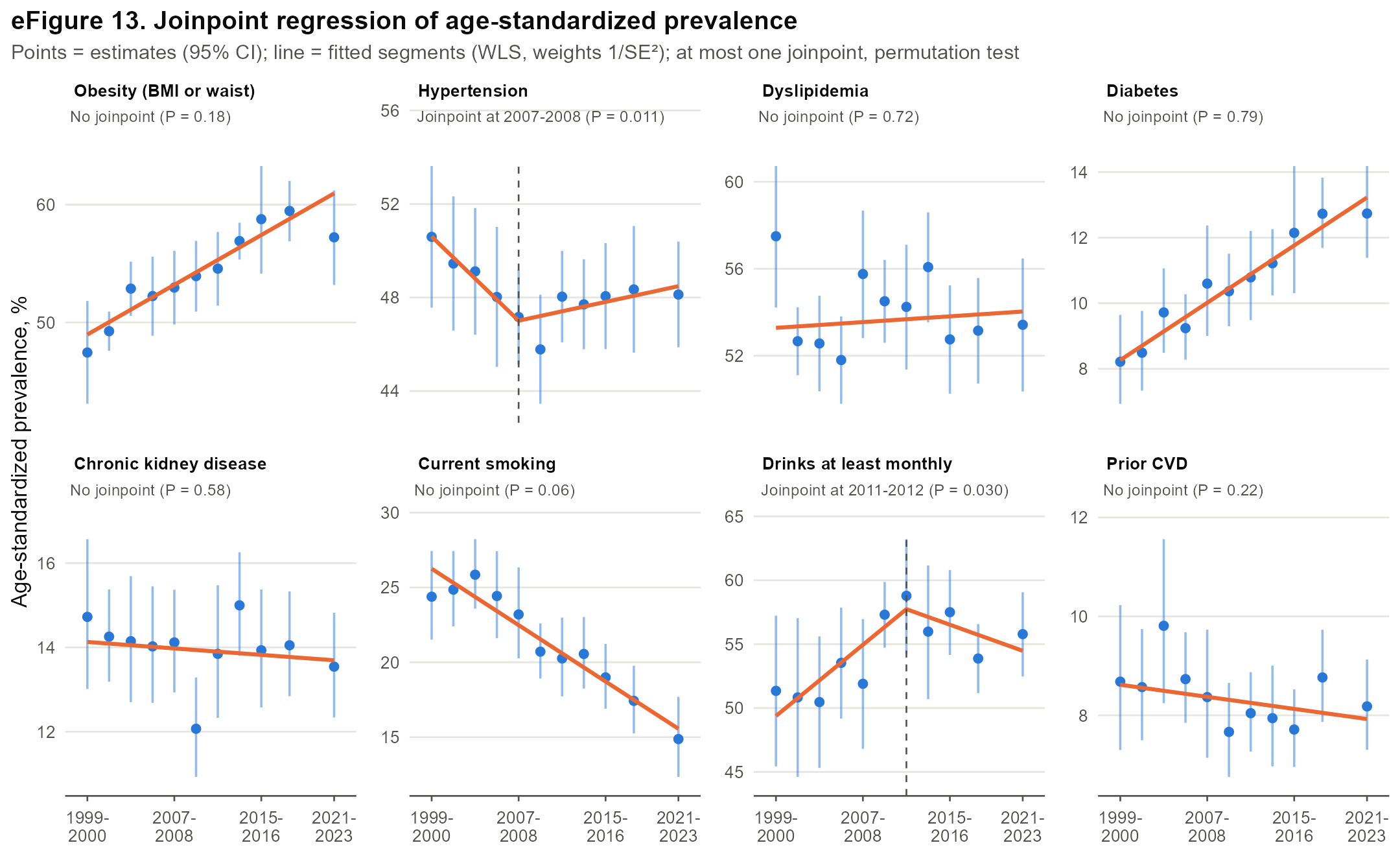

**eFigure 14. Distribution of 10-year PREVENT total CVD risk**

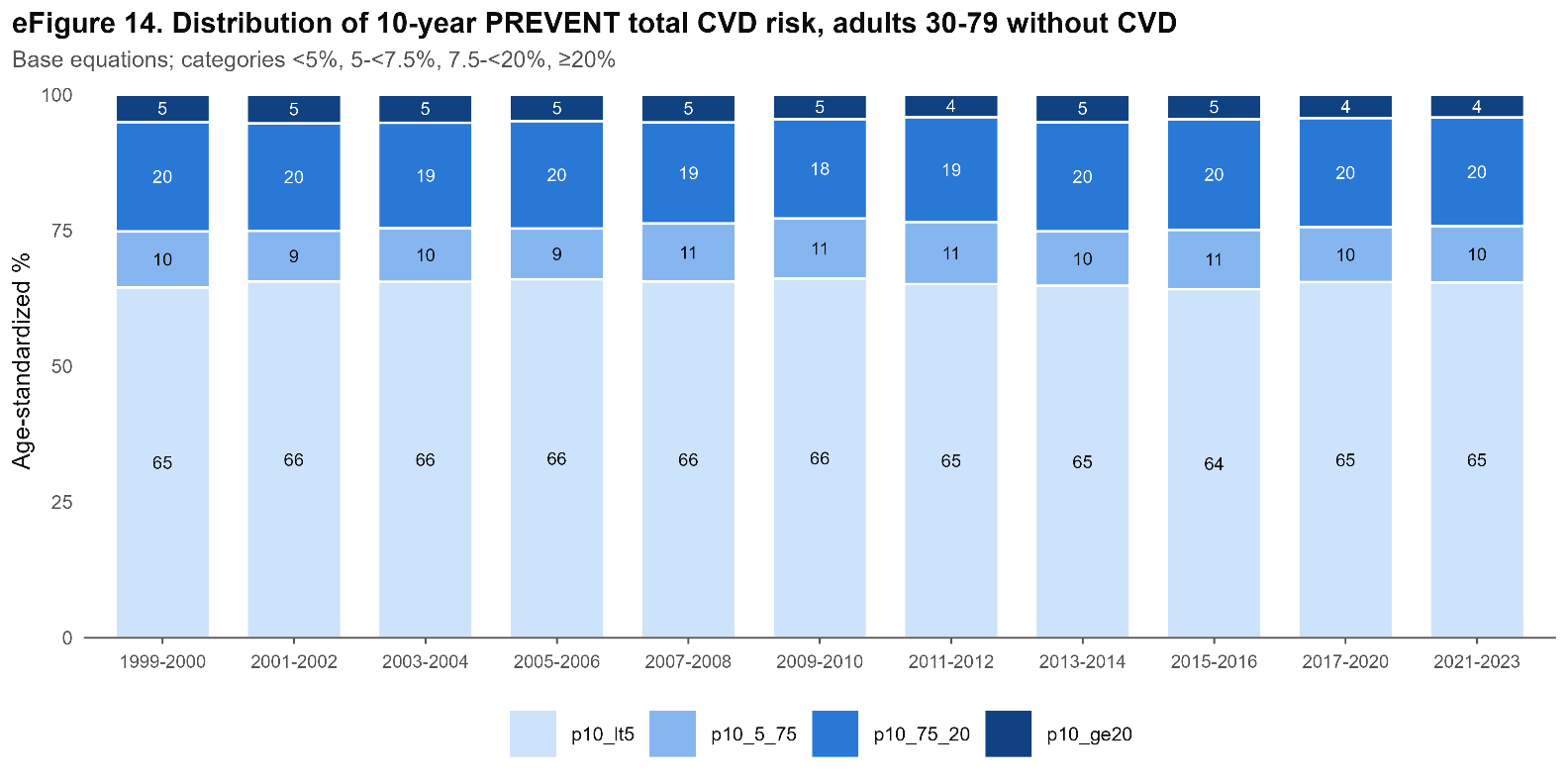

**eFigure 15. Smoking, alcohol use and prior CVD components**

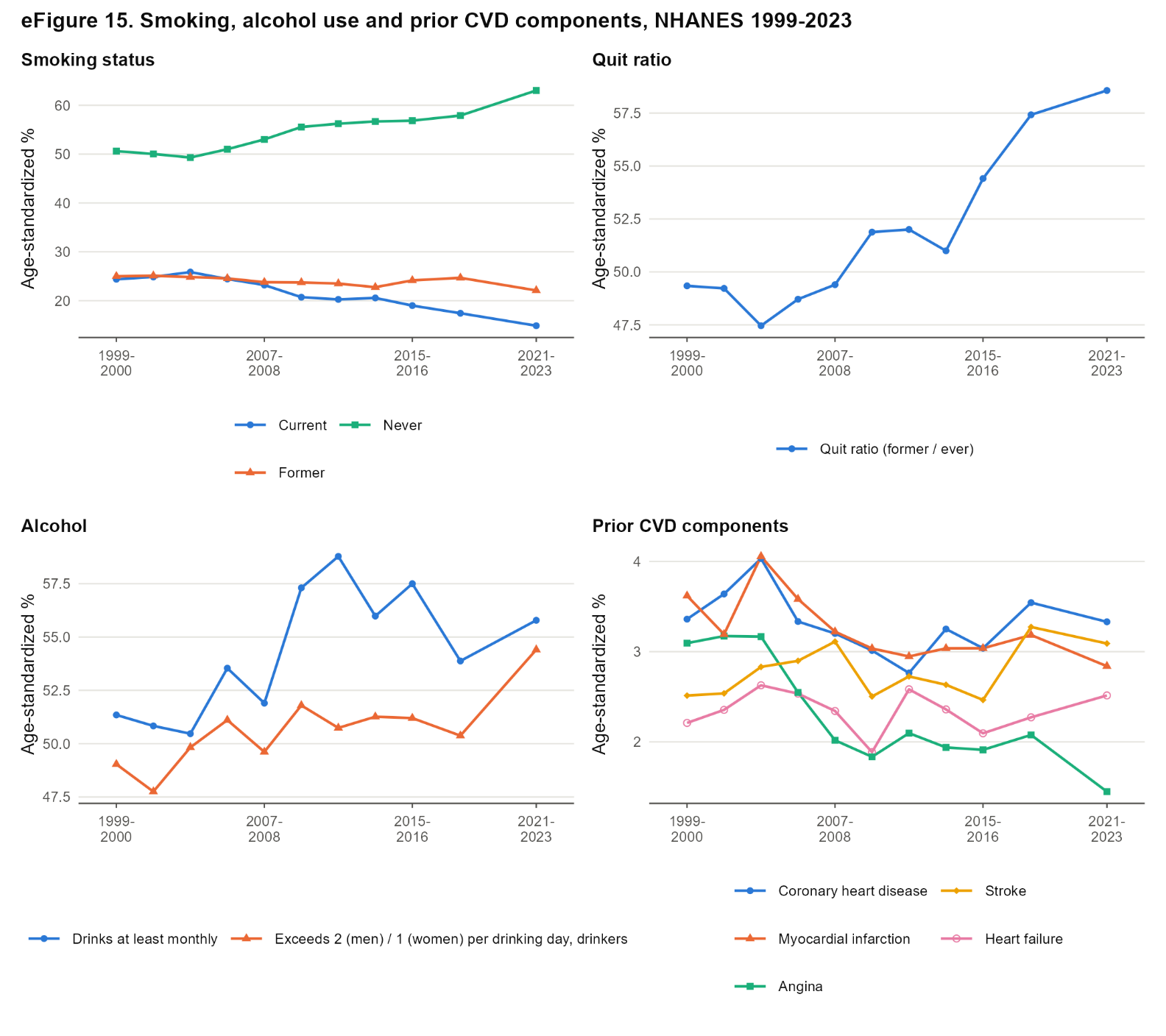

**eFigure 16. Item missingness among eligible adults, by cycle**

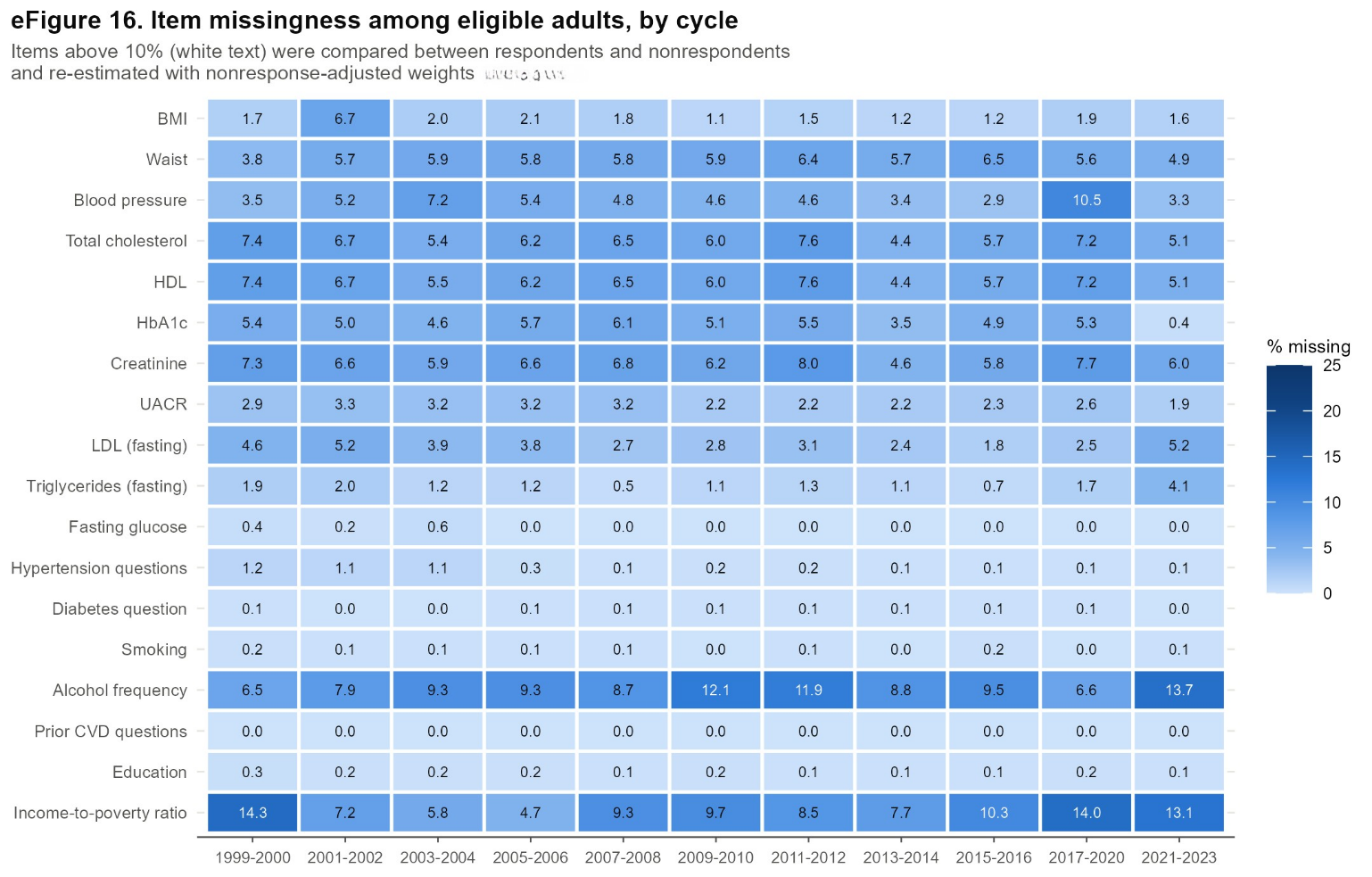

**eFigure 17. Primary and extended (fasting) definitions of dyslipidemia and diabetes**

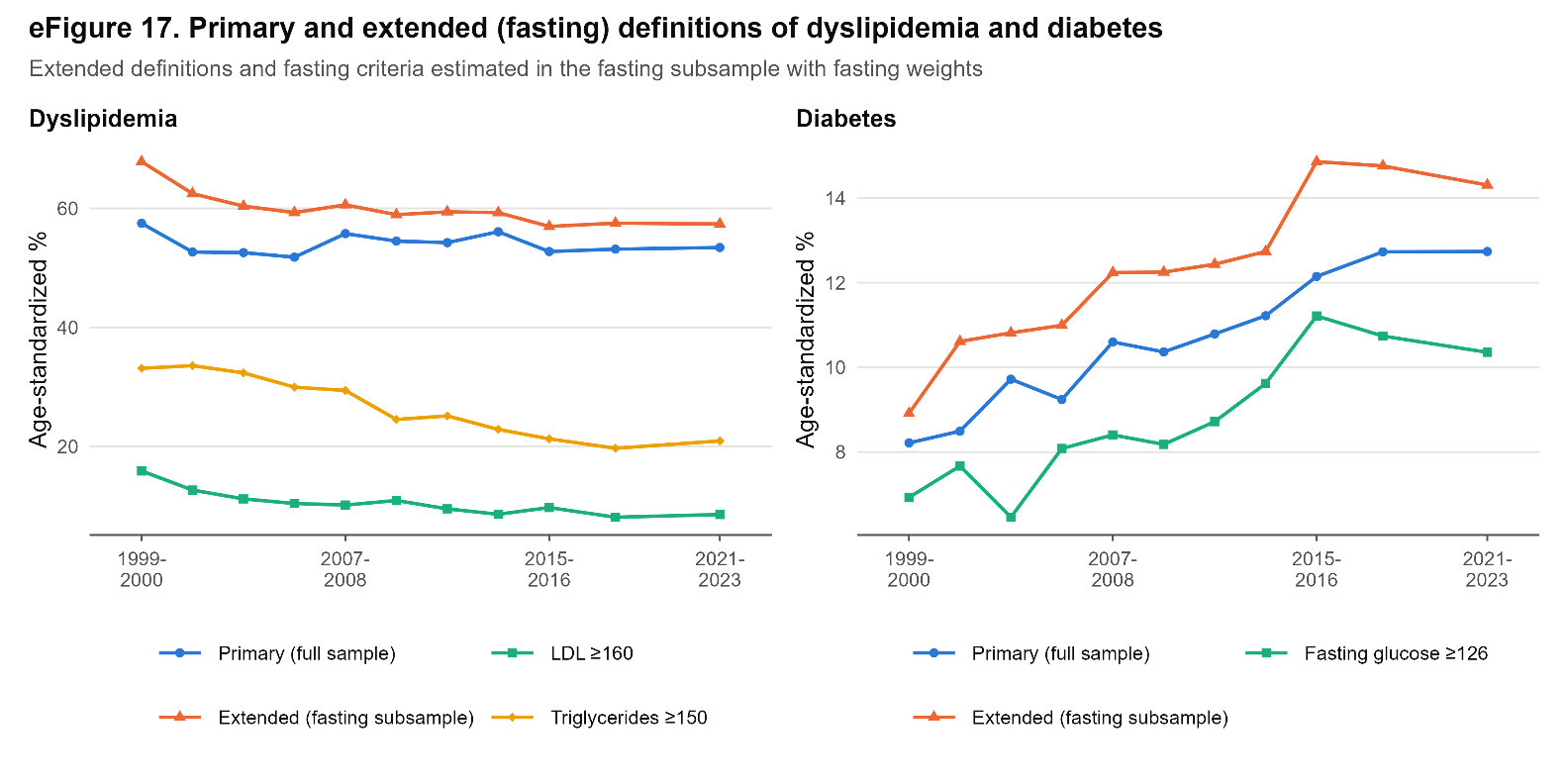
